# Explainable AI as a Double-Edged Sword in Dermatology: The Impact on Clinicians versus The Public

**DOI:** 10.64898/2025.12.19.25342205

**Authors:** Xuhai Xu, Haoyu Hu, Haoran Zhang, Will Ke Wang, Reina Wang, Luis R. Soenksen, Omar Badri, Sheharbano Jafry, Elise Burger, Lotanna Nwandu, Apoorva Mehta, Erik P. Duhaime, Asif Qasim, Hause Lin, Janis Pereira, Jonathan Hershon, Paulius Mui, Alejandro A. Gru, Noémie Elhadad, Lena Mamykina, Matthew Groh, Philipp Tschandl, Roxana Daneshjou, Marzyeh Ghassemi

## Abstract

Artificial intelligence (AI) is increasingly permeating healthcare, from physician assistants to consumer applications. Since AI algorithm’s opacity challenges human interaction, explainable AI (XAI) addresses this by providing AI decision-making insight, but evidence suggests XAI can paradoxically induce over-reliance or bias. We present results from two large-scale experiments (623 lay people; 153 primary care physicians, PCPs) combining a fairness-based diagnosis AI model and different XAI explanations to examine how XAI assistance, particularly multimodal large language models (LLMs), influences diagnostic performance. AI assistance balanced across skin tones improved accuracy and reduced diagnostic disparities. However, LLM explanations yielded divergent effects: lay users showed higher automation bias – accuracy boosted when AI was correct, reduced when AI erred – while experienced PCPs remained resilient, benefiting irrespective of AI accuracy. Presenting AI suggestions first also led to worse outcomes when the AI was incorrect for both groups. These findings highlight XAI’s varying impact based on expertise and timing, underscoring LLMs as a “double-edged sword” in medical AI and informing future human-AI collaborative system design.

## Introduction

Artificial intelligence (AI) has been intensively investigated as a tool to enhance clinical decision-making. Dermatology is one area with several developed and FDA-approved tools such as Nevisense^1^ and DermaSensor^2^, as skin conditions are primarily diagnosed through image assessment. In light of the national shortage of dermatologists^3,4^, effective AI assistance could help to improve early detection rates and reduce unnecessary clinical visits. The development of AI-powered interfaces has also been proposed to assist the general public in making informed healthcare decisions (e.g., self-diagnosis of skin diseases with Google Lens^5^)

Explainable AI (XAI) has been used in health generally to target AI usability and adoption^6–9^. In dermatology, physicians have clear features to look for, such as the ABCD’s (asymmetry, irregular borders, multiple colors, diameter greater than a pencil eraser) of melanoma. Similarly, in AI, gradient-weighted class activation mapping (GradCAM)^10^ and content-based image retrieval (CBIR)^11,12^ have been used to highlight relevant image regions and retrieve similar cases, respectively. A recent survey found that GradCAM and CBIR are the top two most commonly used XAI techniques in dermatology^13^. In addition, with the recent surge of generative AI^14^, multimodal large language models (multimodal LLMs) operating over textual and visual modalities^15–18^ have also been used to analyze dermatological images and explain AI decisions^19–21^.

Unfortunately, prior work has shown that XAI can increase subject over-reliance on AI in decision-making^22–24^, and human-AI collaborative medical decisions do not always surpass those made by humans or AI alone^25,26^. To date, there is no consensus about whether medical experience improves human-AI collaborative diagnosis. While some research indicates that medical knowledge is needed for better AI resilience and clinical decision making^27^, there is also research showing it makes minimal difference^12^, and in dermatology, AI may even mislead humans and lead to worse outcomes in diagnosis^28–30^. As dermatological AI tools expand, it is crucial to understand how different XAI methods, especially with the vast spread of LLMs, affect skin disease diagnostic accuracy across both the general public and medical experts^31–35^.

In this paper, we designed two large-scale experiments to systematically investigate how different XAI methods and human-AI decision paradigms impact diagnostic performance across expertise levels. We chose clinical-image-based skin condition diagnosis as a plausible real-world scenario (i.e., ecological validity) given the influx of patient-facing and physician-facing diagnostic models in this space^30,36^. We investigated the overall effectiveness of XAI assistance in improving dermatological diagnostic accuracy^37,38^, reducing the disparities across skin tones^39^, and influencing accuracy-confidence calibration (i.e., accuracy and confidence are consistent). We compared both correct and incorrect multimodal LLM-based explanations to traditional XAI approaches in both scenarios. We also examined how individual differences in AI deference (i.e., the propensity to follow AI regardless of accuracy, sometimes referred to as AI susceptibility^40^) impact diagnostic performance. Finally, we explored the impact of the human-AI decision paradigm (i.e., the order of human or AI making decisions) on diagnostic outcomes^41,42^, providing insights into optimal implementation strategies for clinical settings.

We explore these questions in two populations. First in the general public (N=623) with a binary classification task to distinguish melanoma and nevus, and second with medical providers (N=153 primary care physicians [PCPs]) on a more challenging open-ended differential diagnosis of skin diseases. Our findings have important implications for designing and deploying image-based medical AI systems for skin diseases. Our work comprehensively studies the collaboration between explainable AI and both the general public and medical experts. By elucidating how different traditional (GradCAM, CBIR) and advanced XAI methods (multimodal LLM), levels of medical expertise, and human-AI decision paradigms influence outcomes, we make unique contributions to the understanding of human-AI collaborative decision making and the development of more effective and appropriate human-AI collaborative medical systems.

## Results

### Study Design

#### Study Populations

We designed two complementary large-scale digital studies to evaluate human-AI collaborative diagnostic performance (Fig. 1a). Both studies were based on clinical images and designed to resemble real-world practices: a regular lay person may take a photo of their skin and resort to search engines or AI tools^43,44^, and an expert often needs to make a differential diagnosis based on a clinical image (e.g., patient communication through electronic health record messaging systems). To ensure data quality, we provided comprehensive tutorial materials for each participant group.

**Fig. 1.**
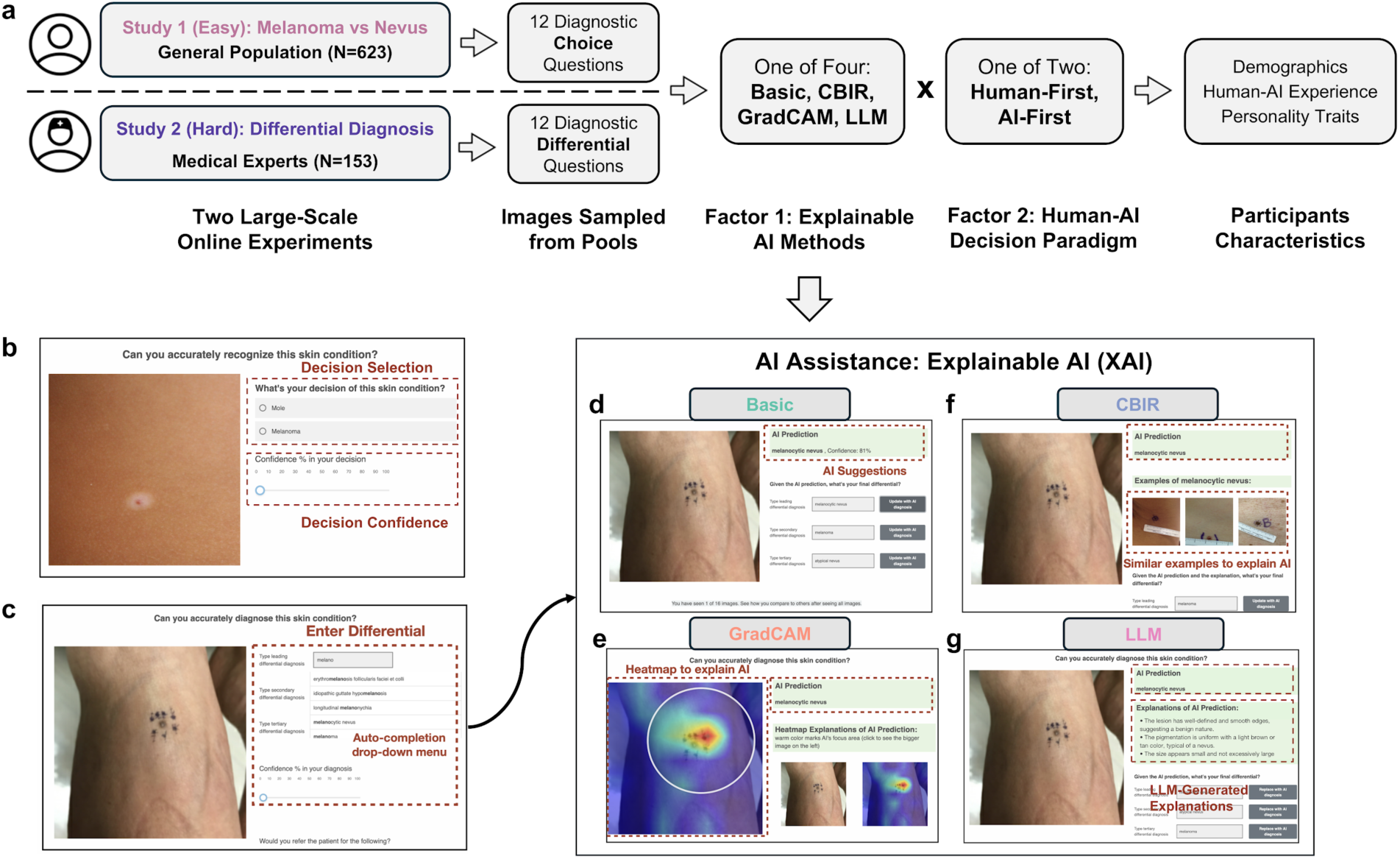
Study Design and Interface Components of Human-AI Collaborative Diagnostic System. **a**, Overview of the experimental workflow comprising two studies: Study 1 recruited general public participants for melanoma versus nevus classification tasks, while Study 2 engaged medical experts for open-set differential diagnosis. 12 diagnostic images were randomly drawn from a high-quality image database with an equal skin tone split. Participants completed either human-first or AI-first diagnostic sequences followed by demographics, human-AI collaboration experience, and personality assessment. **b**, Interface for the binary classification task showing a clinical image for melanoma vs. nevus discrimination. **c**, Interface for the differential diagnosis task with top-3 free-text entry. To assist user input, the text box has a string-matching-based auto-completion function that covers a comprehensive set of 445 skin diseases. **d**-**g**, Four XAI assistance examples with (**d**) Basic AI providing straightforward predictions with model confidence, (**e**) GradCAM highlighting relevant image regions, (**f**) CBIR showing similar reference cases, and (**g**) LLM providing natural language explanations. **Abbreviations**: AI, artificial intelligence; XAI, explainable AI; CBIR, content-based image retrieval; GradCAM, gradient-weighted class activation mapping; LLM, large language model.

Study 1 engaged 623 subjects (314 females, 305 males, 4 non-binary or others, aged 33±9) from the general public without medical backgrounds, in which participants reviewed clinical images of skin and performed a binary classification task to distinguish melanoma vs. nevus (Fig. 1b). The general public assessed 12 images that were randomly sampled from a pool of 82 images. These **images** were evenly distributed between light skin (Fitzpatrick 1-4) and dark skin (Fitzpatrick 5-6) patients^45^ and also evenly distributed between nevus and melanoma. To ensure consistent assessment of human-AI collaboration while maintaining ecological validity, we preserved AI model outputs while strategically sampling images to maintain a consistent accuracy of 83.3% (always 10 correct and 2 incorrect predictions per participant), which is close to the model’s actual performance (see AI model details below). Each participant went through 12 clinical images and made two rounds of decisions per image (with/without AI).

Study 2 involved 153 PCPs (66 females, 84 males, 3 non-binary or others, aged 34±10, years of practice experience range from 1 to 25 years, average experience 6 years). They performed a more complex open-ended differential diagnosis with free-text entry. Participants were asked to enter their top three diagnoses (Fig. 1c). In Study 2, we focused on four main conditions previously identified as having potential diagnostic disparities across skin tones (better performance on patients with light skin than those with dark skin)^30,46^: atopic dermatitis, pityriasis rosea, Lyme disease, and cutaneous T-cell lymphoma (CTCL). To simulate real-life applications, these four conditions were mixed with the other 30 common skin conditions. Text entry was assisted by auto-completion based on string matching from a comprehensive list of 445 skin diseases. Each participant evaluated 12 cases, including 2 images for each of the four main conditions with correct AI predictions, 2 images sampled from the four main conditions with incorrect AI predictions, and 2 images sampled from other conditions with a mix of AI correctness (expected 1.5 correct and 0.5 incorrect), resulting in expected AI predictions to be 9.5 correct and 2.5 incorrect (i.e., expected accuracy of 79.2%). Similar to Study 1, skin tone was balanced in each condition.

We note that Study 1 and 2 are not directly comparable as they emphasize different tasks appropriate for groups with different medical expertise. In order to measure the impact of medical training experience within the same task, we additionally recruited 320 medical students (171 females, 141 males, 8 non-binary or others, aged 28±9) with less expertise in dermatology to compare to the cohort reported in Study 2 (see EFig. 4 & 5).

We further employed validated questionnaires to collect additional data on human-AI collaboration experience^47^, XAI quality^48^, and cognitive traits of critical thinking^49^ and open-mindness^50^. More details can be found in the experiment design sections in Methods. Overall, we collected 14,952 diagnostic decisions from the general public (Study 1), 3,672 diagnoses from PCPs, and 7,680 diagnoses from medical students (Study 2).

#### Human-AI Decision Paradigm

For both studies, we adopted a between-subject factorial design with 4 x 2 conditions: including four XAI methods (**Basic** outcomes with model prediction and confidence, **GradCAM**: highlighting which areas contribute to the model prediction, **CBIR**: presenting similar images with the same label as the prediction, and **multimodal LLM**: semantic explanations of the reason for the model prediction, Fig. 1d-g), For the rest of the paper, we omit “multimodal” and use **LLM** for simplicity, as our studies mainly focus on the language capability of the multimodal LLM, but do note we used a vision-language model GPT-4V, see Methods for more details), as well as two human-AI decision paradigms (**Human-First,** where users make a decision first before reviewing AI suggestions, and **AI-First**, where users review both images and AI suggestions before making the final decision). Each participant was randomly assigned to one condition and went through 12 clinical images. Since the Human-First condition naturally required participants to make two rounds of decisions (the first round without AI and the second round with AI), to control the effect of cognitive effort, participants also made two decision rounds in AI-First: after making the first round of decision with AI, they were asked to review and reconsider their final decisions in the second round when AI results were hidden. Besides decisions, participants also rated their confidence in their decisions at each round.

#### Model Training

We trained a deep-learning model ViT-B/32^51^ for the binary classification task in Study 1. We applied the training strategy of conditional domain adversarial neural network (CDANN)^52^ to achieve the fairness constraint and ensure performance balance between skin tones (weighted AUROC of 0.930, accuracy Δ=2.1% between skin tones). In Study 2, we trained a DenseNet-121^53^ with an AUROC of 0.772, accuracy Δ=5.6% between skin tones. Post-hoc explanations (GradCAM, CBIR, and LLM) were generated using model outputs. See the Methods section for more details.

#### Evaluation Metrics

Responses for Study 1 were evaluated using standard binary outcome measures. For the free-text task in Study 2, two authors with medical expertise manually evaluated all text entered in the “Diagnosis” section to determine the top-1 (based on the first entry) and top-3 accuracy (based on all three entries) compared to the ground truth.

All AI-generated explanations for GradCAM, CBIR, and LLM were evaluated by three dermatologists with at least 2 years of experience and independently scored for correctness (i.e., how accurate the explanation is based on the skin image) and informativeness (i.e., how much this explanation can support diagnosis) on a 5-point Likert scale, achieving fair to moderate interrater reliability (Fleiss’ κ=0.42 and 0.37). We calculated the average and divided explanation quality into high (informativeness score ≥ 3 and correctness score ≥ 3) and low (informativeness score < 3 or correctness score < 3, see examples of high and low-quality explanations in SFig. 7-12).

### SOTA AI Improves General Public’s Performance Due to AI Deference, LLM Explanations Amplify Such Deference

***Advanced AI improves the general public’s performance.*** We first measured the general public’s performance (Study 1) without and with AI assistance. We found that AI improved average accuracy of the nevus vs. melanoma detection task from 69.7±0.8% to 75.8%±0.7% (effect of AI assistance: β=0.061, 95%CI=[0.049, 0.074], p<0.001, linear mixed model on accuracy, with AI assistance, XAI methods, and their interaction as the main factor, controlling gender, age, race, skin disease experience, and the covariate of self-reported human-AI collaboration experience. See STab 7 for details. Other Study 1 statistical models below control the same set of confounders unless noted differently), and the majority of the improvement came from nevus classification (β=0.111, 95%CI=[0.091, 0.132], p<0.001, STab 8, Fig. 2a) and the largest improvement in explainable AI comes from multimodal LLM (see next section). Participants also had a modest increase in diagnosis confidence by 1.5% with AI assistance (β=0.018, 95%CI=[0.010, 0.019], p<0.001, EFig. 1a, STab 10). With the help of AI model humans achieved a more balanced diagnosis performance across patient skin tones (Round 1: β=0.033, 95%CI=[0.009, 0.057], p=0.007; Round 2: β=0.017, 95%CI=[−0.007, 0.041], p=0.166, Δ*_rel_*=46.9%, Fig. 2c, STab 11). These findings demonstrate that collaboration with well-trained AI has the potential to mitigate diagnostic biases while improving overall accuracy.

**Fig. 2.**
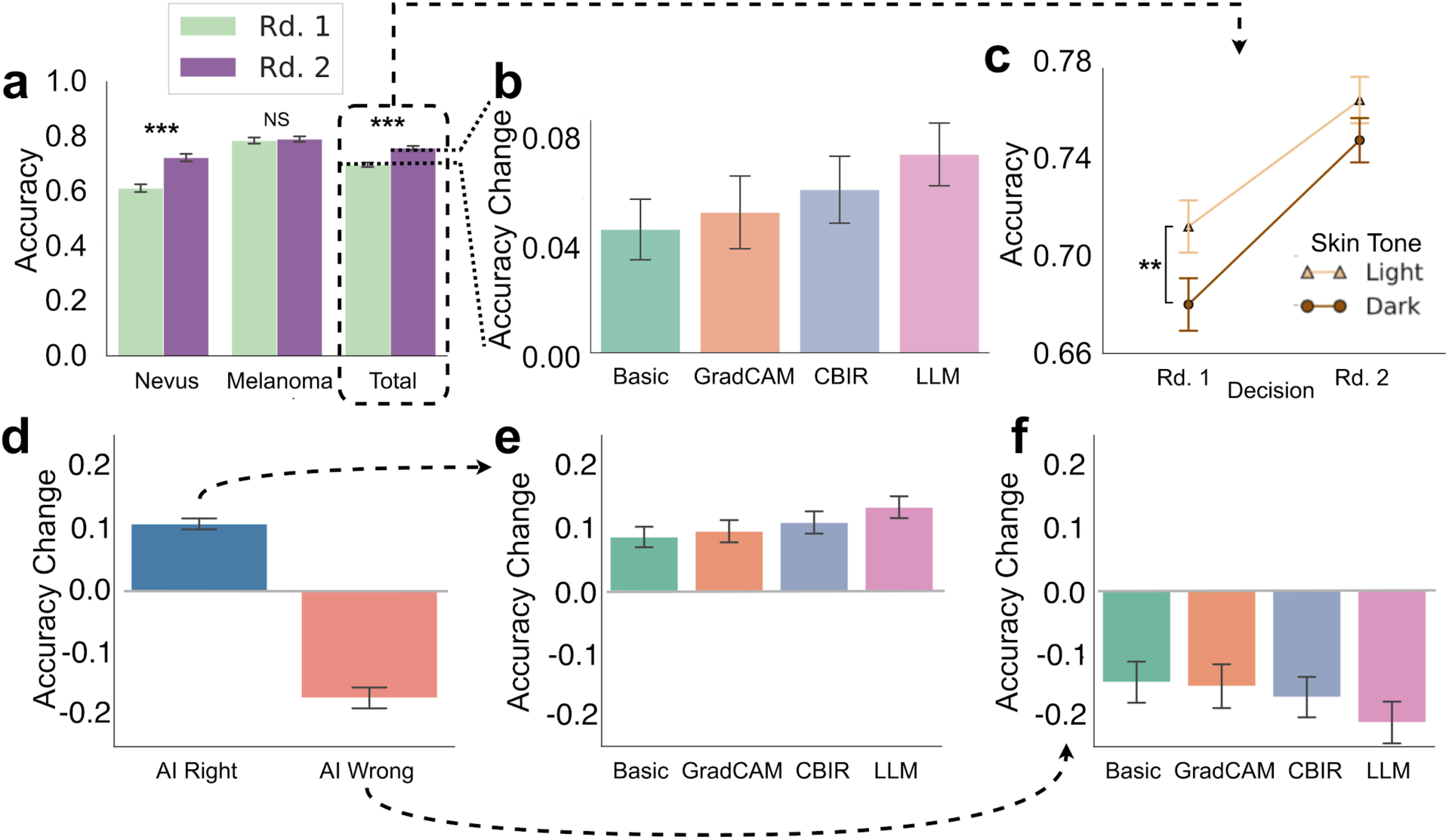
Overall Diagnostic Performance with AI Assistance for the General Public. **a**, Accuracy of the general public in detecting nevus, melanoma, and the overall diagnosis in two rounds: Rd. 1 (initial, human accuracy without AI assistance) and Rd. 2 (AI-assisted accuracy). **b**, Diagnostic accuracy changes across different AI explanations (Basic, GradCAM, CBIR, and LLM). **c**, Diagnostic accuracy when patients are light-skinned or dark-skinned across two rounds. **d**, Overall accuracy changes under AI-right and AI-wrong conditions. **e**, **f**, Diagnostic accuracy changes of different AI explanations under AI-right (**e**) and AI-wrong (**f**) predictions. All error bars in this figure (and the rest of the figures) indicate the standard error of the mean. The same metrics were applied in Fig. 3.

***Performance improvement stems from AI deference & LLM-based explanations amplify such deference.*** We investigated the impact of the four XAI methods on diagnostic accuracy. The LLM explanations provided an improvement of +7.7% (β=0.077, 95%CI=[0.053, 0.101], p<0.001), followed by CBIR (+6.3%, β=0.063, 95%CI=[0.039, 0.087], p<0.001), GradCAM (+5.5%, β=0.054, 95%CI=[0.028, 0.081], p<0.001), and the basic method (+4.8%, β=0.048, 95%CI=[0.023, 0.073], p<0.001), as shown in Fig. 2b and STab 7. However, the general public’s diagnostic accuracy improved when AI provided correct predictions, while incorrect predictions reduced the performance significantly (β=-0.233, 95%CI=[−0.307, −0.158], p<0.001, Fig. 2d, STab 12). Correct LLM advice (+13.4%) enhanced performance more than other AI explanations (basic method +8.6%, GradCAM +9.5%, CBIR +10.9%, Fig. 2e), and incorrect LLM advice decreased performance most (−21.1%, basic −14.6%, GradCAM −15.3%, and CBIR −17.0%, Fig. 2f). These findings indicate that LLM explanations amplify the general public’s tendency to follow AI guidance, regardless of AI accuracy. Moreover, compared to basic explanation, LLM leds to significantly more reduction of performance when AI becomes inaccurate (i.e., delta of detla, β=-0.048, 95%CI=[−0.093, −0.003], p=0.035, STab 13).

***Misplaced trust in LLM explanations for the general public.*** When AI predictions were correct, participants trusted LLM explanations more than other methods (Fig. 2e), and this was more noticeable when explanations were of low quality (post-hoc pairwise estimated marginal means (EMMs) comparisons, LLM over GradCAM: β=0.117, 95%CI=[0.041, 0.192], p=0.002; LLM over CBIR: β=0.092, 95%CI=[0.021, 0.163], p=0.011, EFig. 2c, STab 15). When AI predictions were incorrect, LLM explanations negatively impacted the alignment between participants’ confidence and accuracy (z=-3.788, p<0.001, two-side Fisher r-to-z test to compare correlation difference, EFig. 3c). These results further suggest people struggle to assess LLM explanation reliability and can be easily misled by LLM.

**Fig. 3.**
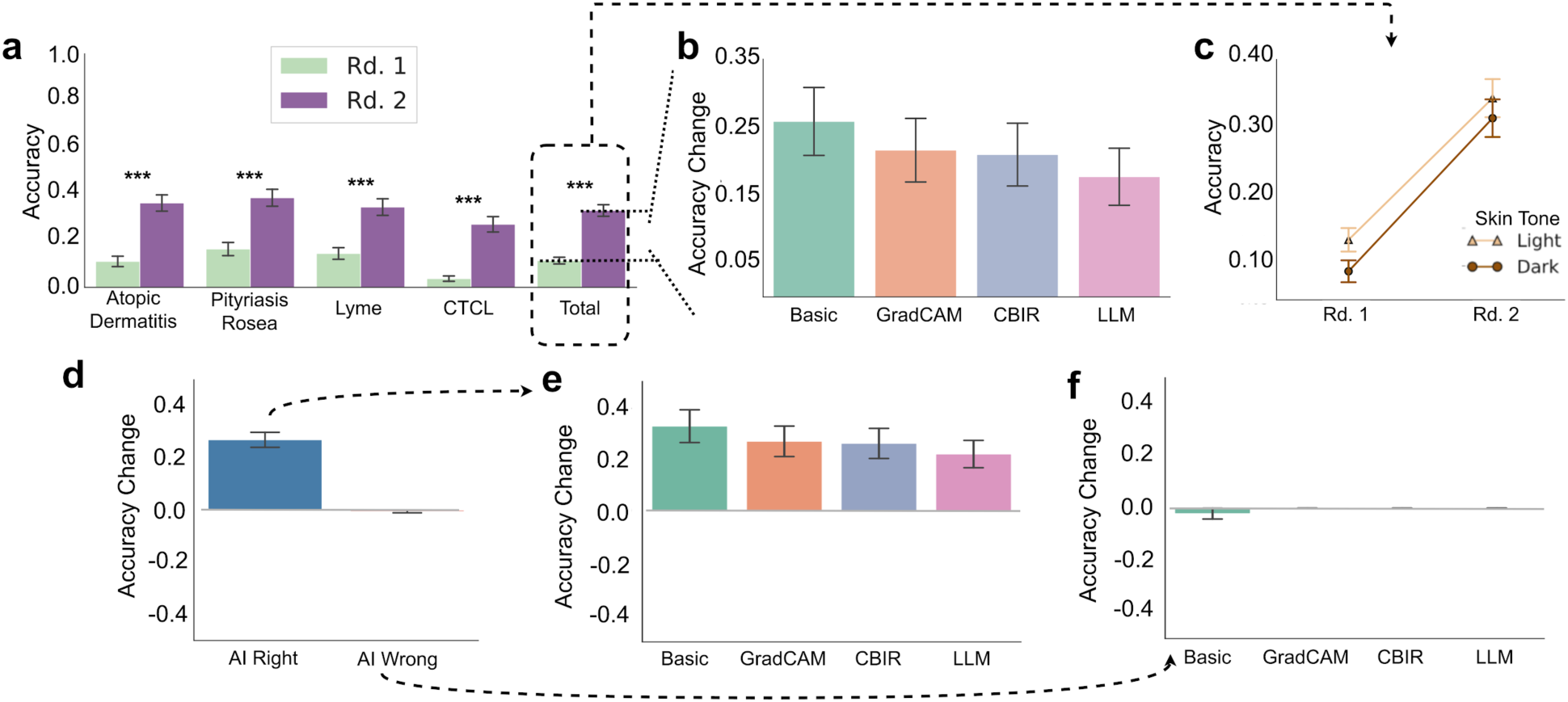
Overall Diagnostic Performance with AI Assistance for PCPs. This figure shows similar analysis results as Fig. 2. **a**, Accuracy of PCPs in identifying atopic dermatitis, pityriasis rosea, Lyme disease, CTCL, and the overall performance on four main diseases, with results presented for two rounds. **b**, Diagnostic accuracy changes across different AI explanations. **c**, Diagnostic accuracy when patients are light-skinned or dark-skinned across two rounds. **d**, Overall accuracy changes under AI-right and AI-wrong conditions. **e**, **f**, Diagnostic accuracy changes of different AI explanations under AI-right (**e**) and AI-wrong (**f**) predictions.

To summarize, our results in Study 1 indicate that **LLMs are a “double-edged sword” in skin disease diagnosis for the general public with amplified AI deference**: When AI was correct, LLM explanations boosted diagnosis performance, even when the quality of the explanation was low. However, when AI was incorrect, the general public was misled by seemingly plausible reasons generated by LLM. Although our LLM explanations rarely contained hallucinated or wrong clinical signs (i.e., the multimodal LLM is capable of visual analysis), they frequently referenced ambiguous dermatologic criteria even when these features were only partially present or visually unclear. EFig. 10 presents several specific cases when AI was incorrect and misled humans to the wrong answer. For instance, in the first case in EFig. 10a, the AI model provided a wrong prediction as melanoma while the ground truth is nevus. In this case (dysplastic nevus), the LLM-generated explanation (based on an incorrect AI prediction) was biased towards melanoma and mistakenly align abnormal nevus visual features with melanoma criteria on symmetry, color, and border (e.g., “The shape of one half does not match the other half”). To a general public without medical background, such wrong explanations appear to be plausible but are misleading. This is supported by prior work on the automation bias and algorithmic over-reliance when humans are working with AI. Our work further indicates that LLM can amplify such risks for the general public. We discuss the implications of these findings in the discussion section.

### PCPs Reliably Leverage Accurate AI Guidance, While Resisting Errors with LLM-based AI Explanation

***Basic AI improves PCP’s performance.*** We conducted a similar analysis of the PCP participants in Study 2. This task was more challenging and required detailed dermatological knowledge. We found 11.5%±1.4% top-1 accuracy and 16.1%±1.8% top-3 accuracy in PCP’s differential diagnoses without AI, which is aligned with the prior work^30^. The performance was significantly improved with AI suggestions, with a +21.5% in top-1 accuracy (β=0.258, 95%CI=[0.170, 0.260], p<0.001, Fig. 3a, linear mixed model on accuracy with AI assistance, XAI methods, and their interaction as the main factor, controlling gender, age, race, and medical expertise in skin, year of experience, and personality traits. See STab 16 for details. Other Study 2 statistical models below control the same set of confounders unless noted differently), and a +43.5% in top-3 accuracy (β=0.450, 95%CI=[0.352, 0.548], p<0.001, EFig. 6a, STab 16). In contrast to the general public in Study 1, for PCPs,basic AI assistance helped the most(top-1 accuracy β=0.258, 95%CI=[0.168, 0.349], p<0.001, Fig. 3b,STab 16). Interestingly, we only observed significant confidence increase when PCPs were assisted by GradCAM (β=0.028, 95%CI=[0.005, 0.051], p=0.019) and LLM explanations (β=0.035, 95%CI=[0.012,0.058], p=0.003, EFig. 1b, STab 21). Improvements were significant in all four major diseases (+19.9-25.0%, ps<0.001, Fig. 3a, STab 17-20). PCPs had disparate performance across skin tones as 4.6% (β=0.046, 95%CI=[0.013, 0.078], p=0.069, Fig. 3c), which was reduced to 2.9% (β=0.029, 95%CI=[−0.018, 0.076], p=0.248, STab 22) after AI assistance.

***PCPs are resilient to AI deference when AI is wrong.*** In contrast with the general public, incorrect AI predictions had minimal impact on PCPs’ final decisions across all XAI methods (Fig. 3f, EFig. 2d-2f, βs=0-0.021, ps = 0.328-1.000). This suggests PCPs relied on their own expertise and training rather than erroneous AI guidance. To control the effect of task difficulty between Study 1 and 2, we further compared PCPs’ results against the medical students’ results on the same task. EFig. 4 showed that medical students had more reliance on AI, regardless of AI correctness compared with results from PCPs. We term participants who only answered correctly when AI was correct, and therefore answered incorrectly when AI was wrong, as “deferential participants”. We observe that the proportion of deferential participants are higher among medical students than PCPs (linear mixed model on the proportion of deferential participants, with medical role and medical expertise in skin as the main factor, controlling other confounders, main effect of medical role: β=0.067, 95%CI=[0.004, 0.130], p = 0.037; main effect of skin expertise: β=0.073, 95%CI=[0.022, 0.124], p = 0.005, STab 25). These results indicate that higher expertise levels are associated with more careful AI adoption, which is supported by prior work^12^.

**Fig. 4.**
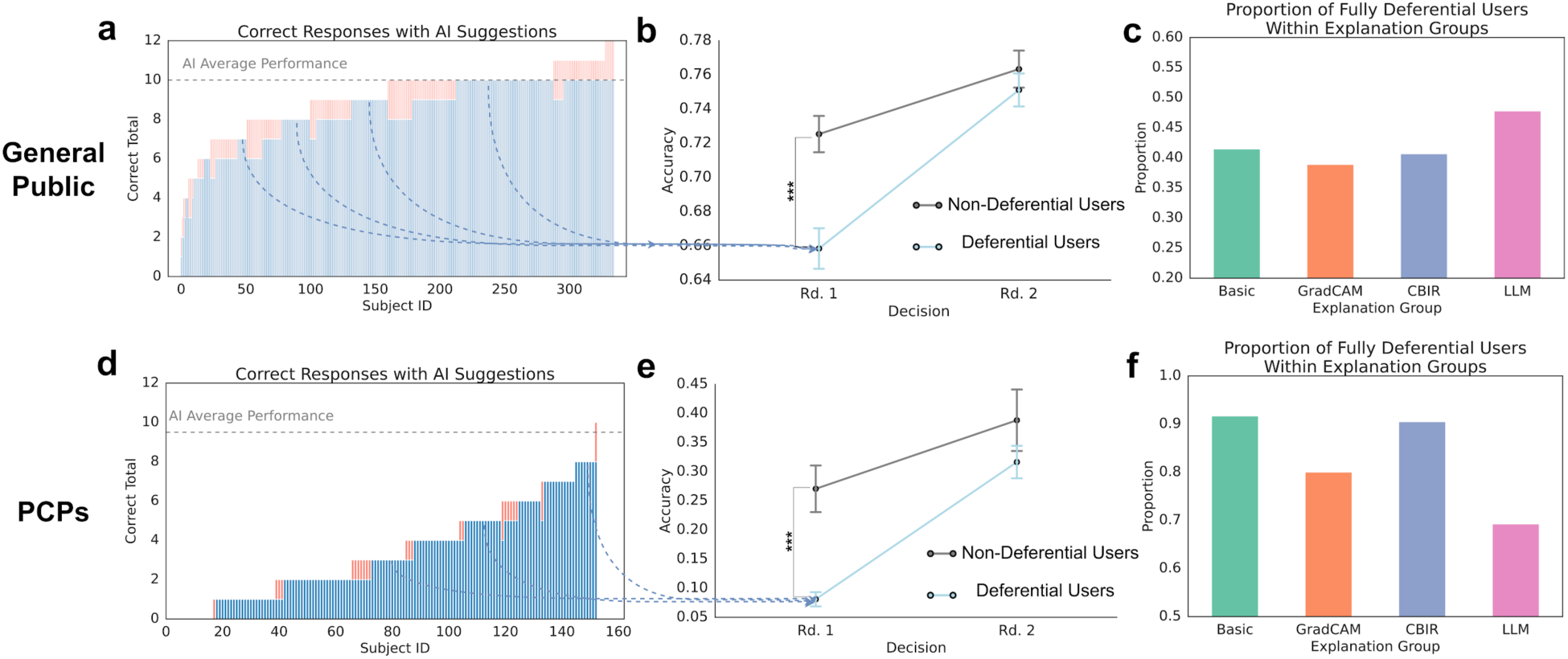
Deference Patterns towards AI in HAI Collaboration. **a**-**b**, Performance comparison between deferential (fully compliant with AI suggestions) and non-deferential general populations. (**a**) Distribution of correct responses with AI assistance. The blue bars represent instances where both AI predictions and participants were correct, and the red overlays indicate correct responses by participants while AI predictions were incorrect, (**b**) accuracy trajectories across two decision rounds. **c**, Proportion of participants that defer for each XAI output. **d**-**f**, Equivalent results for PCP participants as (**a**)-(**c**).

***LLM explanations do not aid in accuracy, but in confidence calibration.*** Interestingly, for PCPs, LLM explanations were the least helpful method (+17.7%) and were 8.1% lower than the best improvement from the Basic explanations (β=-0.081, 95%CI=[−0.207, 0.044], p = 0.268, Fig. 3b, STab 16). This finding was consistent across explanation quality (EFig. 2e, 2g, STab 27) and top-3 accuracy (EFig. 6c-6f). These are opposite to the results of LLM’s best improvement for the general public in Study 1. Medical student data confirmed the task was not biased towards certain explanations (EFig. 4c), implying expertise drove interactions: the general public over-relies on LLMs, while PCPs are more resilient to incorrect LLM suggestions.

However, LLM explanations did help improve the alignment between PCP participants’ confidence and accuracy (correlation r=0.494, p=0.010, EFig. 3d, similar findings in top-3 performance, EFig. 5d) over No AI (correlation r=0.084, p=0.415, two-sided Fisher r-to-z test comparing the two correlations: z=2.368, p=0.018, EFig. 3d). This calibration benefit held even with incorrect AI predictions, where non-LLM explanations impaired alignment (Fisher r-to-z test z=2.572, p=0.010, EFig. 3f, similar in top-3 performance, p=0.059, EFig. 5f). PCP caution towards LLMs likely enforces cognitive engagement, thus enhancing diagnostic accuracy-confidence calibration regardless of AI correctness^54,55^.

**Fig. 5.**
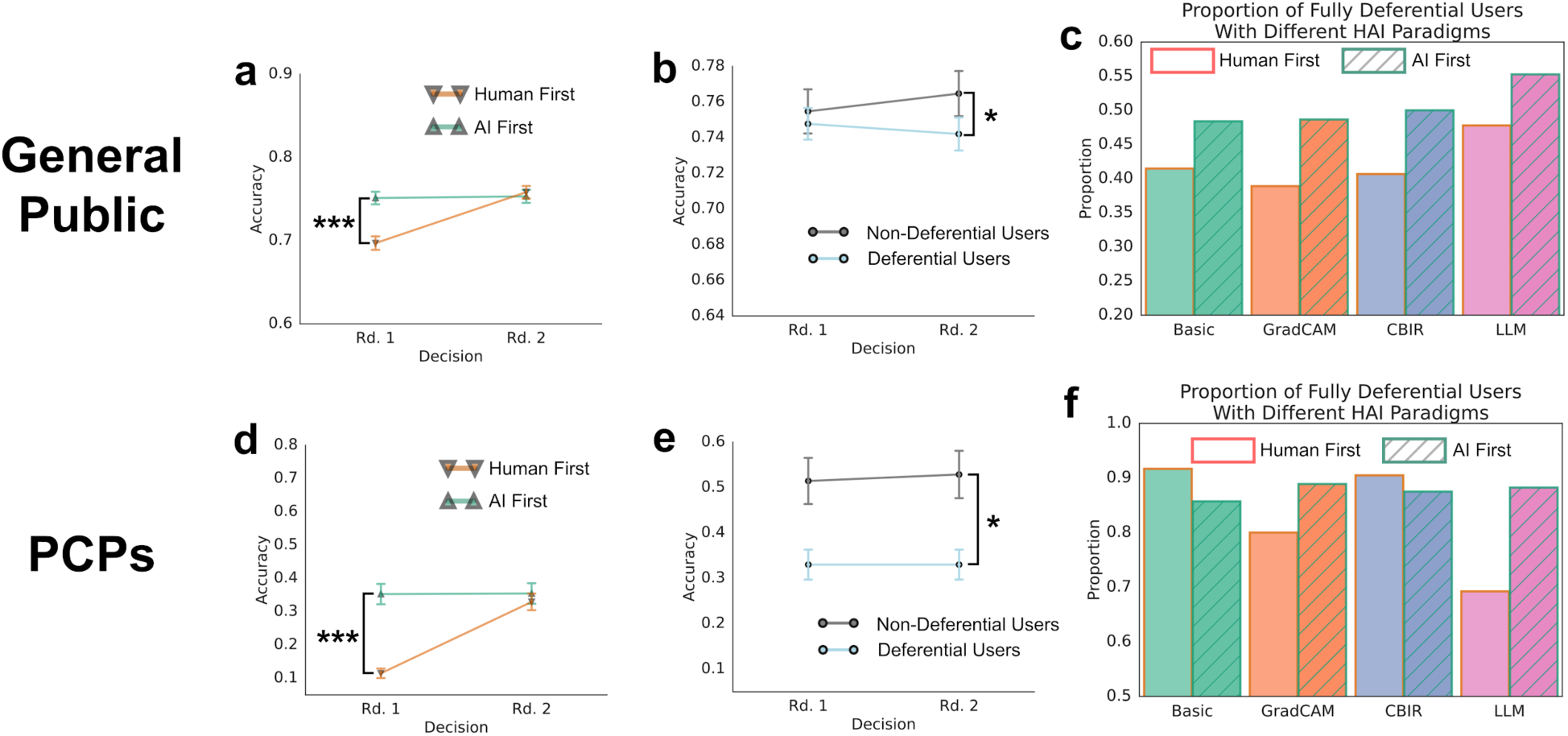
Influence of HAI Paradigm on Diagnostic Performance. **a**-**d**, Analysis for the general public. **a**, Comparison between “AI-First” and “Human-First” HAI systems on diagnostic accuracy across two rounds. **b**, Accuracy trajectories across two decision rounds for deferential and non-deferential people in the “AI-First” paradigm. **c**, Comparison of full-deference proportion between two HAI paradigms. **d**-**f**, Equivalent results for PCP participants as (**a**)-(**c**).

Our findings identify the difference between the general public and PCPs: while the general public showed vulnerability toward the influence of LLM-based explanations and exhibited higher AI deference, **PCPs improved performance with demonstrated resilience against incorrect predictions**.

### Higher AI Deference Correlates with Lower Initial Performance

We inspected the relationship between participants’ deference tendency toward AI suggestion deference and its relationship with initial performance^40^. Among participants’ final decisions that are correct (ranging from 0 to 12, 12 images total), we visualize the number of images with AI suggestions that are correct (up to 10, shown in blue) and incorrect (up to 2, shown in red) for each participant (Fig. 4a). As mentioned above, “deferential participants” are those who only got correct results when AI was correct and always got incorrect results when AI was wrong (i.e., a blue bar in Fig. 4a & 4d). Participants who got at least 1 correct outcome even when AI is incorrect are considered as “non-deferential participants” (i.e., a red bar on top of the blue bar in Fig. 4a & 4d).

While the general public had similar performance after AI assistance, deferential participants had significantly lower initial diagnostic accuracy (65.8%) compared to non-deferential participants (72.5%) before receiving AI suggestions (β=0.082, 95%CI=[0.074, 0.111], p<0.001, Cohen’s d=0.462 Fig. 4b, linear mixed model with deferential group as the main factor, controlling the same confounders as Study 1 analysis, see STab 28). Deferential PCPs also had a significantly lower performance in the initial round (β=0.194, 95%CI=[0.079, 0.310], p<0.001, Cohen’s d=1.578, Fig. 4e, STab 29), which could result from a lower level of critical thinking (p=0.028, measured by critical thinking questionnaire^49^, details see Method).

LLM explanations led to relatively the largest proportion of fully deferential participants in the general public (Fig. 4c), although no significance between LLM and others was observed (ps=0.169-0.433, STab 30). In contrast, LLM resulted in relatively the lowest proportion of deferential PCPs (Fig. 4f, ps=0.268-0.587, STab 31). This is also aligned with our findings of experts’ capability in maintaining resilience against the misdirection of wrong semantic AI explanations^12^.

### Putting AI Before Human Decisions Amplifies Deference Across Expertise Levels

In addition to XAI methods, a practical design factor for human-AI collaboration systems is the decision-making order between humans and AI, specifically either Human-First or AI-First paradigms. We found that both the general public and PCPs had significantly better performance in the first round with AI-First (ps<0.001, Fig. 5a & d), which is not surprising due to superior AI performance. In the second round, after humans received the same amount of information, there was no difference between Human-First and AI-First in either study (round 2, the general public: β=0.005, 95%CI=[−0.017, 0.026], p=0.650; PCPs: β=-0.020, 95%CI=[−0.091, 0.050], p=0.572. Linear mixed models on accuracy with human-AI collaboration paradigm, decision round, and their interaction as the main factors, controlling decision making time and other confounders. See STab 32 and STab 34 for details). This indicates that the decision order may not influence the final performance.

With the AI-First paradigm, non-deferential participants still had better performance, especially at round 2 after reviewing the examples again without AI (the general public: β=0.031, 95%CI=[0.000, 0.062], p=0.049, STab 33; PCPs: β=0.218, 95%CI=[0.009, 0.426], p=0.041, Stab 35). This is similar to the results in the Human-First paradigm in Fig. 4. In contrast, putting AI suggestions ahead increased the proportion of deferential participants in most cases (Fig. 5c, f). For the general public, the proportion was increased across all XAI methods (average Δ=+8.4%, although no significance observed after controlling all confounders, ps=0.067-0.170, Fig. 5c, STab 36). For PCPs, the largest deference increase was observed from LLM explanations (Δ=+19.0%, Fig. 5f, EFig. 5m, STab 37). This indicates that putting AI ahead may lead to stronger anchoring bias. It also suggests that although PCPs showed resistance against AI’s mislead in the Human-First paradigm, providing LLM-based explanations ahead of human choices can still cause more bias than other XAI methods and introduce risks of overreliance, even for PCPs.

### Human-AI Collaboration to Combine Each Side’s Strength

Although AI deference risks misleading participants when AI makes mistakes, deference may lead some humans to improved performance. Fig. 6 visualizes cases where either humans or AI routinely outperform each other. We found that AI tends to outperform humans in cases where the presentation of the disease is subtle, but struggles with atypical symptoms or unexpected features in the image (additional examples in EFig. 8, EFig. 9 and EFig. 10). These qualitative examples provide some initial directions for future work in understanding the complementary strengths of humans and AI in dermatological diagnosis.

**Fig. 6.**
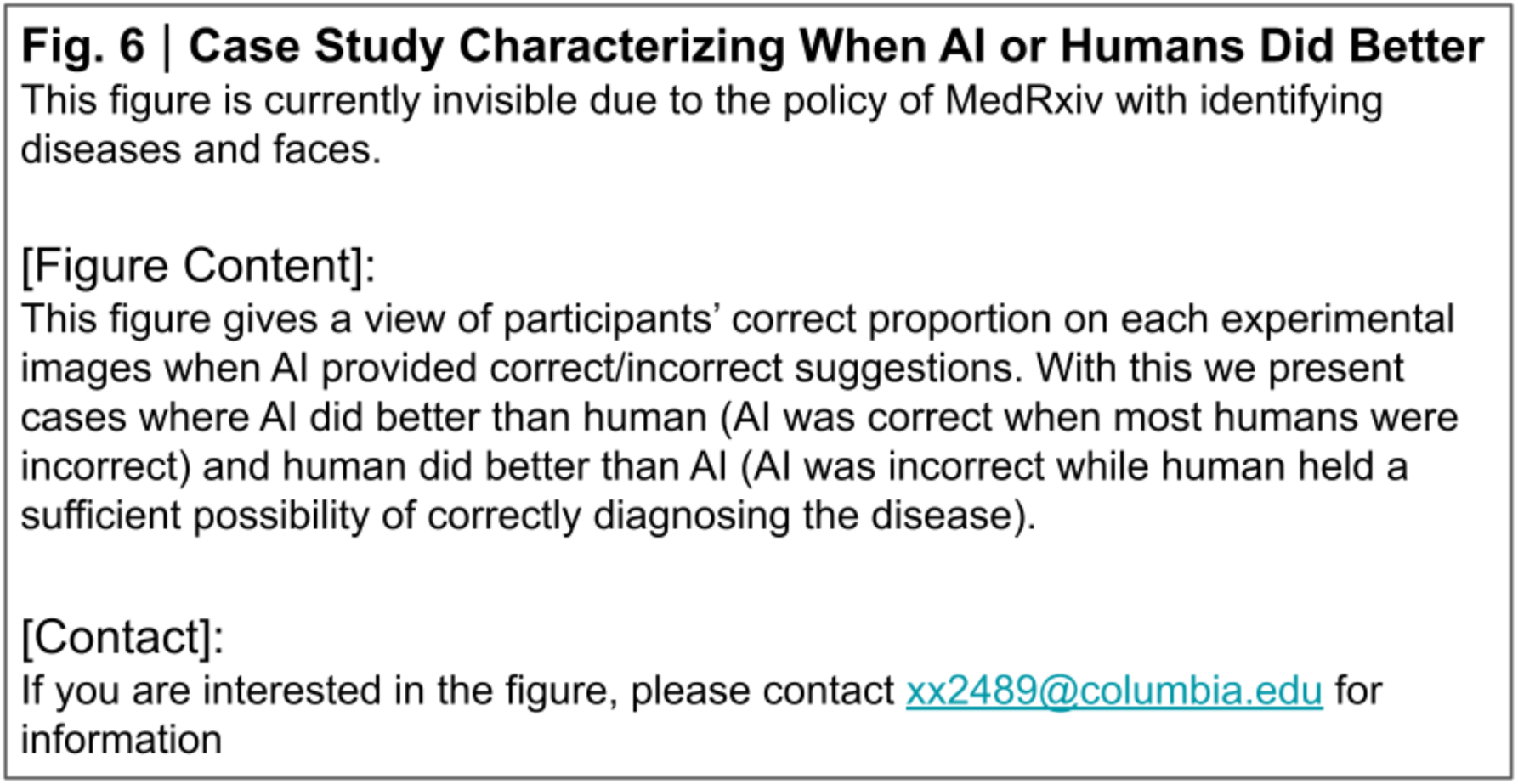
Case Study Characterizing When AI or Humans Did Better. **a**, Accuracy proportion of images in Study 1 for the general public, categorized by whether the AI prediction was correct or incorrect. The blue bars represent the proportion of correct human participants’ decisions, while the red overlays indicate their incorrect decisions. Representative images labeled “AI did Better than Human” (AI-right images with the highest human error rate) and “Human did Better than AI” (AI-wrong images with the highest human accuracy) are shown to the right. **b**, Same as (**a**) for Study 2 for PCPs. More examples of each category can be found in EFig. 8 - 10.

## Discussion

Our work contributes to the knowledge gap in understanding how various XAI methods and human-AI decision paradigms may impact human-AI collaborative diagnosis outcomes across populations with different expertise levels (the general public vs PCPs). Through two large-scale experiments, we provide comprehensive results on how humans collaborate with explainable AI assistance in skin disease diagnosis. Our studies reveal both the potential and challenges of human-AI collaboration in dermatology.

Our study demonstrated that state-of-the-art AI suggestions can create significant diagnostic improvements regardless of participants’ expertise levels and task difficulty. Congruent with previous research^30^, PCPs’ diagnostic top-1 and top-3 accuracy are increased by 21.4% and 43.5% from a baseline of 11.5% and 16.1%. The general public’s performance also increased by 6.1% from 69.7% to 75.8%. Our fairness-constrained AI model helped humans reduce diagnostic disparities on skin tones, with a relative reduction of 46.9% (accuracy disparity from 3.2% to 1.7%) among the general public and 35.6% (from 4.5% to 2.7%) among PCPs. In line with our results, Groh et al^56^ find similar diagnostic disparities across skin tones in PCPs without AI assistance and similar reductions in these disparities when PCPs had access to a similarly high accurate AI assistance tool (79.2% accuracy in this paper and 84.0% accuracy in the treatment DLS in Groh et al^56^). However, when the AI assistance tool exhibited moderate performance (the 47.0% accuracy in the control DLS in Groh et al^56^), PCP diagnostic disparities across skin tones increased. Together, these two studies reveal that the dose-response relationship between AI accuracy and its impact on diagnostic accuracy and equity deeply matters. As AI assistance accuracy decreases, physicians may be prone to incorporating incorrect AI predictions into their diagnoses especially for more challenging cases such as images of dark skin.

We found interesting nuances of different XAI methods on different populations, especially LLMs. Although as shown in Fig. 2b and Fig. 3b, a direct comparison on the final diagnosis performance across XAI methods does not reveal significance (aligned with prior findings in Chanda et al.^57^), our breakdown analysis across AI correctness and XAI quality setups provides more nuanced insights. LLM acted as a ‘double-edged sword’ for the general public. While it provided the highest accuracy improvements (+13.4%) when its suggestions were right, it also led to the largest performance decrease (−21.1%) when wrong. The general public also tended to trust LLM explanations regardless of their quality, suggesting that readable, semantic explanations are more convincing for those without medical backgrounds. This heightened deference to LLMs likely stems from the cognitive ease of narrative persuasion^58,59^. Unlike GradCAM, which requires a user to interpret an abstract heatmap, or CBIR, which demands a high-effort visual comparison, an LLM provides a cohesive and seemingly complete diagnostic story. It connects visual features to a conclusion with authoritative, human-like prose (e.g., “The lesion is suspicious *because* of its irregular border…”). This fluent narrative can create an illusion of understanding, making the reasoning feel more intuitive and plausible to a non-expert. Moreover, when the LLM highlights specific features like “irregular border,” it can anchor the user’s perception, causing them to reinterpret the ambiguous visual data in a way that confirms the AI’s plausible-sounding rationale. Our work extends prior research on non-expert overreliance and automation bias on AI^60–62^ and finds greater risk of LLM-based explanations, which has been supported in recent human-LLM trust work outside healthcare^63^. With numerous AI-assisted medical decision support tools available to the general public^64,65^, and the advancement of LLM in health AI research^66–68^, our findings highlight the importance of strong scrutiny and careful design for AI safety.

In contrast, PCPs applied LLM assistance more carefully and maintained their diagnostic accuracy even when AI suggestions were wrong. This was true regardless of explanation quality. To isolate the impact of task difficulty levels, we compared medical students to PCPs and found that students are more easily misled by LLM than PCPs. This suggests that PCPs expertise could be a mediator to help participants override or defer incorrect AI suggestions^69–71^. Although this is in contrast to prior findings that medical experts (non-dermatology) are also misled by AI outcomes^40,70^, our results are aligned with prior studies in dermatology^12^ that experts are better at adopting AI than non-experts. Our further investigation revealed that AI deference is less common when medical student and the PCP groups were equipped with more medical expertist. This indicates that medical knowledge and experience could be one of the key reasons for appropriate AI adoption for medical experts. Compared to the general public, experts’ medical expertise acts as a crucial *cognitive firewall*^40,72^. While a layperson uses the explanation to *form* a belief, a PCP uses it to *validate* an existing hypothesis against their own structured clinical knowledge.

### Design Implications

These findings have important design implications for how to implement AI suggestions for both the general public and PCPs. First, our work strongly suggests that a “one-size-fits-all” approach to medical XAI is not only suboptimal but potentially hazardous. Instead, systems must be carefully tailored to the user’s expertise and the specific collaborative goal. For the general public, where the risk of over-reliance on persuasive LLM narratives is high, designs should prioritize safety over persuasiveness. This may involve explicitly showing the AI’s uncertainty and fallibility on a case-by-case basis to calibrate user trust, framing suggestions not as diagnoses but as preliminary information that requires professional validation, or favoring less-narrative XAI modalities like content-based image retrieval (CBIR), which encourage active comparison rather than passive acceptance. This is also supported by the recent call for medical safety disclaimer in generative AI model outcomes^35^. For clinicians, the goal shifts from simple guidance to expert augmentation. Systems should support adaptive explanations, potentially offering a concise rationale for routine cases but revealing more in-depth evidence—such as similar cases (CBIR) or conflicting features—when the clinician’s initial diagnosis differs from the AI’s suggestion.

Furthermore, the structure of the interaction itself is a powerful design tool for mitigating cognitive bias. Our finding shows AI suggestions first leads to stronger anchoring bias^73^ and higher deference, which enriches the current understanding of the effect of human-AI decision orders.^41,74^ This is clear evidence for a “Human-First” workflow, where the clinician formulates and commits to an initial diagnosis before being exposed to the AI’s input, to preserve independent human reasoning and minimize the powerful anchoring bias that an upfront AI suggestion can create.

Finally, we need to design systems that account for inevitable disagreements (EFig. 8-9) and individual differences in AI deference. Our finding that participants prone to AI deference often have lower baseline performance reveals a crucial challenge: the users most in need of help are also the most vulnerable to being misled. This extends recent work^40^ on AI-deference among humans and points toward the future of an adaptive system. Instead of a static information handoff, the system should dynamically tailor their interaction style to individual users. A highly deferential user might require iterative explanation^75^ or explicit prompts for the user’s confidence before finalizing a diagnosis^76^, or automatically trigger a workflow for a second human opinion when the AI’s suggestion significantly alters an initial assessment^77^. Designing these feedback loops is essential to better align AI reliance with actual user skill and minimizing the risks of misjudgment in high-stakes medical contexts.

### Limitation

Our work has several key limitations. First, although we design our study to be close to real-world scenarios, the ecological validity of the study is still limited. Only a small subset of the datasets we adopted contain patients’ demographic information. Participants did not have access to contextual data (e.g., age, gender, socioeconomic status), which are crucial factors in real-world diagnosis. The task designs were also simplified: for the general public, the binary melanoma detection task may not reflect how laypersons typically judge whether a skin lesion is concerning and requires medical attention. For PCPs, making a diagnosis based solely on an image is a significant departure from standard clinical practice, which integrates physical exams and patient history. Second, our online studies only include 12 images per participant. This relatively small number may be insufficient to fully capture enough data to analyze individual patterns, potentially limiting the generalizability of our findings. Third, our results are specific to the AI models and XAI methods we implemented. The characteristics of the post-hoc explanations are inherently tied to the underlying model’s training algorithm (i.e., our model necessarily shapes the internal representations from which explanations are derived). This is a potential confounding factor common in studies of post-hoc XAI. Similarly, the stochasticity in LLM explanation generation, together with the specific prompt in STab 3 that may elevate a confident tone, introduces a confounder that is difficult to control. In our current studies, LLM explanations were generated in a single pass without selection, refinement or post-hoc optimization. Besides, the observed effects might not generalize to different AI architectures, alternative explanation techniques, or future, more advanced models^78,79^. Future work should aim to address these limitations by incorporating richer patient context in more authentic clinical workflows and evaluating a larger number of cases.

## Conclusion

In conclusion, our work shows both the potential and challenges of using explainable AI, especially LLMs, in skin disease diagnosis. AI collaboration can improve diagnostic accuracy and fairness, but the significant differences in how PCPs and the general public use AI assistance demonstrate the need for carefully designed systems for different users. Our research also opens several directions for future work. Our findings should be replicated in other medical domains, and verified with additional patient information, e.g., medical history and environment, in AI systems. We must also target more work into making explanations helpful for different users, while encouraging critical thinking.

## Methods

### Image Data Collection

We collected a set of diverse datasets of clinical images spanning multiple skin tones for both studies with the general public and experts. Specifically, we combined 16 public and private datasets (see STab. 4 for details of each dataset), resulting in a total of 108,585 images after removing duplicates. These images contain human expert annotated ground truth labels and cover a wide range of skin conditions, including nevus (N=24,746, 22.8%), melanoma (N=8,686, 8.0%), atopic dermatitis (N=1,912, 1.8%), pityriasis rosea (N=609, 0.6%), Lyme disease (N=248, 0.2%), CTCL (N=330, 0.3%), and other skin diseases. In the Study 2 differential diagnosis task, we mainly focused on four diseases – atopic dermatitis, pityriasis rosea, Lyme disease, CTCL – as these four were found to have the greatest performance disparities across skin tones in a recent study^30^. 18,265 images (16.8%) also contain expert-annotated Fitzpatrick labels (Fitzpatrick labels 1-4: 15,845 out of 18,265 images, 86.8%; Fitzpatrick labels 5-6: 2,420 images, 13.2%). For the rest, we used individual typology angle (ITA) calculated from the segmented skin pixels with the YCbCr algorithm^45,80^. Using the thresholds from Kinyanjui et al., 2020^81^, we assign Fitzpatrick labels to these images. Note that these images with weak Fitzpatrick labels were only used for fair model training. For the actual user study, we randomly sampled 82 high-quality, expert-annotated images from the image pool for Study 1 with the general public (equally distributed between nevus and melanoma, and between Fitzpatrick labels 1-4/5-6), and 182 images for Study 2 with medical experts (82 atopic dermatitis, 54 pityriasis rosea, 27 Lyme disease, 27 CTCL, and 92 other diseases, equally distributed between skin tones within each category). The sample sizes balanced the variance of the images and adequate number of decisions per image.

### AI Models and Explanations

We trained fair AI models for skin disease diagnosis. For Study 1, we trained a binary classification model (nevus vs melanoma). For Study 2, we implemented a hierarchical structured multi-class model that distinguished a total of 34 most common skin diseases (see the total list in STab. 4). We trained two models, including a primary 5-class classification model (four main diseases and an “other” class) and a secondary 30-class classification model (30 other common diseases). The secondary model was only used when the first model predicted “other”.

All three models were trained following the protocol in Yang et al^82^. to achieve fair medical AI models. Specifically, for each task, we train a set of models using five well-established fairness algorithms, including empirical risk minimization (ERM)^83^, Domain-Adversarial Neural Networks (DANN)^84^, Conditional DANN (CDANN)^85^, group distributionally robust optimization (GroupDRO)^86^, and exponential moving average (MA)^87^. We performed a grid search on three pretrained models (DenseNet-121^53^ pretrained on ImageNet 1k^88^, ViT-B/32^51^ pretrained on ImageNet 1k, and ViT-B/32 pretrained on ImageNet 21k^89^) and model hyperparameters via a random search of 25 trials. Each model was finetuned for up to 30,000 steps with a batch size of 64 using the Adam optimizer, with checkpointing every 1,000 steps, and early stopping if the overall validation macro AUROC does not improve for 5 checkpoints. For each algorithm, we first selected the architecture and hyperparameter combination that maximizes the worst-group validation AUROC. We then selected algorithms based on the tradeoff between overall accuracy and fairness. Our final binary classification model for Study 1 (CDANN, ViT-B/32 pretrained on ImageNet 21k, λ=0.218, learning rate=0.0005) achieved an overall weighted AUROC of 0.930 (light skin 0.933, dark skin 0.898, Δ=0.035) a weighted balanced accuracy of 0.850 (light skin 0.852, dark skin 0.831, Δ=0.021). Compared to the baseline method ERM (balanced accuracy 0.845, Δ=0.091), our model reduced the performance gap by 76.9%. For Study 2, the final primary model (CDANN, DenseNet-121 pretrained on ImageNet 1k, λ=8.388, learning rate = 0.0029) achieved an overall AUROC of 0.772 (light skin 0.782, dark skin 0.691, Δ=0.091), weighted AUROC of 0.753 (light skin 0.725, dark skin 0.693, Δ=0.192. Note that it is possible for a total AUROC falls out of range of its breakdown), and a balanced accuracy of 0.478 for the four main diseases (light skin 0.487, dark skin 0.431, Δ=0.056), consistently outperforming the baseline ERM (balanced accuracy 0.457, Δ=0.144). The final secondary model (CDANN, ViT-B/32 pretrained on ImageNet 1k, λ = 0.377, learning rate = 0.0003) achieved an overall a AUROC of 0.728 (light skin 0.714, dark skin 0.641, Δ=0.073), a weighted AUROC of 0.752 (light skin 0.798, dark skin 0.744, Δ=0.053) a balanced accuracy of 0.141 (light skin 0.157, dark skin 0.092, Δ=0.064). STab. 5 and STab. 6 list out the specific performance details of each disease. Finally, we intentionally sampled images from our testing set with high quality disease and skin tone labels (i.e., diagnosis verified through human experts such as biopsy, and image resolution is higher than 400×400 pixels) to achieve the expected accuracy and fair results presented to participants, as described in the Results section.

Building upon the model, we then generate three types of post-hoc explanations (GradCAM, CBIR, and multimodal LLM). For GradCAM, we adopted the algorithm in Selvaraju et al. and picked the last layer in the model to generate a heatmap and overlaid it on the original image. For CBIR, we computed the cosine distance of the embedding of the target image against the embeddings of images in the training set, and selected the top-2 images with the same skin tone and top-1 image with the opposite skin tone, leading to three retrieved images for each target image. As for multimodal LLM-based explanations, we used multimodal GPT-4V (a vision-language model) and constructed the prompt to generate explanations for the target reader (the general public or medical experts, see more prompt details in STab. 3). Note that the diagnoses were determined by our own model (including cases when predictions were wrong) and sent to GPT-4V using the OpenAI API, and GPT-4V was only prompted to generate text-based explanations rather than perform diagnosis.

### Ethics approval for Human Subject Studies

This research complies with all relevant ethical regulations. The Massachusetts Institute of Technology’s Committee on the Use of Humans as Experimental Subjects approved this study as Exempt Category 3—Benign Behavioral Intervention. Participants were informed about the potential sensitive visual content and provided consent before joining the study. They had the option to leave the experiment at any time. Images used in the study were publicly released by prior work.

### Experimental Interface

We developed a reactive experiment interface based on Qualtrics for online digital experiments. For Study 1, the experiment was designed to mimic real-life scenarios where users can self-diagnose using a photo captured by smartphones/webcams. Prior to the study, the general public participants were provided with educational content and common recognition guidelines about melanoma (ABCDE rules^90^, see SFig. 1), together with practice questions (SFig. 2) and experiment instructions of AI and its explanations (SFig. 3). They then went through 12 images and made a diagnosis (distinguishing melanoma vs. nevus, as well as their confidence of the decision). Participants could hover on each image (including GradCAM or CBIR images) to use a magnifier for closer inspection. The interface design of Study 2 was similar to Study 1. Expert participants were asked to do open-ended differential diagnosis as free-text entry tasks, in which they were asked to enter top-3 diagnosis, confidence, and whether they had opinions on referral (including no referral, a dermatologist, biopsy, and both). Each text entry box had auto-completion based on the matched string they entered, and the corpus included 445 (see SFig. 5a). These setups were designed to resemble real-world differential diagnosis scenarios, such as telehealth, or patients sending images through electronic health record messaging systems. They received task-specific instructions with examples (see SFig. 4) and practice cases (see SFig. 5), before going through 12 images.

### Human Subject Study Design and Protocol

We recruited general public participants from Prolific (https://www.prolific.com/) and Facebook Groups for Study 1 (March 20 - May 1, 2024). In Study 2 (June 10 - Sept 3, 2024), we leverage several clinical networks and platforms to recruit medical experts, including local networks at the Boston Medical Center and Stanford University, Centaur Lab (https://www.centaurlabs.com/), MedShr (https://en.medshr.net/), Pathway (https://www.pathway.md/), and XPC (https://www.xprimarycare.com/). For both studies, we embedded attention check questions in the middle of the study (see an example in SFig. 5c.) for quality control. We also filtered answers with abnormal durations (i.e., less than 10 seconds or more than 5 minutes per image on average). After filtering, we collected 623 lay people from the general public for Study 1 and 153 PCPs for Study 2 (320 medical students in addition).

We adopted a between-subjects design. Participants were randomly assigned to one of the eight experimental conditions varying in the four XAI types (Basic confidence explanations, GradCAM, CBIR, or LLM) and two human-AI decision paradigms (Human-First vs AI-First). In both studies, each participant evaluated 12 images and provided two rounds of diagnoses per image. In the Human-First paradigm, participants made unassisted initial diagnoses and reported their confidence before receiving AI input at Round 1. They then reviewed the AI suggestions and explanations (depending on their experiment group) and reported Round 2 decision and confidence. In the AI-First paradigm, AI suggestions and explanations were presented alongside the initial image at Round 1. In Round 2, AI outcomes were hidden and participants were asked to review their decisions again. By introducing Round 2 in the AI-First condition, we controlled the number of decisions and cognitive engagement.

### Assessment of Individual Characteristics

In addition to the diagnostic tasks of 12 images, participants completed a series of questionnaires, including demographics and established assessments to measure human-AI collaboration experience^47^ and AI explanation quality^48^ (STable. 2). For medical experts, we further collected additional cognitive traits, including open-mindness^50^, critical thinking^49^ (STab. 1). Although it is not the main focus of our work, EFig. 4 presents the relationship between these subjective aspects and the final decision performance.

## Data Availability

For ML model training, our datasets are organized and available in this link. It’s description is available in the README file in the Figshare subfolder of ml-model (https://figshare.com/s/eaf2110da509abbe9a6a). For the two human-subject studies, the images used in the human experiment and the study results are organized and available in the Figshare subfolder of human-study-results-analysis (https://figshare.com/s/eaf2110da509abbe9a6a). We will make these data available to any researchers who agree to only use this dataset for scientific and medical purposes and share a public link upon acceptance.

## Code Availability

The data and code are organized together. For ML model training, our model training code, and explanation generation code are organized and available in the uploaded Figshare subfolder of ml-model (https://figshare.com/s/eaf2110da509abbe9a6a). For the two human-subject studies, the code for data analysis visualization are organized and available in the Figshare subfolder of human-study-results-analysis (https://figshare.com/s/eaf2110da509abbe9a6a). We will host and share a public link upon acceptance.

## Extended Data Figs & Tables

**EFig. 1.**
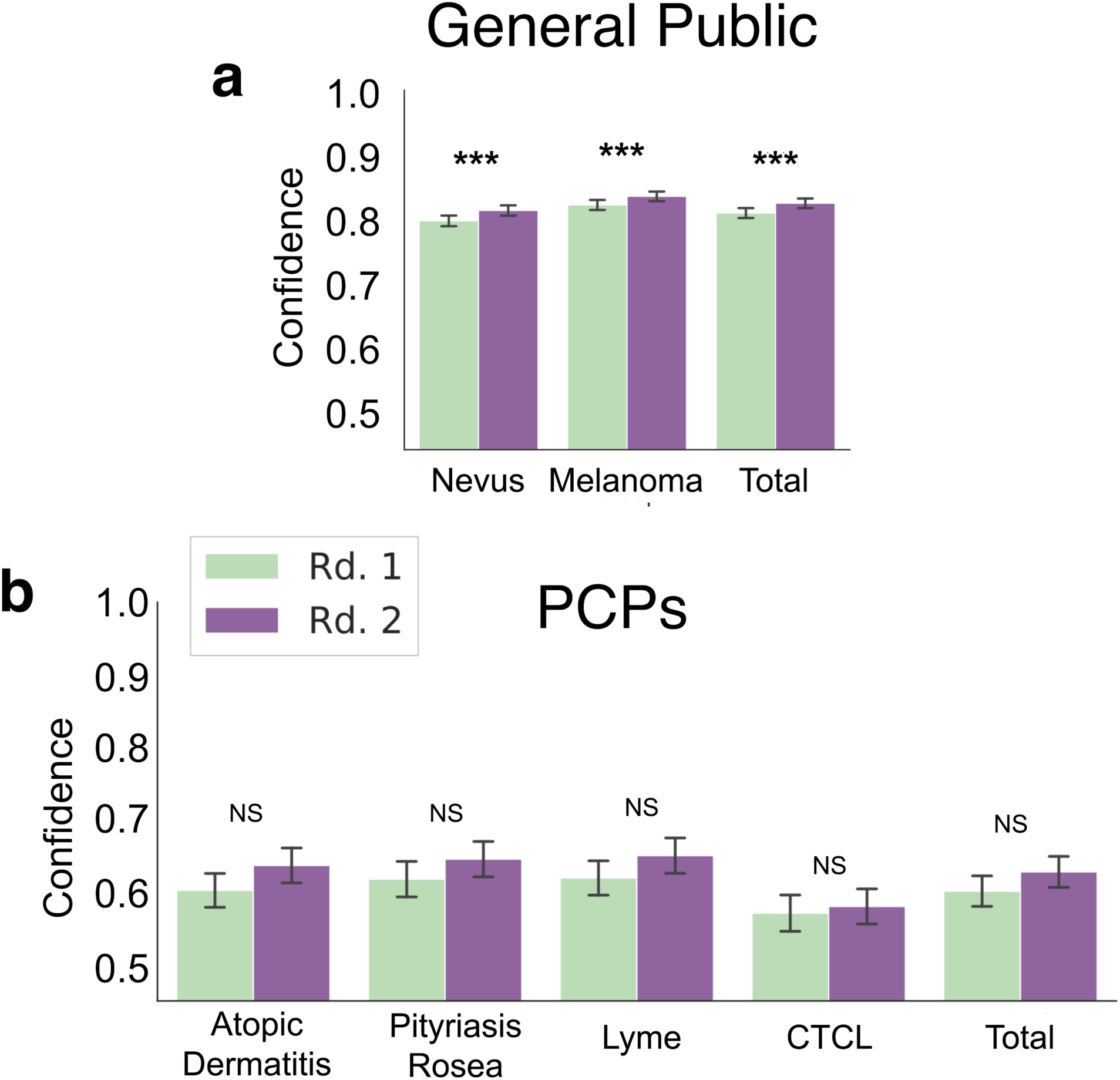
Diagnostic Confidence of the General Public and PCPs. **a**, Confidence levels of the general public in detecting nevus, melanoma, and the overall diagnosis in two rounds: Rd. 1 (initial, human accuracy without AI assistance) and Rd. 2 (AI-assisted accuracy). **b**, Confidence levels of PCPs in identifying atopic dermatitis, pityriasis rosea, Lyme disease, CTCL, and the overall performance on four main diseases, with results presented for two rounds.

**EFig 2.**
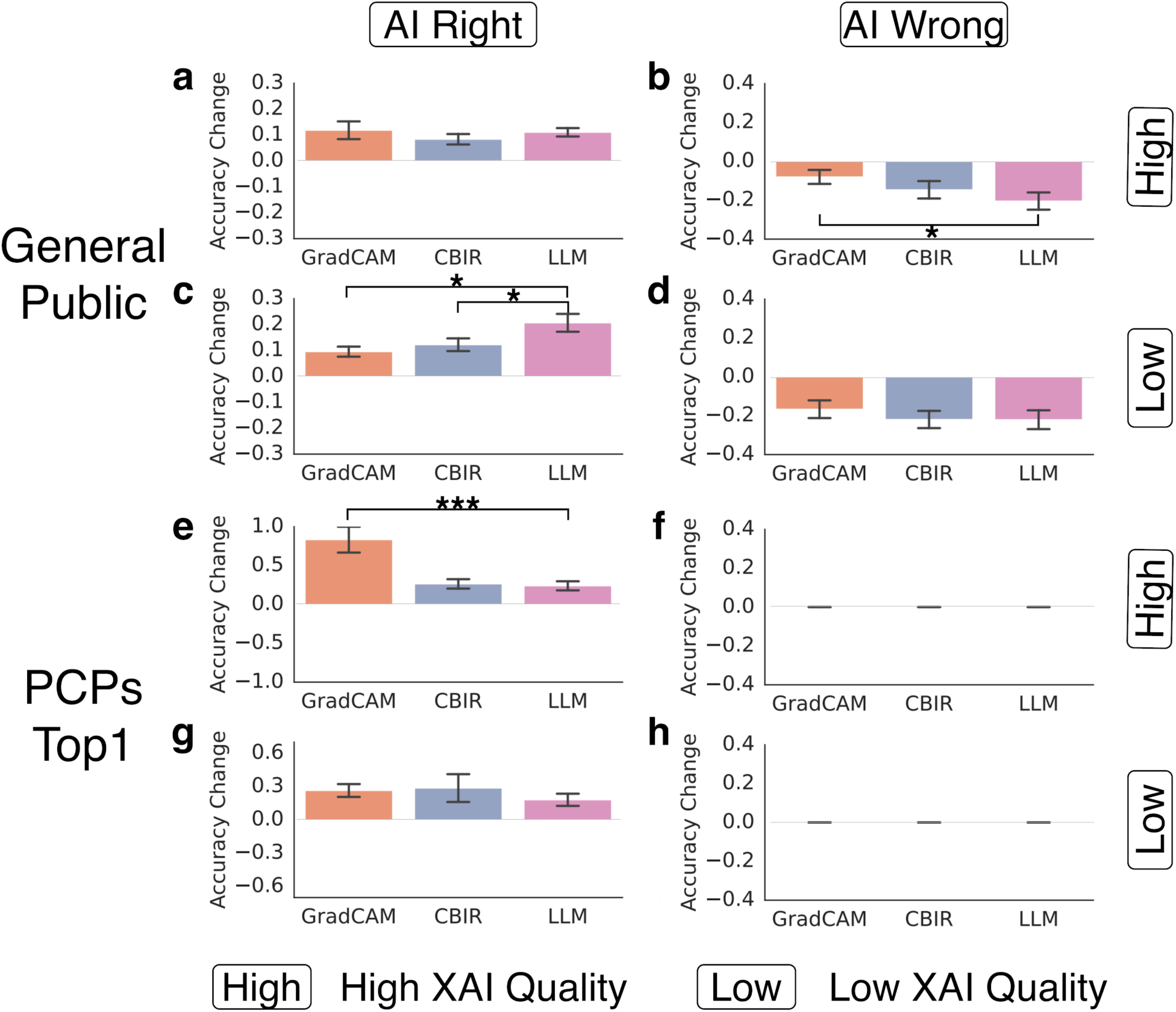
Diagnostic Accuracy Under Different XAI Qualities. **a**, **b**, Changes in diagnostic accuracy for the general public under high XAI quality, comparing GradCAM, CBIR, and LLM explanations when AI is right (**a**) and AI is wrong (**b**). **c**, **d**, Visualization of diagnostic accuracy changes for the general public under low XAI quality conditions, highlighting the effects of right AI suggestions (**c**) and wrong suggestions (**d**) for each XAI approach. **e**-**h**, Equivalent results as (**a**)-(**d**), shown for top1 diagnostic accuracy of PCP participants.

**EFig. 3.**
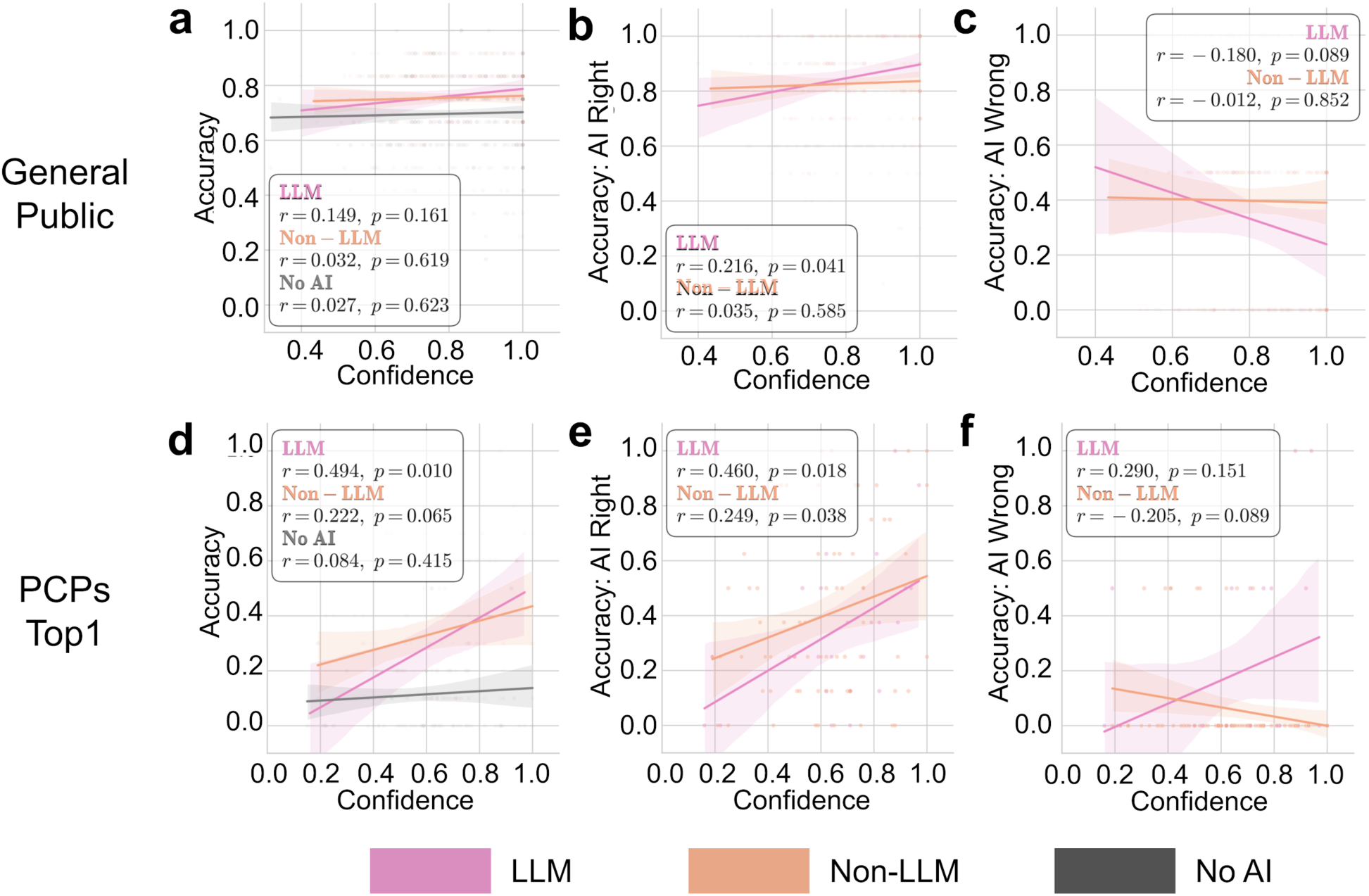
Accuracy-Confidence Consistency of the General Public and Top1 Diagnosis of PCPs. **a**-**c**, General public’s performance comparison between LLM and non-LLM assisted systems, in (a) overall diagnostic accuracy-confidence consistency (A-C consistency), (**b**) A-C consistency with correct AI predictions, and (**c**) A-C consistency with wrong AI predictions. **d**-**f**, Equivalent results for top1 diagnosis of PCP participants as (**a**)-(**c**). Statistical significance was assessed using Pearson correlation coefficients (r) with associated p-values. Shaded areas represent 95% confidence intervals. Same metrics are used in EFig. 5.

**EFig. 4.**
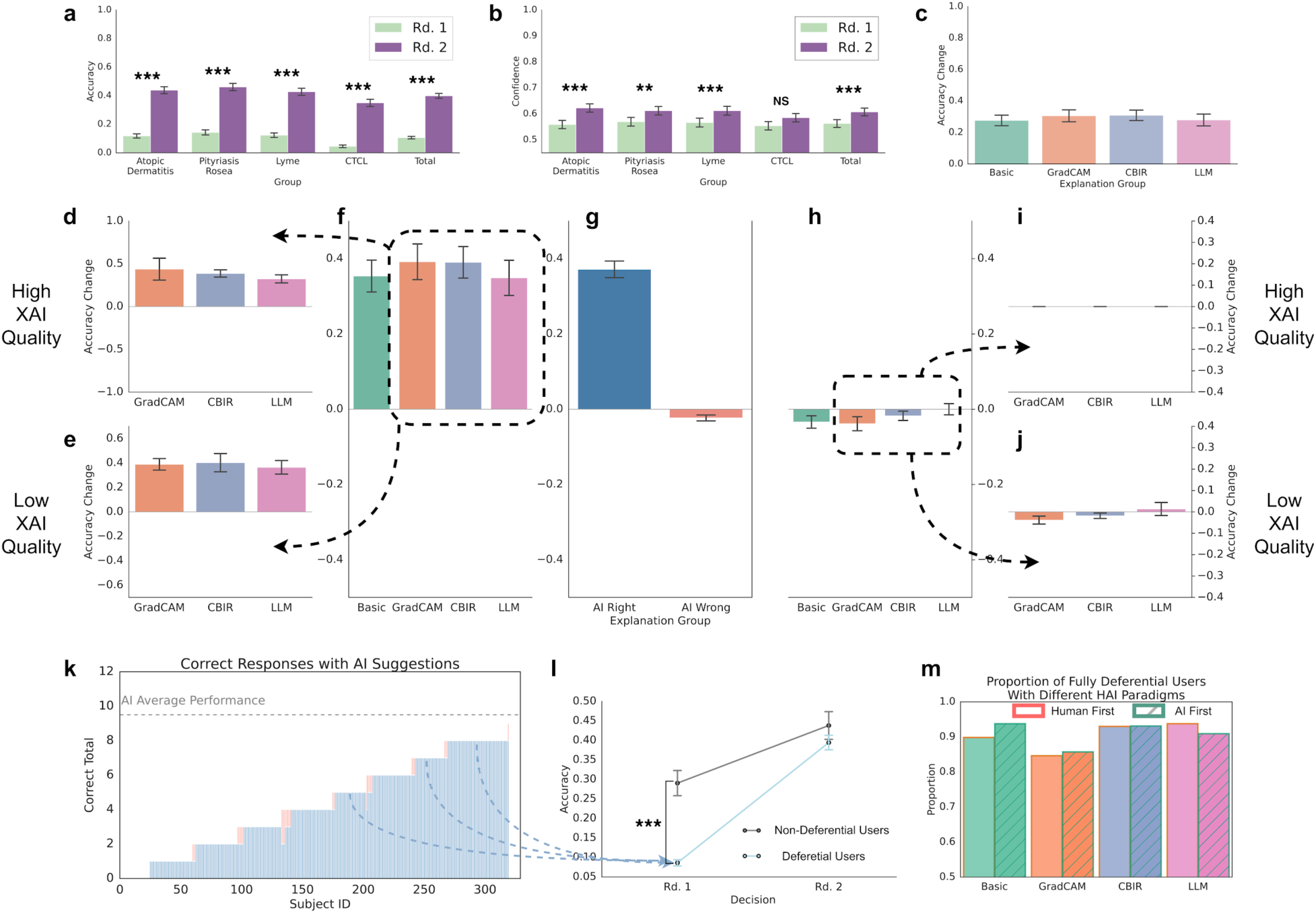
Medical Student in Human-AI Collaboration. **a**, Accuracy in identifying atopic dermatitis, pityriasis rosea, Lyme disease, CTCL, and the overall performance on four main diseases, with results presented for two rounds. **b**, Confidence levels of participants in their diagnostic decisions for tasks in (**a**). **c**, Diagnostic accuracy improvement across different XAI. **d, e,** Changes in diagnostic accuracy under good XAI quality (**d**) and bad XAI quality (**e**) when AI is right, comparing GradCAM, CBIR, and LLM explanations. **f, h** Diagnostic accuracy changes under AI-right (**f**) and AI-wrong (**h**) predictions for different XAI approaches. **g,** Overall accuracy changes under AI-right and AI-wrong conditions, integrating the effects of explanation quality and correctness. **i, j,** Visualization of diagnostic accuracy changes under AI-wrong conditions, highlighting the effects of good XAI quality (**i**) and bad XAI quality (**j**). **k**-**l**, Performance comparison between deferential and non-deferential participants. (**k**) Distribution of correct responses with AI assistance, (**l**) accuracy trajectories across two decision rounds. **m**, Proportion of fully deferential participants across different XAI approaches and HAI paradigms.

**EFig. 5.**
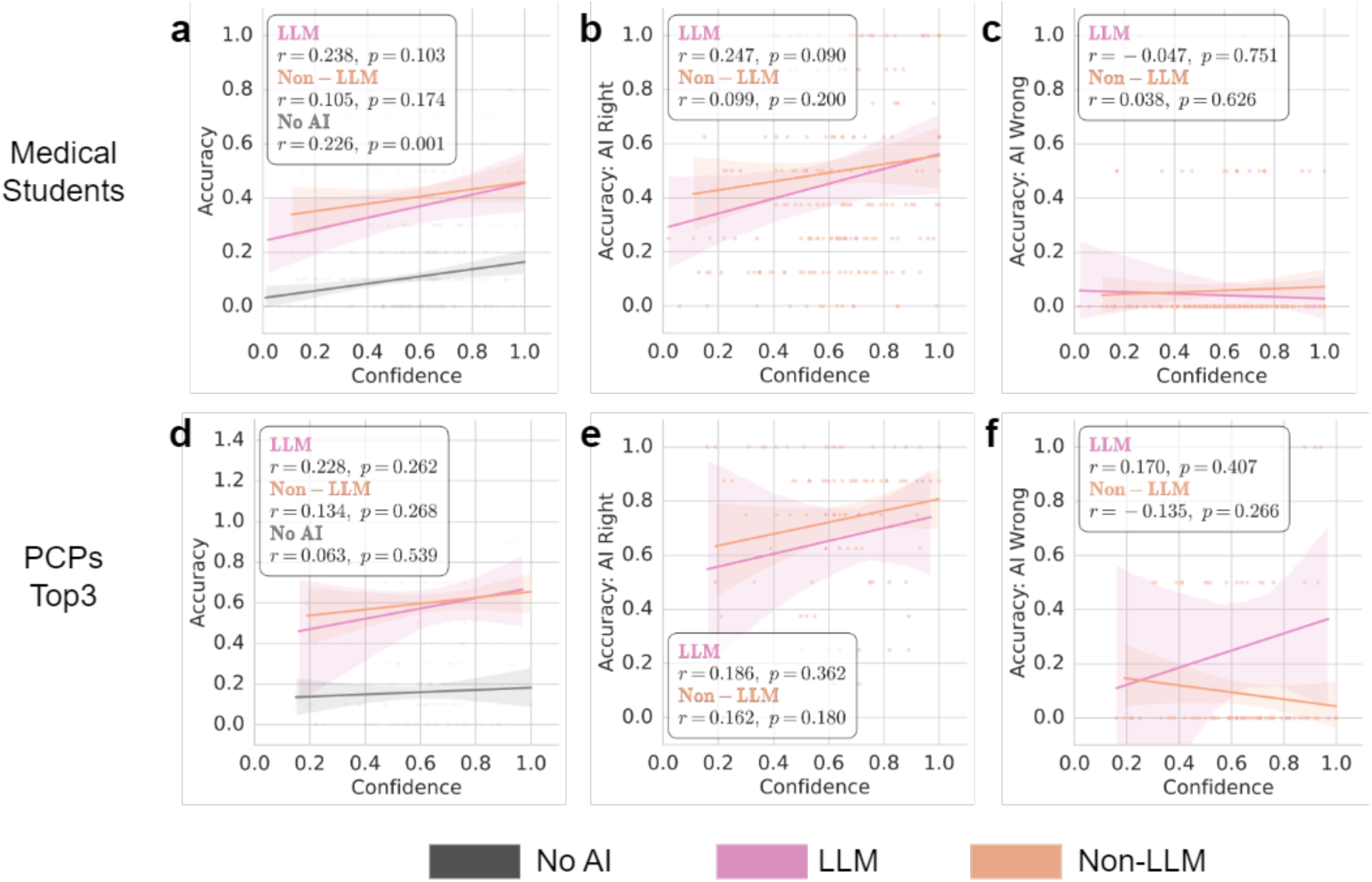
Accuracy-Confidence Consistency of Medical Students and Top3 Diagnosis of PCPs. **a**-**c**, Medical students’ performance comparison between LLM and non-LLM assisted systems, in (a) overall diagnostic accuracy-confidence consistency (A-C consistency), (**b**) A-C consistency with correct AI predictions, and (**c**) A-C consistency with wrong AI predictions. **d**-**f**, Equivalent results for top3 diagnosis of PCP participants as (**a**)-(**c**).

**EFig. 6.**
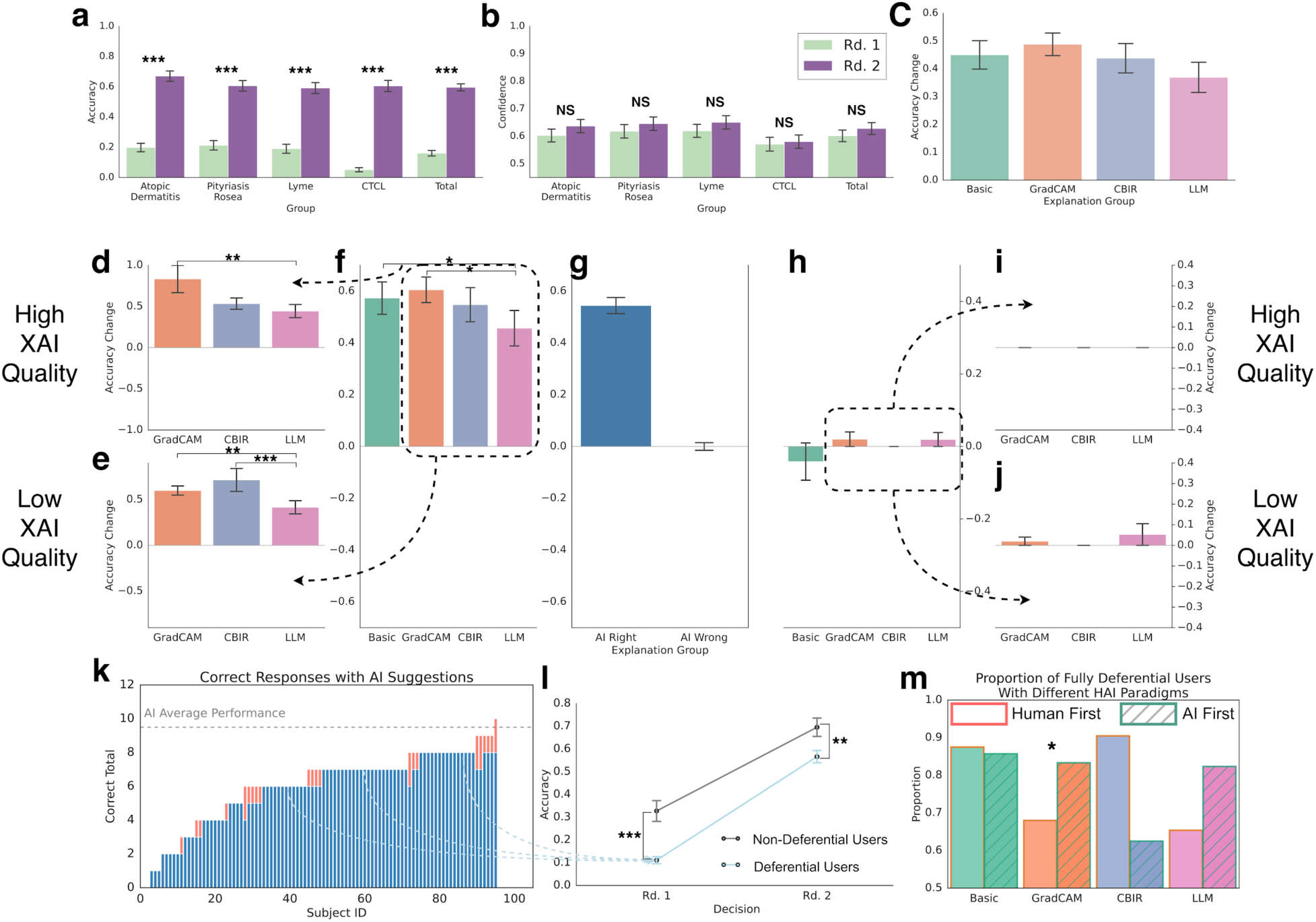
PCP Performance - Top3 Accuracy. **a**, Accuracy in identifying atopic dermatitis, pityriasis rosea, Lyme disease, CTCL, and the overall performance on four main diseases, with results presented for two rounds. **b**, Confidence levels of participants in their diagnostic decisions for tasks in (**a**). **c**, Diagnostic accuracy improvement across different XAI. **d, e,** Changes in diagnostic accuracy under good XAI quality (**d**) and bad XAI quality (**e**) when AI is right, comparing GradCAM, CBIR, and LLM explanations. **f, h** Diagnostic accuracy changes under AI-right (**f**) and AI-wrong (**h**) predictions for different XAI approaches. **g,** Overall accuracy changes under AI-right and AI-wrong conditions, integrating the effects of explanation quality and correctness. **i, j,** Visualization of diagnostic accuracy changes under AI-wrong conditions, highlighting the effects of good XAI quality (**i**) and bad XAI quality (**j**). **k**-**l**, Performance comparison between deferential and non-deferential participants. (**k**) Distribution of correct responses with AI assistance, (**l**) accuracy trajectories across two decision rounds. **m**, Proportion of fully deferential participants across different XAI approaches and HAI paradigms.

**EFig. 7.**
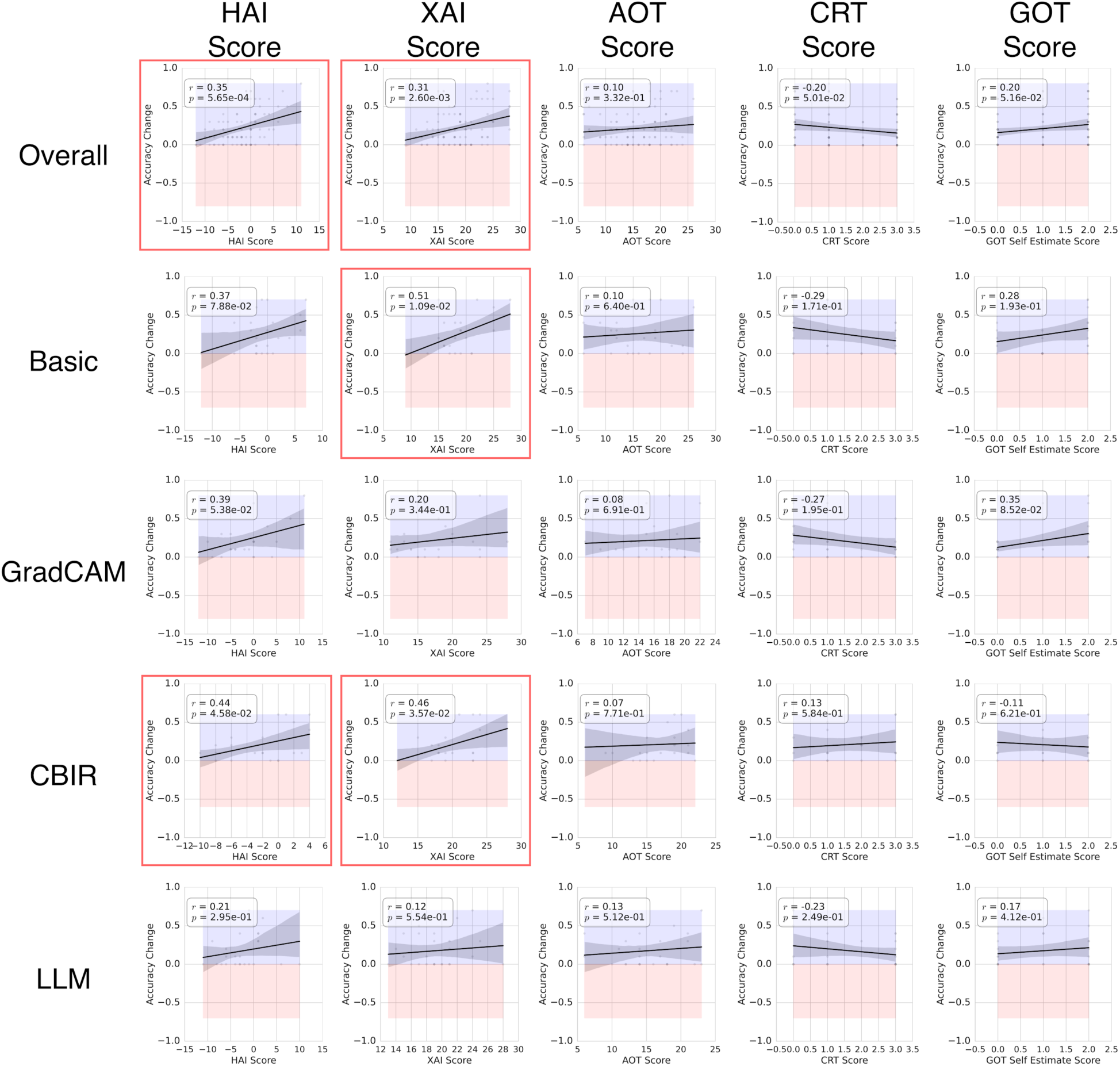
Correlation Between Accuracy Change with AI Assistance And Personality Test Score. In each subfigure, the blue band (maximum value) and red band (minimum value) indicate the range of accuracy change. The red border around the subfigure represents a significant correlation found (p<0.05).

**EFig. 8.**
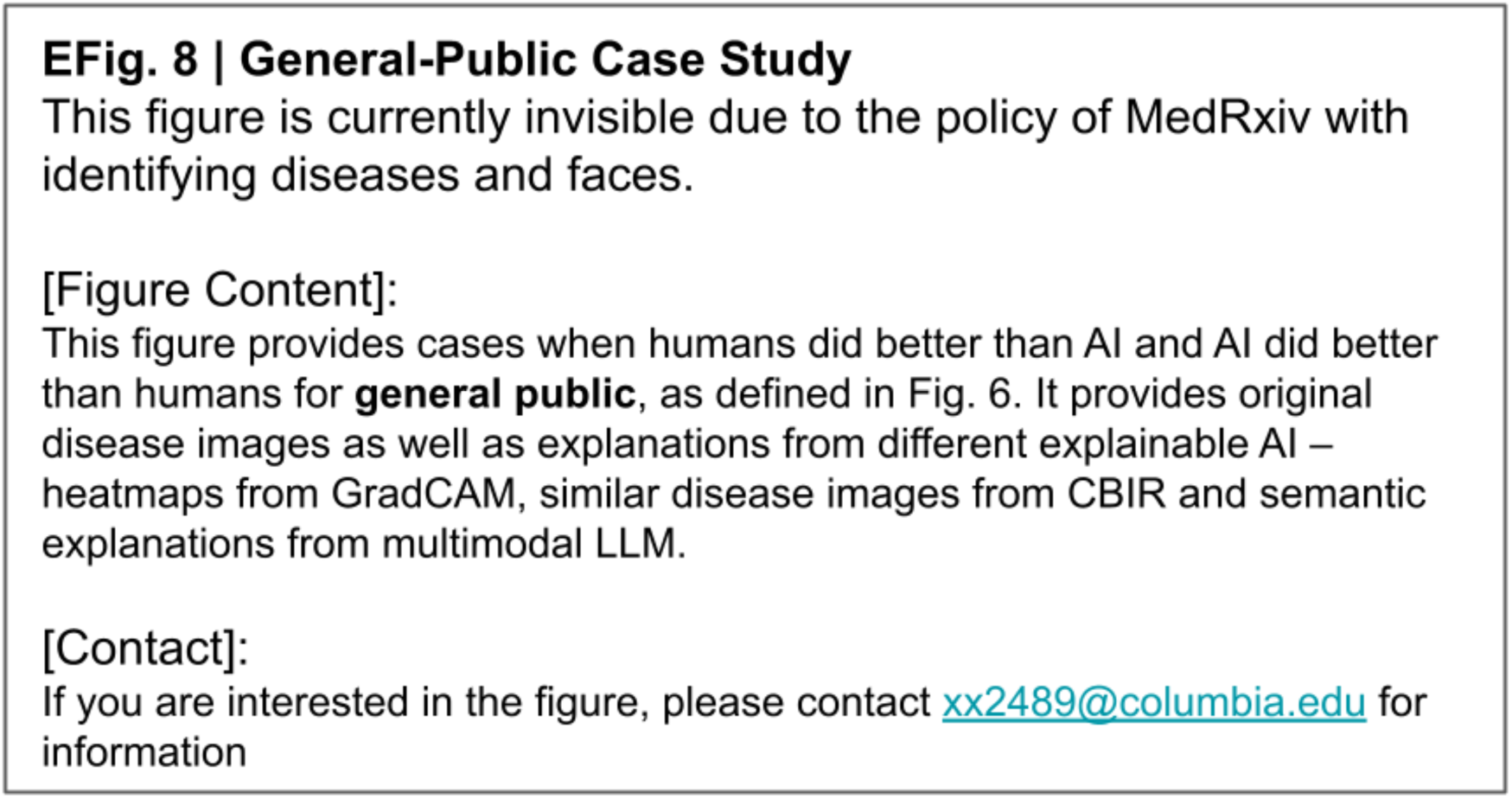
General-Public Case Study. **a,** AI did a better diagnostic performance than humans**. b,** Humans did a better performance than AI.

**EFig. 9.**
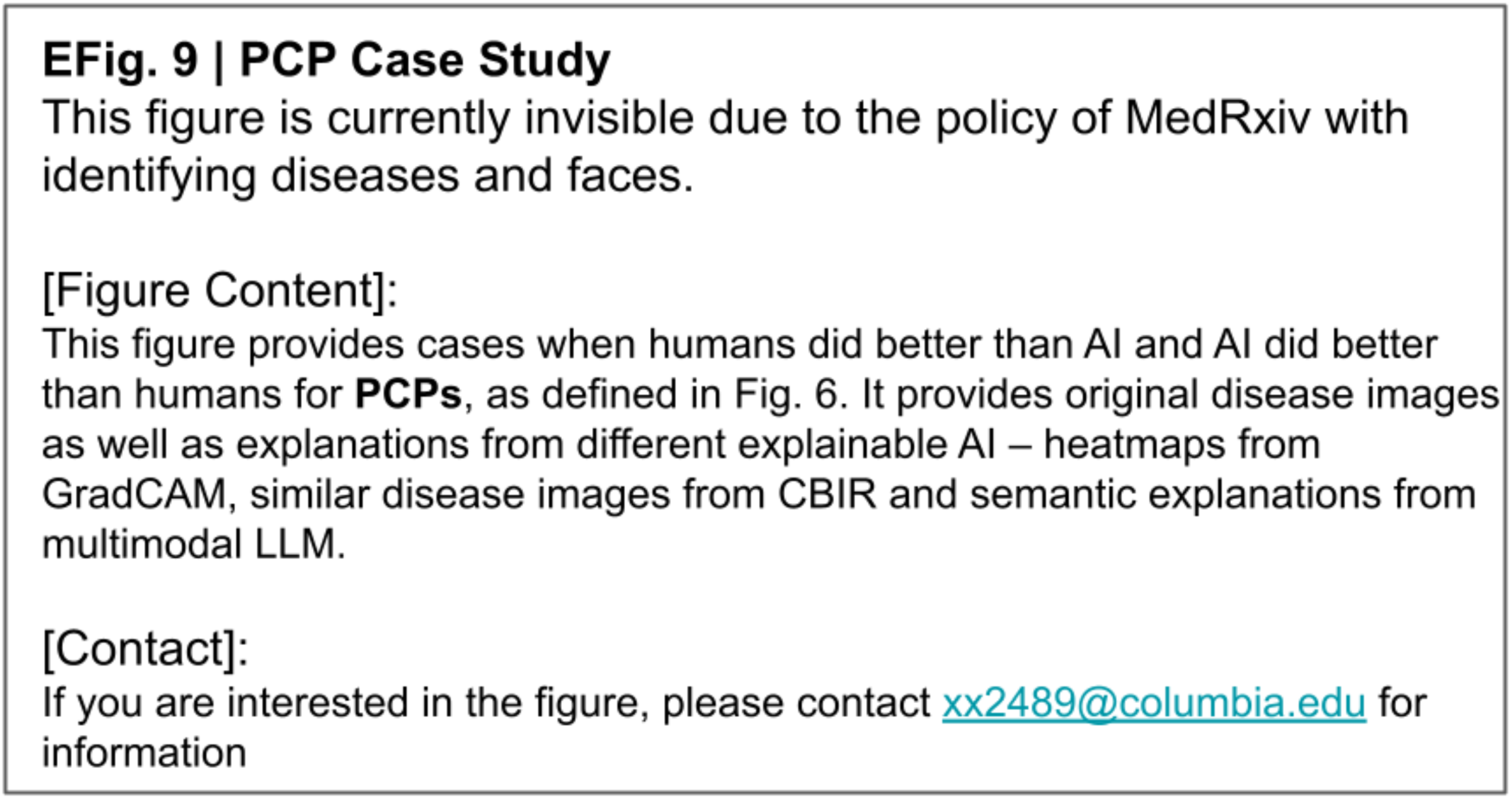
PCP Case Study. **a,** AI had a better diagnostic performance than humans**. b,** Humans did a better performance than AI.

**EFig. 10.**
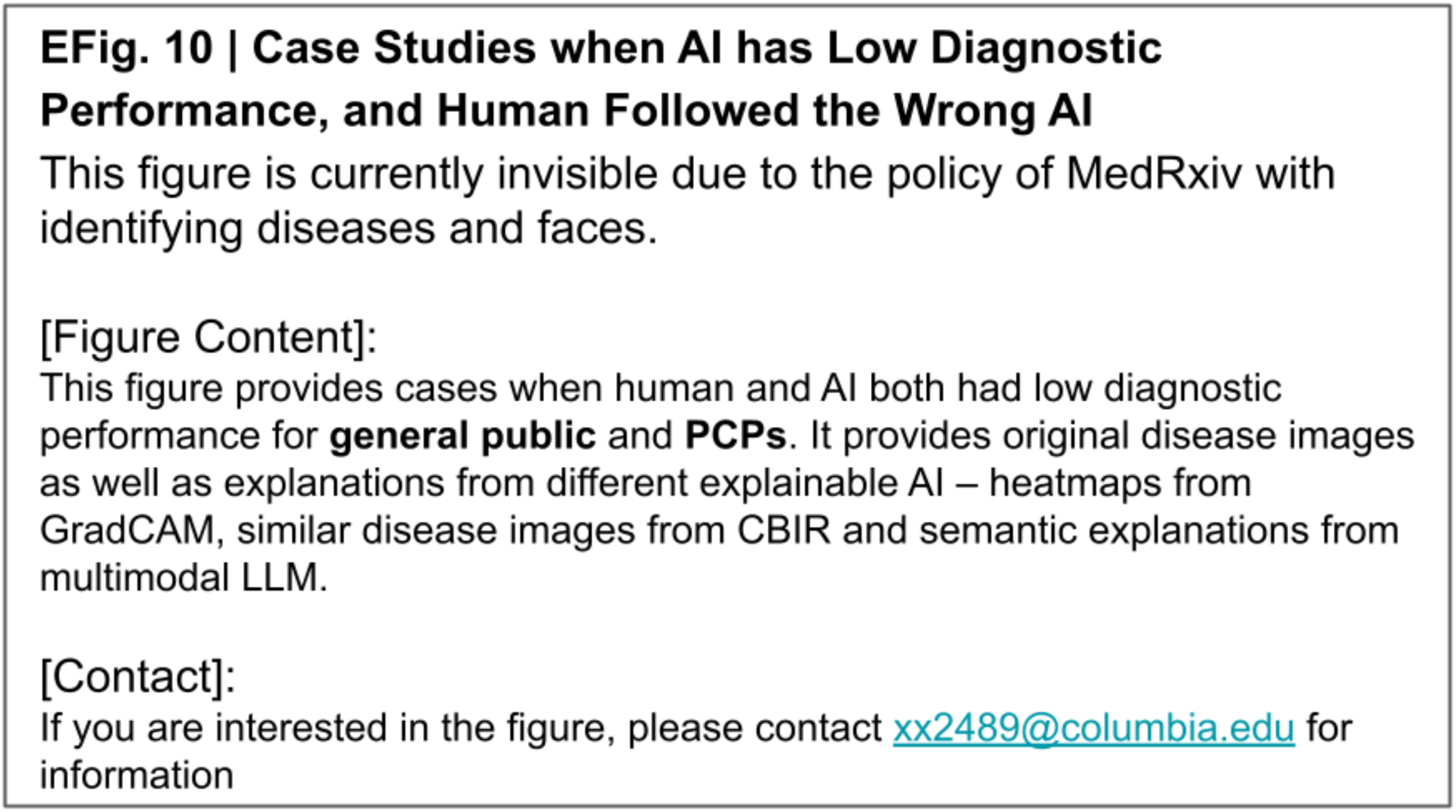
Case Studies when AI has Low Diagnostic Performance, and Human Followed the Wrong AI. a, Examples for the general public. b, Examples for the PCPs.

