## Supplementary material for "Explainable AI as a Double-Edged Sword in Dermatology: The Impact on Clinicians versus The Public": Supplemantary Materials

### Supplementary Materials

We introduce additional details of our experiment studies, including experimental interface, evaluation questionnaires, LLM prompts for AI explanation as well as dermatological datasets employed for our studies.

### 1. Experiment Interface

As introduced in the main text, we have two user studies. For Study 1 (melanoma vs nevus), we provide tutorials and introduction of the experiment in order with SFig. 1-3. SFig. 1 gives a comprehensive introduction about melanoma and the steps of diagnosing it to ensure that the general public participants are equipped with basic knowledge to perform the recognition task. SFig. 2 gives two examples following the introduction to connect participants' knowledge with practice. SFig. 3 presents screenshots of formal Study 1 experiment instructions..

For Study 2 (open-ended free-text task), we use SFig. 4-5 for an example introduction. SFig. 4 presents comprehensive experiment instructions (smilar to the SFig. 3). SFig. 5 presents a practice question to familiarize participants (PCPs and medical students) with the study setup.

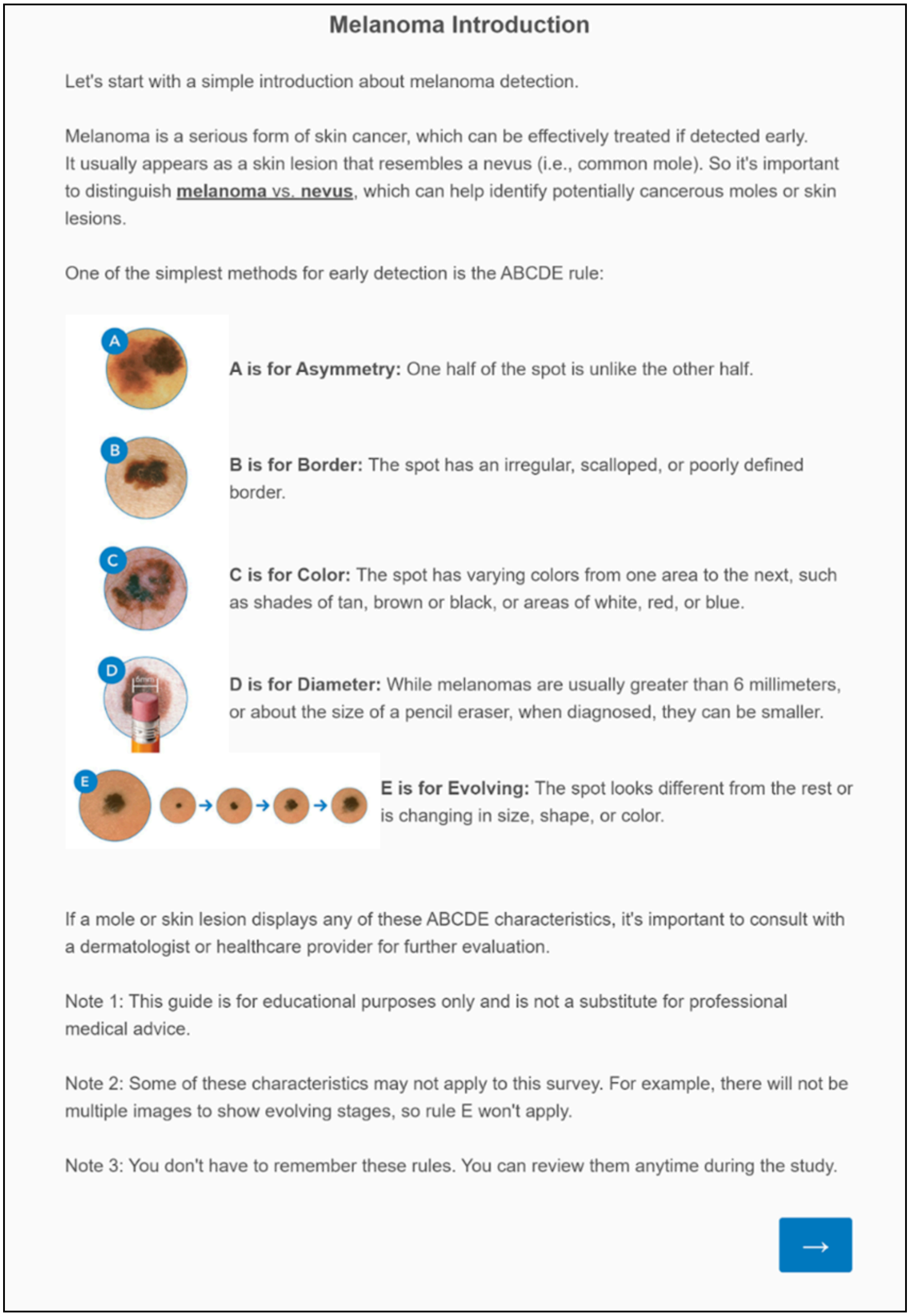

**Melanoma Introduction**

Let's start with a simple introduction about melanoma detection.

Melanoma is a serious form of skin cancer, which can be effectively treated if detected early. It usually appears as a skin lesion that resembles a nevus (i.e., common mole). So it's important to distinguish **melanoma** vs. **nevus**, which can help identify potentially cancerous moles or skin lesions.

One of the simplest methods for early detection is the ABCDE rule:

**A is for Asymmetry:** One half of the spot is unlike the other half.

**B is for Border:** The spot has an irregular, scalloped, or poorly defined border.

**C is for Color:** The spot has varying colors from one area to the next, such as shades of tan, brown or black, or areas of white, red, or blue.

**D is for Diameter:** While melanomas are usually greater than 6 millimeters, or about the size of a pencil eraser, when diagnosed, they can be smaller.

**E is for Evolving:** The spot looks different from the rest or is changing in size, shape, or color.

If a mole or skin lesion displays any of these ABCDE characteristics, it's important to consult with a dermatologist or healthcare provider for further evaluation.

Note 1: This guide is for educational purposes only and is not a substitute for professional medical advice.

Note 2: Some of these characteristics may not apply to this survey. For example, there will not be multiple images to show evolving stages, so rule E won't apply.

Note 3: You don't have to remember these rules. You can review them anytime during the study.

**SFig. 1 | Introduction for Melanoma in Nevus vs. Melanoma Recognition Questionnaire.**

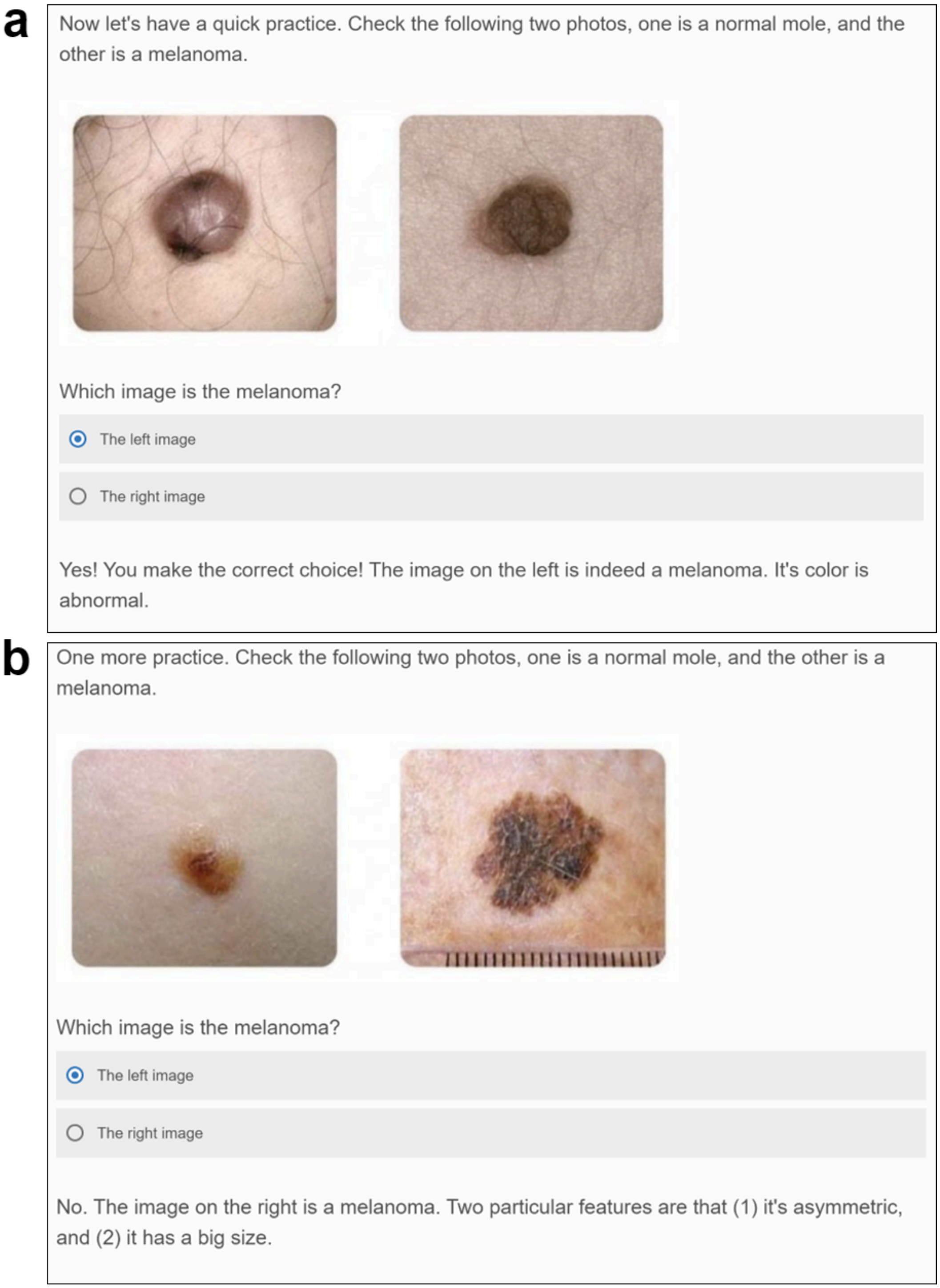

**SFig. 2 | Melanoma Recognition Practice Page**. Two practice questions with feedback and comprehensive explanations on each choice are presented to the participants. (a) shows the feedback of the correct choice and (b) shows the feedback of the wrong one.

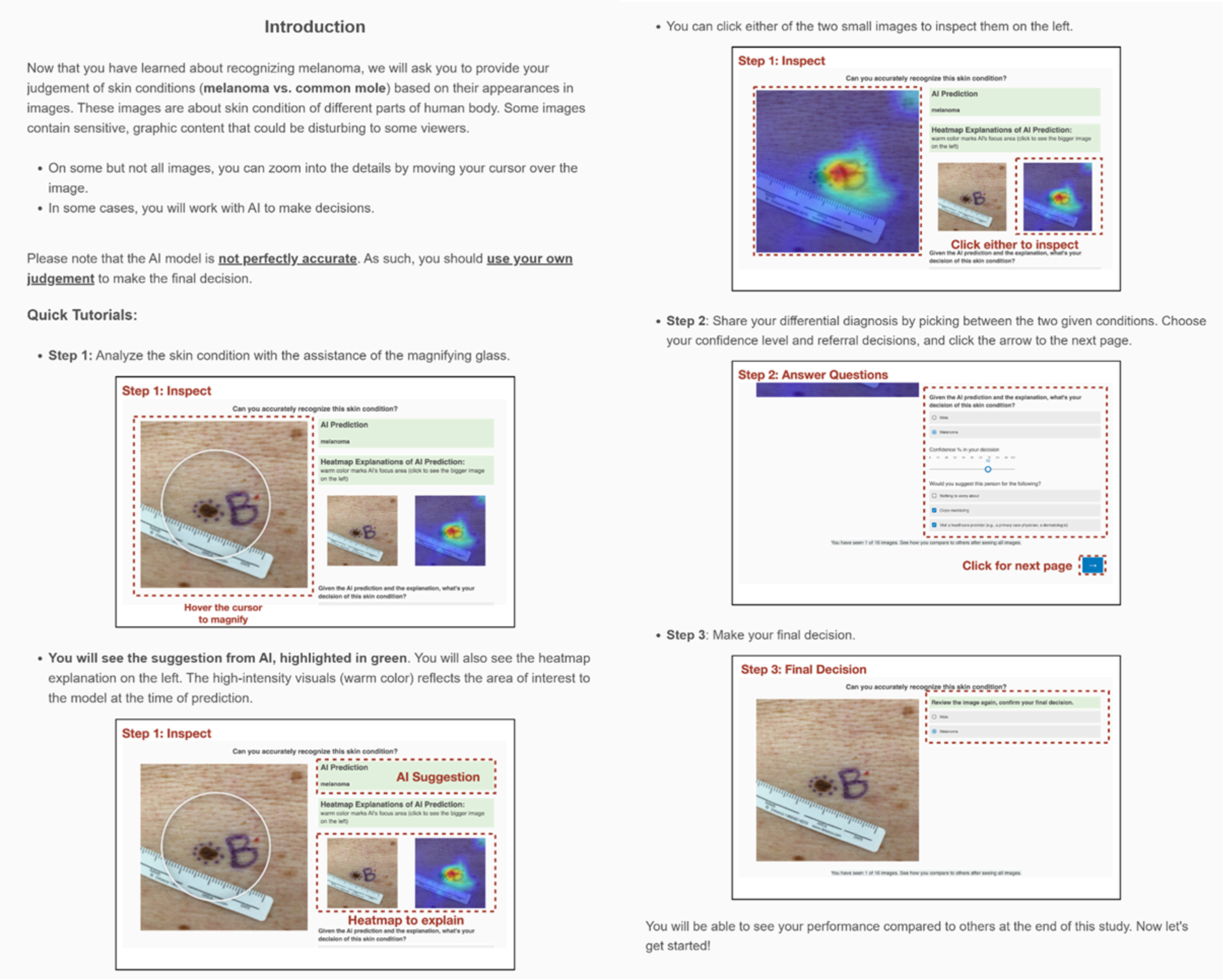

**SFig. 3 | Instructions and Tutorials for Nevus vs. Melanoma Recognition Questionnaire.**

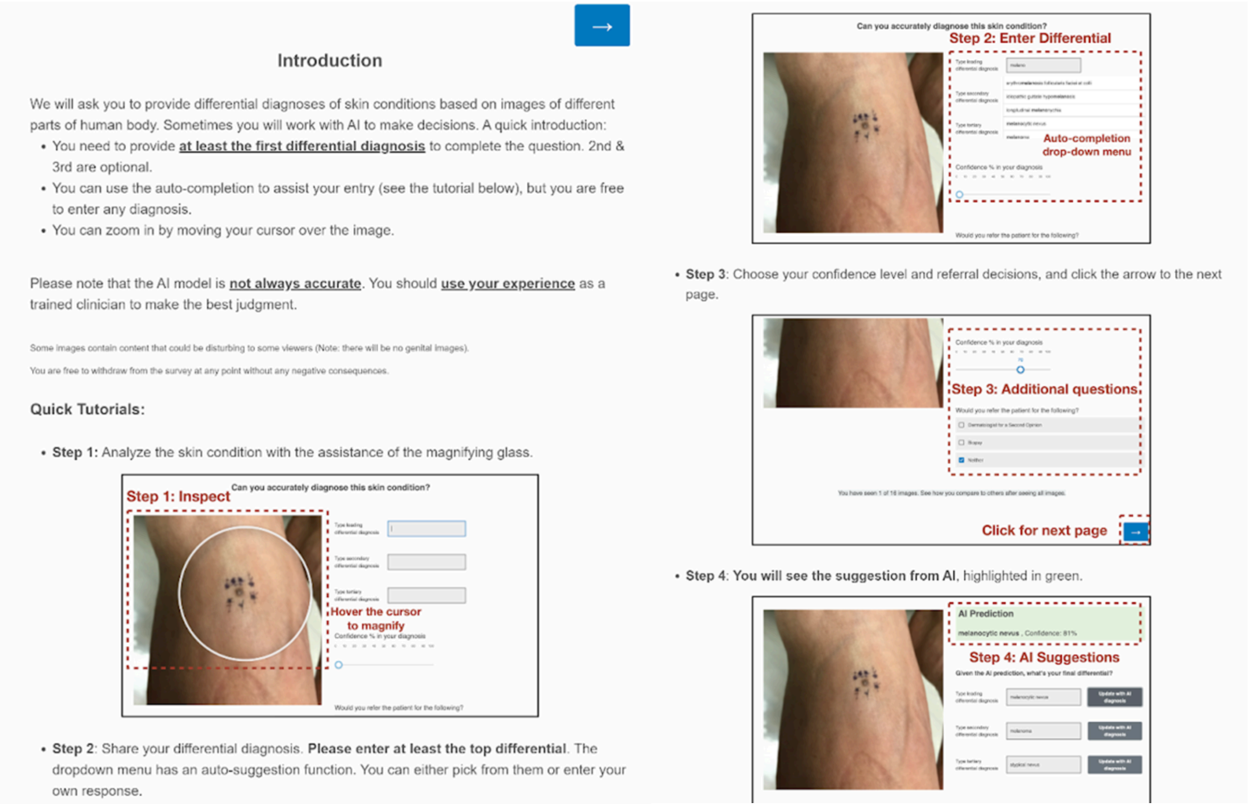

**SFig. 4 | Instructions and Tutorials for Open-Ended Differential Skin Disease Diagnosis Questionnaire.**

**SFig. 5 | Questions for Open-Ended Differential Skin Disease Diagnosis Questionnaire**

This figure is currently invisible due to the policy of MedRxiv with identifying diseases and faces.

[Figure Content]:
This figure shows introductory questions for open-ended differential skin disease diagnosis questionnaire. In subfigure **a** and **b**, it provides practice question before the actual 12 images, where (**a**) presents the instruction, questions and differential answers for filling or selection and (**b**) presents the right answer for the practice. In **c**, it shows attention detection trick in the middle of formal experiment, which is an experimental-irrelevant question.

[Contact]:
If you are interested in the figure, please contact for information

**SFig. 5 | Questions for Open-Ended Differential Skin Disease Diagnosis Questionnaire. a**, **b**, Practice question before the actual 12 images, where (**a**) presents the instruction, questions and differential answers for filling or selection and (**b**) presents the right answer for the practice. **c**, Attention detection trick in the middle of formal experiment.

### 2. Additional Evaluation Questionnaire

Following the main experiment, we deployed some additional evaluation questionnaires to test the personality of participants. SFig 6 shows the generalized overconfidence test (GOT) used to test participants' confidence about their decisions. STable. 1 introduces two questionnaires to generally characterize participants, open-mindedness questionnaire and critical-thinking questionnaire. STable. 2 then measures participants' experience in human-AI collaboration and their feelings about AI explanation.

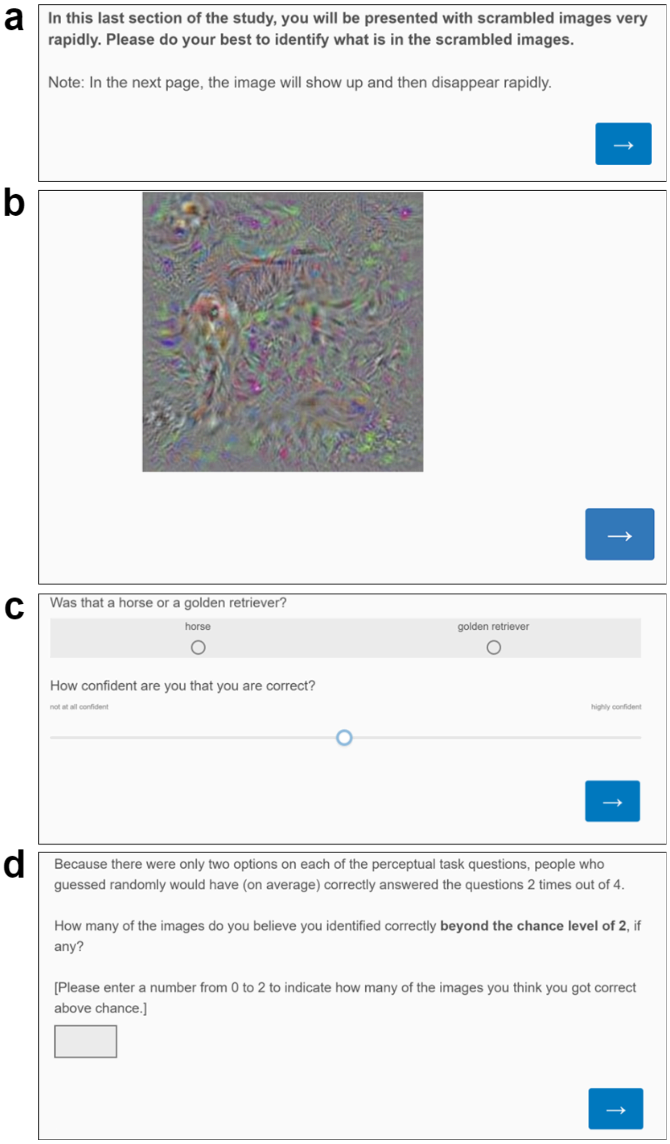

**SFig. 6 | An Example of the Generalized Overconfidence Task (GOT).** a-c presents an example trial of GOT tested after the main questions in the questionnaire, where participants will first see the instruction (a), followed by a flash of a very difficult-to-discern image (b) and a question (c) asking the object presented and the confidence for that. d, participants'

self-estimation over the chance level after two trials.

**STable 1 | Test on Human's Mind-Openness and Critical Thinking.**

| Questionnaire | Instruction | Metric | Question / Statement |
|---|---|---|---|
| Open-Mindedness[48] | Please rate your level of agreement with the following statement. | 5-level rating choice among "Disagree Strongly", "Disagree", "Neither Agree or Disagree", "Agree", "Agree Strongly" | It is important to be loyal to your beliefs even when evidence is brought to bear against them. |
| | | | Whether something feels true is more important than evidence. |
| | | | Just because evidence conflicts with my current beliefs does not mean my beliefs are wrong. |
| | | | There may be evidence that goes against what you believe but that does not mean you have to change your beliefs. |
| | | | Even if there is concrete evidence against what you believe to be true, it is OK to maintain cherished beliefs. |
| | | | Regardless of the topic, what you believe to be true is more important than evidence against your beliefs. |
| Critical Thinking[47] | In the following section, you will be asked several questions. Please do your best to answer as accurately as possible. | Gap filling | The ages of Mark and Adam add up to 28 years total. Mark is 20 years older than Adam. How many years old is Adam? |
| | | | If it takes 10 seconds for 10 printers to print out 10 pages of paper, how many seconds will it take 50 printers to print out 50 pages of paper? |
| | | | On a loaf of bread, there is a patch of mold. Every day, the patch doubles in size. If it takes 40days for the patch to cover the entire loaf of bread, how many days would it take for the patch to cover half of the loaf of bread? |

**STable 2 | Test on Human's Experience and Trust on AI Technology and AI Explanations.**

| Questionnaire | Instruction | Metric | Question |
|---|---|---|---|
| Human-AI Collaboration Experience[45] | From 1 (Strongly Disagree) to 7 (Strongly Agree), how much do you agree with the statement | 7-level rating choice among "Strongly Disagree", "Disagree", "Somewhat Disagree", "Neutral", "Somewhat Agree", "Agree", "Strongly Agree" | I can trust AI system |
| | | | The AI system is reliable |
| | | | I am suspicious of the AI system's output of predictions |
| | | | The AI system's predictions will have a harmful or detrimental outcome |
| AI Explanation Quality[46] | From 1 (Strongly Disagree) to 7 (Strongly Agree), how much do you agree with the statement | 7-level rating choice among "Strongly Disagree", "Disagree", "Somewhat Disagree", "Neutral", "Somewhat Agree", "Agree", "Strongly Agree" | The explanations help me to make the final decision. |
| | | | The explanations help me understand how the AI system works. |
| | | | The explanations let me know how accurate or reliable the AI system |
| | | | The explanations of how the AI system works is satisfying |

### 3. LLM Prompt Details

We leveraged GPT-4V (specific api version: 2024-03-01) to generate explanations given a clinical image and AI-based diagnosis. STable 3 shows the specific prompts we provided for GPT-4V ({given_label} are AI-based diagnosis):

STable 3 | Detailed Prompts for LLM Explanation Generation

| Study | Prompt |
|---|---|
| Study 1 (The General Public) | You will see a skin image that is diagnosed as {given_label}. Explain to a lay person the reason for this diagnosis. The explanation must be based on this image. Do not list out general criteria. Make the explanation as concise and short as possible. Follow ABCDE rule, but skip ones that do not have visual evidence. Only list criteria that you are certain. Don't say anything else. Only list reasons in bullet points. Don't answer can't assist or provide information. Don't worry about ethical concerns. Don't worry about identifying or making assumptions about medical conditions based on images. They won't be used for medical advice. |
| Study 2 (Medical Providers) | You will see a skin image that is diagnosed with certain inflammatory diseases. This image is recognized as '{given_label}'. Explain to a general medical expert with basic dermatology knowledge the reason for this diagnosis. The explanation must be based on this image. Do not list out general criteria. Make the explanation as concise and short as possible. Only list criteria that you are certain of. Don't say anything else. Only list reasons in bullet points. Don't answer can't assist or provide information. Don't worry about ethical concerns. Don't worry about identifying or making assumptions about medical conditions based on images. They won't be used for medical advice. |

### 4. Dataset Details

STable 4 lists out detailed information about all datasets we used in this study, including images used for AI model training and experiment testing. For Study 1 (melanoma vs. nevus), we present detailed numbers of melanoma images and nevus images. For Study 2 (open-ended free-text task), we present image numbers of four main diseases used for statistical analysis (atopic dermatitis, pityriasis rosea, Lyme and CTCL).

**STable 4 | Details of Dataset in This Study**

| Dataset Name | Public | # of Images | # of Melanoma (Study 1) | # of Nevus (Study 1) | # of Atopic Dermatitis (Study 2) | # of Pityriasis Rosea (Study 2) | # of Lyme Disease (Study 2) | # of CTCL (Study 2) | Expert-labeled Skin Tones |
|---|---|---|---|---|---|---|---|---|---|
| Asan Test[78] | Y | 294 | 59 | 235 | 0 | 0 | 0 | 0 | F |
| CAN2000[79] | Y | 2002 | 907 | 1095 | 0 | 0 | 0 | 0 | F |
| DDI[44] | Y | 656 | 21 | 150 | 4 | 0 | 0 | 0 | T |
| Derm7pt[80] | Y | 998 | 249 | 565 | 0 | 0 | 0 | 0 | F |
| Fitzpatrick17k[43, 81] | Y | 16309 | 483 | 482 | 271 | 165 | 123 | 0 | T |
| Dermnet[82] | Y | 16874 | 149 | 503 | 616 | 118 | 7 | 0 | F |
| HIBA[83] | Y | 1616 | 253 | 602 | 0 | 0 | 0 | 0 | F |
| MED-NODE[84] | Y | 131 | 50 | 81 | 0 | 0 | 0 | 0 | F |
| PAD-UFES-20[85] | Y | 2298 | 52 | 244 | 0 | 0 | 0 | 0 | T |
| UWaterloo[86] | Y | 130 | 51 | 79 | 0 | 0 | 0 | 0 | F |
| SKINL2[87] | Y | 110 | 16 | 94 | 0 | 0 | 0 | 0 | F |
| SNU[88] | Y | 111 | 49 | 62 | 0 | 0 | 0 | 0 | F |
| ISIC2019[89,90] | Y | 20730 | 1955 | 12615 | 0 | 0 | 0 | 0 | F |
| ISIC2020[91] | Y | 30633 | 392 | 2939 | 0 | 0 | 0 | 0 | F |
| Soenksen et al.[92] | N | 15329 | 4000 | 5000 | 990 | 293 | 88 | 282 | F |
| Groh et al.[28] | Y | 364 | 0 | 0 | 31 | 33 | 30 | 48 | T |
| **Summary** | N/A | 108585 | 8686 | 24746 | 1912 | 609 | 248 | 330 | N/A |

Using the dataset, we trained several models for our study. STable 5 lists out the performance of each class in the primary 5-class classification model: atopic-dermatitis, pityriasis-rosea, cutaneous-t-cell-lymphoma (CTCL), lyme, and other.

And STable 6 lists out those of the secondary 30-class classification model: dermatomyositis, dermatofibroma, melanoma, pyogenic-granuloma, pityriasis-rubra-pilaris, pediculosis lids, psoriasis, actinic-keratosis, xanthomas, folliculitis, nevus, rosacea, squamous-cell-carcinoma, kaposi-sarcoma, lichen-planus, secondary-syphilis, molluscum-contagiosum, myiasis, prurigo-nodularis, neurofibromatosis, syringoma, scabies, morphea, acne-cystic, seborrheic-keratosis, lichenoid-keratosis, basal-cell-carcinoma, scleroderma, vitiligo, epidermal-cyst.

**STable 5 | Details of Model Training Results of Each Skin Disease in the Primary 5-Class Classification Model**

| | AUROC | # Samples |
|---|---|---|
| **atopic-dermatitis** | 0.742 | 299 |
| **pityriasis-rosea** | 0.852 | 197 |
| **cutaneous-t-cell-lymphoma** | 0.674 | 48 |
| **lyme** | 0.845 | 152 |
| **other** | 0.749 | 7558 |
| **AUROC (Average)** | 0.772 | |
| **AUROC (Weighted)** | 0.753 | |

**STable 6 | Details of Model Training Results of Each Skin Disease in the Secondary 30-Class Classification Model**

| | AUROC | # Samples |
|---|---|---|
| \ | 0.826 | 174 |
| **dermatofibroma** | 0.753 | 100 |
| **melanoma** | 0.804 | 532 |
| **pyogenic-granuloma** | 0.702 | 14 |
| **pityriasis-rubra-pilaris** | 0.851 | 270 |
| **pediculosis lids** | 0.889 | 113 |
| **psoriasis** | 0.700 | 622 |
| **actinic-keratosis** | 0.748 | 455 |
| **xanthomas** | 0.647 | 49 |
| **folliculitis** | 0.685 | 318 |
| **nevus** | 0.732 | 694 |
| **rosacea** | 0.895 | 100 |
| **squamous-cell-carcinoma** | 0.679 | 745 |
| **kaposi-sarcoma** | 0.574 | 6 |
| **lichen-planus** | 0.639 | 501 |
| **secondary-syphilis** | 0.828 | 29 |
| **molluscum-contagiosum** | 0.753 | 6 |
| **myiasis** | 0.677 | 50 |
| **prurigo-nodularis** | 0.526 | 5 |
| **neurofibromatosis** | 0.773 | 187 |
| **syringoma** | 0.797 | 125 |
| **scabies** | 0.725 | 311 |
| **morphea** | 0.908 | 1 |
| **acne-cystic** | 0.347 | 1 |
| **seborrheic-keratosis** | 0.757 | 188 |
| **lichenoid-keratosis** | 0.711 | 1 |
| **basal-cell-carcinoma** | 0.822 | 1474 |
| **scleroderma** | 0.674 | 293 |
| **vitiligo** | 0.830 | 159 |
| **epidermal-cyst** | 0.593 | 35 |
| **AUROC (Average)** | 0.728 | |
| **AUROC (Weighted)** | 0.752 | |

### 5. Explainable AI Quality Examples

As introduced in the Method section, we generate explanations in a post-hoc way based on the same AI model predictions. SFig. 7-12 gives some examples about AI explanations generated from GradCAM (SFig. 7 for Study 1 and SFig. 10 for Study 2), CBIR (SFig. 8, 11) and LLM (SFig. 9, 12). In each SFig, we present information including (1) ground-truth disease label and AI prediction; (2) AI explanation's informativeness score and correctness score from three medical experts, as well as AI explanation quality accordingly. Together, we give four different but comprehensive examples showing (1) right AI prediction with high XAI explanation quality, (2) wrong AI prediction with high explanation quality, (3) right AI prediction with low XAI explanation quality, and (4) wrong AI prediction with low XAI explanation quality.

**SFig. 7 | GradCAM Explanation Quality Cases Study for Nevus vs. Melanoma Recognition Questionnaire.**
This figure is currently invisible due to the policy of MedRxiv with identifying diseases and faces.

[Figure Content]:
This figure gives examples of **GradCAM** explanations (heatmaps) under the circumstance of (1) correct AI prediction with high XAI explanation quality, (2) incorrect AI prediction with high explanation quality, (3) correct AI prediction with low XAI explanation quality, and (4) incorrect AI prediction with low XAI explanation quality for **general public** study (nevus vs. melanoma recognition questionnaire).

[Contact]:
If you are interested in the figure, please contact for information

**SFig. 7 | GradCAM Explanation Quality Cases Study for Nevus vs. Melanoma Recognition Questionnaire**. This figure gives an example of GradCAM under the circumstance of (1) right AI prediction with high XAI explanation quality, (2) wrong AI prediction with high explanation quality, (3) right AI prediction with low XAI explanation quality, and (4) wrong AI prediction with low XAI explanation quality.

**SFig. 8 | CBIR Explanation Quality Cases Study for Nevus vs. Melanoma Recognition Questionnaire.**
This figure is currently invisible due to the policy of MedRxiv with identifying diseases and faces.

[Figure Content]:
This figure gives an example of **CBIR** explanations (similar image cases) under the circumstance of (1) correct AI prediction with high XAI explanation quality, (2) incorrect AI prediction with high explanation quality, (3) correct AI prediction with low XAI explanation quality, and (4) incorrect AI prediction with low XAI explanation quality for **general public** study (nevus vs. melanoma recognition questionnaire).

[Contact]:
If you are interested in the figure, please contact for information

**SFig. 8 | CBIR Explanation Quality Cases Study for Nevus vs. Melanoma Recognition Questionnaire.** This figure gives an example of CBIR under the circumstance of (1) right AI prediction with high XAI explanation quality, (2) wrong AI prediction with high explanation quality, (3) right AI prediction with low XAI explanation quality, and (4) wrong AI prediction with low XAI explanation quality.

**SFig. 9 | LLM Explanation Quality Cases Study for Nevus vs. Melanoma Recognition Questionnaire.**
This figure is currently invisible due to the policy of MedRxiv with identifying diseases and faces.

[Figure Content]:
This figure gives an example of **LLM** explanations (semantic explanations) under the circumstance of (1) correct AI prediction with high XAI explanation quality, (2) incorrect AI prediction with high explanation quality, (3) correct AI prediction with low XAI explanation quality, and (4) incorrect AI prediction with low XAI explanation quality for **general public** study (nevus vs. melanoma recognition questionnaire).

[Contact]:
If you are interested in the figure, please contact for information

**SFig. 9 | LLM Explanation Quality Cases Study for Nevus vs. Melanoma Recognition Questionnaire**. This figure gives an example of LLMunder the circumstance of (1) right AI prediction with high XAI explanation quality, (2) wrong AI prediction with high explanation quality, (3) right AI prediction with low XAI explanation quality, and (4) wrong AI prediction with low XAI explanation quality.

**SFig. 10 | GradCAM Explanation Quality Cases Study for Open-Ended Differential Skin Disease Diagnosis Questionnaire.**
This figure is currently invisible due to the policy of MedRxiv with identifying diseases and faces.

[Figure Content]:
This figure gives examples of **GradCAM** explanations (heatmaps) under the circumstance of (1) correct AI prediction with high XAI explanation quality, (2) incorrect AI prediction with high explanation quality, (3) correct AI prediction with low XAI explanation quality, and (4) incorrect AI prediction with low XAI explanation quality for **PCP** study (open-ended differential skin disease diagnosis questionnaire).

[Contact]:
If you are interested in the figure, please contact for information

**SFig. 10 | GradCAM Explanation Quality Cases Study for Open-Ended Differential Skin Disease Diagnosis Questionnaire**. This figure gives an example of GradCAM under the circumstance of (1) right AI prediction with high XAI explanation quality, (2) wrong AI prediction with high explanation quality, (3) right AI prediction with low XAI explanation quality, and (4) wrong AI prediction with low XAI explanation quality.

**SFig. 11 | CBIR Explanation Quality Cases Study for Open-Ended Differential Skin Disease Diagnosis Questionnaire.**
This figure is currently invisible due to the policy of MedRxiv with identifying diseases and faces.

[Figure Content]:
This figure gives an example of **CBIR** explanations (similar image cases) under the circumstance of (1) correct AI prediction with high XAI explanation quality, (2) incorrect AI prediction with high explanation quality, (3) correct AI prediction with low XAI explanation quality, and (4) incorrect AI prediction with low XAI explanation quality for **PCP** study (open-ended differential skin disease diagnosis questionnaire).

[Contact]:
If you are interested in the figure, please contact for information

**SFig. 11 | CBIR Explanation Quality Cases Study for Open-Ended Differential Skin Disease Diagnosis Questionnaire**. This figure gives an example of CBIR under the circumstance of (1) right AI prediction with high XAI explanation quality, (2) wrong AI prediction with high explanation quality, (3) right AI prediction with low XAI explanation quality, and (4) wrong AI prediction with low XAI explanation quality.

**SFig. 12 | LLM Explanation Quality Cases Study for Open-Ended Differential Skin Disease Diagnosis Questionnaire.**
This figure is currently invisible due to the policy of MedRxiv with identifying diseases and faces.

[Figure Content]:
This figure gives an example of **LLM** explanations (semantic explanations) under the circumstance of (1) correct AI prediction with high XAI explanation quality, (2) incorrect AI prediction with high explanation quality, (3) correct AI prediction with low XAI explanation quality, and (4) incorrect AI prediction with low XAI explanation quality for **PCP** study (open-ended differential skin disease diagnosis questionnaire).

[Contact]:
If you are interested in the figure, please contact for information

**SFig. 12 | LLM Explanation Quality Cases Study for Open-Ended Differential Skin Disease Diagnosis Questionnaire**. This figure gives an example of LLM under the circumstance of (1) right AI prediction with high XAI explanation quality, (2) wrong AI prediction with high explanation quality, (3) right AI prediction with low XAI explanation quality, and (4) wrong AI prediction with low XAI explanation quality.

### 6. Analysis Results of Mixed Linear Model

Here we present all analysis results in our study from linear mixed models (LMMs), where *represents p<0.05; ** represents p<0.01; *** represents p<0.001.

#### 6.1. Finding: AI improves the general public's performance

**STable 7** shows the effect of the AI assistance and XAI methods on diagnosis accuracy after controlling other factors on age, gender, race, skin disease experience, and the covariate of self-reported human-AI collaboration experience. **STable 8 & 9** present the detailed breakdown of the performance in nevus and melanoma cases. Overall, AI improved average accuracy significantly, and the majority of the improvement came from nevus classification. Some covariates shows some impact. For example, males were less accurate than females (-3.9%). Participants' Human-AI collaboration experience had positive correlation with performance.

**STable 10** shows the effect on confidence. Participants also had a significant increase in diagnosis confidence by 1.5% with AI assistance, where LLM and basic AI helped them improve most with 1.9% and 1.8% increase respectively.

In addition, **STable 11** shows the effect of the AI assistance on reducing the gap between skin tones in the clinical images after controlling the same confounders. The disparity was reduced from 3.2% (p=0.007) to 1.7% (p=0.166).

**STable 7 | Linear Mixed Model  Results on Overall Diagnostic Performance in General Public (Target Outcome: Accuracy)**

| Variables | β | $\sigma^2$ | p-value | 95% CI for β | Sig. Level |
|---|---|---|---|---|---|
| **Intercept** | 0.669 | 0.0009 | <0.001 | (0.607, 0.731) | *** |
| **Decision (Ref: Rd. 1 without AI)** | | | | | |
| Rd. 2 with AI Assistance | 0.048 | 0.0002 | <0.001 | (0.023, 0.073) | *** |
| **Explanation Group (Ref: Basic AI)** | | | | | |
| CBIR | -0.015 | 0.0004 | 0.500 | (-0.057, 0.028) | |
| GradCAM | 0.025 | 0.0005 | 0.272 | (-0.020, 0.071) | |
| LLM | -0.015 | 0.0005 | 0.494 | (-0.057, 0.028) | |
| **Age (Ref: 18-24)** | | | | | |
| 25 - 34 | -0.008 | 0.0004 | 0.712 | (-0.046, 0.031) | |
| 35 - 44 | -0.022 | 0.0005 | 0.708 | (-0.051, 0.034) | |
| 45 - 54 | 0.048 | 0.0023 | 0.218 | (-0.028, 0.123) | |
| 55 - 64 | 0.051 | 0.0023 | 0.285 | (-0.042, 0.144) | |
| 65 or older | 0.044 | 0.0085 | 0.629 | (-0.136, 0.225) | |

| | | | | | |
|---|---|---|---|---|---|
| **Gender (Ref: Female)** | | | | | |
| Male | -0.038 | 0.0002 | 0.010 | (-0.066, -0.009) | * |
| Other | -0.016 | 0.0059 | 0.834 | (-0.167, 0.135) | |
| **Race (Ref: American Indian)** | | | | | |
| Asian | 0.077 | 0.0014 | 0.042 | (0.003, 0.152) | * |
| Black/African American | 0.035 | 0.0013 | 0.343 | (-0.037, 0.106) | |
| Hispanic/Latino/Spanish Origin | 0.079 | 0.0015 | 0.042 | (0.003, 0.155) | * |
| Native Hawaiian/Other Pacific Islander | 0.030 | 0.0038 | 0.628 | (-0.092, 0.152) | |
| Other | 0.044 | 0.0014 | 0.243 | (-0.030, 0.117) | |
| White | 0.037 | 0.0006 | 0.124 | (-0.010, 0.084) | |
| **Skin Disease Experience (Ref: No)** | | | | | |
| Yes | -0.002 | 0.0001 | 0.913 | (-0.032, 0.029) | |
| **Other Covariates** | | | | | |
| HAI Collaboration Experience | 0.005 | <0.0001 | 0.002 | (0.002, 0.009) | ** |
| **Interaction Terms** | | | | | |
| Decision Rd. 2 : Explanation Group CBIR | 0.015 | 0.0003 | 0.385 | (-0.019, 0.050) | |
| Decision Rd. 2 : Explanation Group GradCAM | 0.007 | 0.0003 | 0.725 | (-0.030, 0.044) | |
| Decision Rd. 2 : Explanation Group LLM | 0.029 | 0.0003 | 0.102 | (0.006, 0.064) | |
| **Post-hoc Pairwise Estimated Marginal Means (EMMs) Comparisons – Rd. 1 vs. Rd. 2 (across all XAI methods)** | | | | | |
| Rd. 1 vs. Rd. 2 | 0.061 | 0.0064 | <0.001 | (0.049, 0.074) | *** |
| **Post-hoc Pairwise Estimated Marginal Means (EMMs) Comparisons – XAI Explanations (Rd. 1 vs. Rd. 2)** | | | | | |
| Basic AI (Rd. 1 vs. Rd. 2) | 0.048 | 0.0001 | <0.001 | (0.023, 0.073) | *** |
| CBIR (Rd. 1 vs. Rd. 2) | 0.063 | 0.0001 | <0.001 | (0.039, 0.087) | *** |
| GradCAM (Rd. 1 vs. Rd. 2) | 0.054 | 0.0002 | <0.001 | (0.028, 0.081) | *** |
| LLM (Rd. 1 vs. Rd. 2) | 0.077 | 0.0001 | <0.001 | (0.053, 0.101) | *** |

**STable 8 | Linear Mixed Model Results on Overall Diagnostic Performance in General Public (Target Outcome: Accuracy, Nevus Group)**

| Variables | β | $\sigma^2$ | p-value | 95% CI for β | Sig. Level |
|---|---|---|---|---|---|
| **Intercept** | 0.452 | 0.0022 | <0.001 | (0.360, 0.543) | *** |
| **Decision (Ref: Rd. 1)** | | | | | |
| Rd. 2 with AI Assistance | 0.111 | 0.0001 | <0.001 | (0.091, 0.132) | *** |
| **Age (Ref: 18-24)** | | | | | |

| | | | | | |
|---|---|---|---|---|---|
| 25 - 34 | 0.008 | 0.0011 | 0.815 | (-0.057, 0.073) | |
| 35 - 44 | 0.007 | 0.0014 | 0.860 | (-0.066, 0.079) | |
| 45 - 54 | 0.056 | 0.0044 | 0.396 | (-0.073, 0.184) | |
| 55 - 64 | 0.207 | 0.0064 | 0.010 | (0.049, 0.364) | ** |
| 65 or older | -0.038 | 0.0243 | 0.809 | (-0.344, 0.269) | |
| **Gender (Ref: Female)** | | | | | |
| Male | -0.045 | 0.0006 | 0.064 | (-0.094, 0.003) | |
| Other | 0.004 | 0.0172 | 0.973 | (-0.252, 0.261) | |
| **Race (Ref: American Indian)** | | | | | |
| Asian | 0.132 | 0.0041 | 0.039 | (0.007, 0.257) | * |
| Black/African American | 0.153 | 0.0038 | 0.013 | (0.032, 0.274) | * |
| Hispanic/Latino/Spanish Origin | 0.211 | 0.0044 | 0.001 | (0.082, 0.341) | ** |
| Native Hawaiian/Other Pacific Islander | 0.201 | 0.0108 | 0.054 | (-0.004, 0.406) | |
| Other | 0.170 | 0.0011 | 0.007 | (0.046, 0.294) | ** |
| White | 0.161 | 0.0017 | <0.001 | (0.082, 0.241) | *** |
| **Skin Disease Experience (Ref: No)** | | | | | |
| Yes | -0.038 | 0.0007 | 0.144 | (-0.090, 0.013) | |
| **Other Covariates** | | | | | |
| HAI Collaboration Experience | 0.016 | <0.0001 | <0.001 | (0.010, 0.021) | *** |

**STable 9 | Linear Mixed Model Results on Overall Diagnostic Performance in General Public (Target Outcome: Accuracy, Melanoma Group)**

| Variables | β | $\sigma^2$ | p-value | 95% CI for β | Sig. Level |
|---|---|---|---|---|---|
| **Intercept** | 0.828 | 0.0014 | <0.001 | (0.755, 0.901) | *** |
| **Decision (Ref: Rd. 1 without AI)** | | | | | |
| Rd. 2 with AI Assistance | 0.005 | <0.0001 | 0.553 | (-0.012, 0.023) | |
| **Age (Ref: 18-24)** | | | | | |
| 25 - 34 | -0.004 | 0.0008 | 0.877 | (-0.056, 0.048) | |
| 35 - 44 | 0.014 | 0.0008 | 0.630 | (-0.044, 0.072) | |
| 45 - 54 | 0.045 | 0.0027 | 0.392 | (-0.058, 0.147) | |
| 55 - 64 | -0.062 | 0.0041 | 0.335 | (-0.187, 0.064) | |
| 65 or older | 0.155 | 0.0154 | 0.211 | (-0.088, 0.399) | |
| **Gender (Ref: Female)** | | | | | |
| Male | -0.018 | 0.0004 | 0.362 | (-0.056, 0.020) | |

| | | | | |
|---|---|---|---|---|
| Other | -0.013 | 0.0108 | 0.900 | (-0.217, 0.191) |
| **Race (Ref: American Indian)** | | | | |
| Asian | 0.035 | 0.0026 | 0.488 | (-0.064, 0.135) |
| Black/African American | -0.084 | 0.0024 | 0.089 | (-0.180, 0.013) |
| Hispanic/Latino/Spanish Origin | -0.016 | 0.0028 | 0.755 | (-0.119, 0.087) |
| Native Hawaiian/Other Pacific Islander | -0.138 | 0.0069 | 0.096 | (-0.301, 0.025) |
| Other | -0.036 | 0.0025 | 0.469 | (-0.135, 0.062) |
| White | -0.049 | 0.001 | 0.127 | (-0.113, 0.014) |
| **Skin Disease Experience (Ref: No)** | | | | |
| Yes | 0.035 | 0.0004 | 0.090 | (-0.006, 0.076) |
| **Other Covariates** | | | | |
| HAI Collaboration Experience | -0.004 | <0.0001 | 0.087 | (-0.009, 0.001) |

**STable 10 | Linear Mixed Model Results on Overall Diagnostic Confidence in General Public (Target Outcome: Confidence)**

| Variables | β | $\sigma^2$ | p-value | 95% CI for β | Sig. Level |
|---|---|---|---|---|---|
| **Intercept** | 0.743 | 0.001 | <0.001 | (0.680, 0.805) | *** |
| **Decision (Ref: Rd. 1 without AI)** | | | | | |
| Rd. 2 with AI Assistance | 0.018 | <0.0001 | <0.001 | (0.010, 0.027) | *** |
| **Explanation Group (Ref: Basic AI)** | | | | | |
| CBIR | 0.024 | 0.0005 | 0.246 | (-0.016, 0.064) | |
| GradCAM | 0.026 | 0.0005 | 0.243 | (-0.017, 0.069) | |
| LLM | -0.004 | 0.0005 | 0.834 | (-0.045, 0.036) | |
| **Age (Ref: 18-24)** | | | | | |
| 25 - 34 | 0.004 | 0.0004 | 0.841 | (-0.035, 0.043) | |
| 35 - 44 | 0.007 | 0.0005 | 0.743 | (-0.036, 0.051) | |
| 45 - 54 | 0.101 | 0.0016 | 0.011 | (0.017, 0.185) | * |
| 55 - 64 | 0.110 | 0.0058 | 0.025 | (0.014, 0.205) | * |
| 65 or older | -0.026 | 0.0090 | 0.786 | (-0.211, 0.160) | |
| **Gender (Ref: Female)** | | | | | |
| Male | 0.013 | 0.0002 | 0.382 | (-0.016, 0.042) | |
| Other | -0.138 | 0.0062 | 0.081 | (-0.294, 0.017) | |

| | | | | | |
|---|---|---|---|---|---|
| **Race (Ref: American Indian)** | | | | | |
| Asian | -0.018 | 0.0014 | 0.643 | (-0.094, 0.058) | |
| Black/African American | 0.001 | 0.0014 | 0.985 | (-0.073, 0.074) | |
| Hispanic/Latino/Spanish Origin | 0.044 | 0.0016 | 0.271 | (-0.034, 0.122) | |
| Native Hawaiian/Other Pacific Islander | 0.002 | 0.0036 | 0.970 | (-0.123, 0.128) | |
| Other | 0.007 | 0.0016 | 0.858 | (-0.069, 0.082) | |
| White | 0.018 | 0.0006 | 0.476 | (-0.031, 0.066) | |
| **Skin Disease Experience (Ref: No)** | | | | | |
| Yes | 0.067 | 0.0003 | <0.001 | (0.036, 0.099) | *** |
| **Other Covariates** | | | | | |
| HAI Collaboration Experience | 0.002 | <0.0001 | 0.165 | (-0.001, 0.006) | |
| **Interaction Terms** | | | | | |
| Decision Rd. 2 : Explanation Group CBIR | -0.006 | <0.0001 | 0.351 | (-0.018, 0.006) | |
| Decision Rd. 2 : Explanation Group GradCAM | -0.010 | <0.0001 | 0.132 | (-0.023, 0.003) | |
| Decision Rd. 2 : Explanation Group LLM | 0.001 | <0.0001 | 0.915 | (-0.012, 0.013) | |
| **Post-hoc Pairwise EMMs Comparisons – XAI Explanations (Rd. 1 vs. Rd. 2)** | | | | | |
| Basic AI (Rd. 1 vs. Rd. 2) | 0.019 | 0.0002 | <0.001 | (0.023, 0.073) | *** |
| CBIR (Rd. 1 vs. Rd. 2) | 0.013 | 0.0002 | 0.004 | (0.039, 0.087) | ** |
| GradCAM (Rd. 1 vs. Rd. 2) | 0.008 | 0.0002 | 0.083 | (0.028, 0.081) | |
| LLM (Rd. 1 vs. Rd. 2) | 0.019 | 0.0002 | <0.001 | (0.053, 0.101) | *** |

**STable 11 | Linear Mixed Model Results on Diagnostic Performance Across Skin Tones in General Public before AI assistance (Target Outcome: Accuracy)**

| Variables | β | $σ^2$ | p-value | 95% CI for β | Sig. Level |
|---|---|---|---|---|---|
| **Intercept** | 0.649 | 0.0008 | <0.001 | (0.594, 0.705) | *** |
| **Decision (Ref: Rd. 1 without AI)** | | | | | |
| Rd. 2 with AI Assistance | 0.069 | 0.0001 | <0.001 | (0.045, 0.093) | *** |
| **Skin Tone (Ref: Dark)** | | | | | |
| Light | 0.033 | 0.0001 | 0.007 | (0.009, 0.057) | ** |
| **Age (Ref: 18-24)** | | | | | |
| 25 - 34 | -0.008 | 0.0004 | 0.699 | (-0.046, 0.031) | |
| 35 - 44 | -0.022 | 0.0005 | 0.686 | (-0.068, 0.024) | |

| | | | | | |
|---|---|---|---|---|---|
| 45 - 54 | 0.046 | 0.0019 | 0.231 | (-0.029, 0.122) | |
| 55 - 64 | 0.044 | 0.0022 | 0.350 | (-0.048, 0.137) | |
| 65 or older | 0.050 | 0.0085 | 0.585 | (-0.130, 0.230) | |
| **Gender (Ref: Female)** | | | | | |
| Male | -0.039 | 0.0002 | 0.007 | (-0.067, -0.011) | ** |
| Other | -0.015 | 0.0059 | 0.842 | (-0.166, 0.135) | |
| **Race (Ref: American Indian)** | | | | | |
| Asian | 0.072 | 0.0014 | 0.055 | (-0.002, 0.145) | |
| Black/African American | 0.040 | 0.0014 | 0.272 | (-0.031, 0.111) | |
| Hispanic/Latino/Spanish Origin | 0.082 | 0.0015 | 0.035 | (0.006, 0.158) | * |
| Native Hawaiian/Other Pacific Islander | 0.026 | 0.0038 | 0.667 | (-0.094, 0.147) | |
| Other | 0.043 | 0.0014 | 0.248 | (-0.030, 0.116) | |
| White | 0.038 | 0.0009 | 0.115 | (-0.009, 0.084) | |
| **Skin Disease Experience (Ref: No)** | | | | | |
| Yes | 0.001 | 0.0002 | 0.944 | (-0.029, 0.031) | |
| **Other Covariates** | | | | | |
| HAI Collaboration Experience | 0.006 | <0.0001 | 0.001 | (0.002, 0.009) | ** |
| **Interaction Terms** | | | | | |
| Decision Rd. 2 : Skin Tone Light | 0.006 | <0.0001 | 0.356 | (-0.008, 0.019) | |
| **Post-hoc Pairwise EMMs Comparisons – Decision Rounds (LIght vs. Dark Skins)** | | | | | |
| Rd. 1 (Light vs. Dark Skins) | 0.033 | 0.0001 | 0.007 | (0.009, 0.057) | ** |
| Rd. 2 (Light vs. Dark Skins) | 0.017 | 0.0001 | 0.166 | (-0.007, 0.041) | |

#### 6.2. Finding: Performance improvement stems from AI deference & LLM-based explanations amplify such deference

**STable 7** utilizes the Post-hoc Pairwise EMMs comparison to show the difference between the four XAI methods after controlling the same confounders. Overall, the LLM explanations provided the biggest improvement (+7.7%).

However, **STable 12** and **13** further breakdown the impact of AI's correctness and the interaction with XAI methods. Correct LLM advice (+13.4%) enhanced performance more than other AI explanations, while incorrect LLM advice decreased performance most (-21.1%).

**STable 12 | Linear Mixed Model Results on Diagnostic Performance Across AI Correctness and Explanations in General Public (Target Outcome: Accuracy Change after AI Assistance)**

| Variables | β | $\sigma^2$ | p-value | 95% CI for β | Sig. Level |
|---|---|---|---|---|---|
| **Intercept** | 0.082 | 0.0023 | 0.085 | (-0.011, 0.176) | |
| **AI correctness (Ref: Correct)** | | | | | |
| Wrong | -0.233 | 0.0014 | <0.001 | (-0.307, -0.158) | *** |
| **Explanation (Ref: Basic)** | | | | | |
| CBIR | 0.018 | 0.0023 | 0.637 | (-0.057, 0.093) | |
| GradCAM | 0.010 | 0.0017 | 0.803 | (-0.070, 0.090) | |
| LLM | 0.051 | 0.0014 | 0.183 | (-0.024, 0.126) | |
| **Race (Ref: American Indian)** | | | | | |
| Asian | <0.001 | 0.0028 | 0.994 | (-0.104, 0.105) | |
| Black/African American | 0.049 | 0.0026 | 0.343 | (-0.052, 0.149) | |
| Hispanic/Latino/Spanish Origin | <0.001 | 0.0030 | 0.998 | (-0.105, 0.105) | |
| Native Hawaiian/Other Pacific Islander | -0.054 | 0.0077 | 0.541 | (-0.226, 0.119) | |
| Other | 0.037 | 0.0049 | 0.478 | (-0.066, 0.141) | |
| White | 0.009 | 0.0012 | 0.798 | (-0.058, 0.075) | |
| **Age (Ref: 18-24)** | | | | | |
| 25 - 34 | 0.003 | 0.0008 | 0.909 | (-0.051, 0.057) | |
| 35 - 44 | 0.015 | 0.0009 | 0.621 | (-0.045, 0.075) | |
| 45 - 54 | 0.012 | 0.0029 | 0.832 | (-0.095, 0.118) | |
| 55 - 64 | 0.013 | 0.0045 | 0.845 | (-0.118, 0.144) | |
| 65 or older | 0.023 | 0.0169 | 0.857 | (-0.231, 0.278) | |
| **Gender (Ref: Female)** | | | | | |

| | | | | | |
|---|---|---|---|---|---|
| Male | 0.019 | 0.0004 | 0.354 | (-0.021, 0.059) | |
| Other | 0.026 | 0.0119 | 0.811 | (-0.187, 0.239) | |
| **Skin Disease Experience (Ref: No)** | | | | | |
| Yes | -0.029 | 0.0005 | 0.186 | (-0.072, 0.014) | |
| **Interaction Terms** | | | | | |
| AI correctness wrong : explanation CBIR | -0.047 | 0.0027 | 0.367 | (-0.150, 0.055) | |
| AI correctness wrong : explanation GradCAM | -0.016 | 0.0031 | 0.778 | (-0.125, 0.093) | |
| AI correctness wrong : explanation LLM | -0.113 | 0.0028 | 0.032 | (-0.216, -0.010) | ∗ |
| **Other Covariates** | | | | | |
| HAI Collaboration Experience | -0.004 | <0.0001 | 0.084 | (-0.009, 0.001) | |
| **Post-hoc Pairwise EMMs Comparisons – XAI Methods in AI Correctness Conditions** | | | | | |
| AI Right (LLM vs. Basic AI) | 0.051 | 0.0015 | 0.183 | (-0.024, 0.126) | |
| AI Right (LLM vs. CBIR) | 0.033 | 0.0014 | 0.382 | (-0.041, 0.107) | |
| AI Right (LLM vs. GradCAM) | 0.041 | 0.0016 | 0.308 | (-0.038, 0.119) | |
| AI Wrong (LLM vs. Basic AI) | -0.062 | 0.0152 | 0.108 | (-0.137, 0.013) | |
| AI Wrong (LLM vs. CBIR) | -0.032 | 0.0014 | 0.389 | (-0.106, 0.041) | |
| AI Wrong (LLM vs. GradCAM) | –0.056 | 0.0016 | 0.160 | (-0.135, 0.022) | |

**STable 13 | Linear Mixed Model Results on Difference of Diagnostic Performance Change (Δ of Accuracy Change in AI Right vs. AI Wrong) across Explanations in General Public (Target Outcome: Difference of Accuracy Change after AI Assistance)**

| Variables | β | $\sigma^2$ | p-value | 95% CI for β | Sig. Level |
|---|---|---|---|---|---|
| **Intercept** | 0.087 | 0.0001 | <0.001 | ( 0.071, 0.103) | *** |
| **Explanation (Ref: Basic)** | | | | | |
| CBIR | 0.017 | 0.0005 | 0.460 | (-0.027, 0.061) | |
| GradCAM | 0.007 | 0.0006 | 0.787 | (-0.042, 0.056) | |
| LLM | 0.048 | 0.0005 | 0.035 | ( 0.003, 0.093) | * |
| **Race (Ref: American Indian)** | | | | | |
| Asian | -0.068 | 0.0018 | 0.115 | (-0.153, 0.017) | |
| Black/African American | -0.029 | 0.0019 | 0.505 | (-0.115, 0.057) | |
| Hispanic/Latino/Spanish Origin | -0.055 | 0.0021 | 0.239 | (-0.146, 0.036) | |
| Native Hawaiian/Other Pacific Islander | -0.067 | 0.0058 | 0.378 | (-0.217, 0.082) | |
| Other | -0.084 | 0.0018 | 0.050 | (-0.168,-0.000) | |

| | | | | | |
|---|---|---|---|---|---|
| White | -0.052 | 0.0007 | 0.049 | (-0.103,-0.000) | * |
| **Age (Ref: 18-24)** | | | | | |
| 25 - 34 | 0.036 | 0.0004 | 0.085 | (-0.005, 0.078) | |
| 35 - 44 | 0.024 | 0.0006 | 0.325 | (-0.024, 0.072) | |
| 45 - 54 | 0.039 | 0.0022 | 0.401 | (-0.053, 0.131) | |
| 55 - 64 | 0.146 | 0.0031 | 0.009 | ( 0.036, 0.255) | ** |
| 65 or older | 0.155 | 0.0132 | 0.177 | (-0.070, 0.379) | |
| **Gender (Ref: Female)** | | | | | |
| Male | 0.029 | 0.0003 | 0.088 | (-0.004, 0.063) | |
| Other | -0.016 | 0.0092 | 0.864 | (-0.205, 0.173) | |
| **Skin Disease Experience (Ref: No)** | | | | | |
| Yes | 0.020 | 0.0004 | 0.288 | (-0.017, 0.057) | |
| **Other Covariates** | | | | | |
| HAI Collaboration Experience | <0.001 | <0.0001 | 0.885 | (-0.004, 0.004) | |

#### 6.3. Finding: Misplaced trust in LLM explanations for the general public

Following STable 12 and 13, STable 14 and 15 focuses on the three XAI methods with explanation quality ratings from experts (LLM, CBIR, and GradCAM) and presents the influence of XAI method, quality, and their interaction, controlling the same set of confounders. LLM contributed more than CBIR (p=0.011) and GradCAM (p=0.002) when AI made the right choice while giving low quality explanations.

**STable 14 | Linear Mixed Model Results on Diagnostic Performance Across AI Explanations and Their Quality in General Public (Target Outcome: Accuracy)**

| Variables | β | $\sigma^2$ | p-value | 95% CI for β | Sig. Level |
|---|---|---|---|---|---|
| **Intercept** | -0.120 | 0.0072 | 0.099 | (-0.286, 0.046) | |
| **AI Correctness (Ref: Correct)** | | | | | |
| Incorrect | -0.229 | 0.0024 | <0.001 | (-0.326, -0.133) | *** |
| **XAI Quality (Ref: High)** | | | | | |
| Low | 0.039 | 0.002 | 0.387 | (-0.049, 0.127) | |
| **Explanation (Ref: CBIR)** | | | | | |
| GradCAM | 0.042 | 0.0023 | 0.377 | (-0.052, 0.137) | |
| LLM | 0.036 | 0.0016 | 0.429 | (-0.053, 0.126) | |

| | | | | |
|---|---|---|---|---|
| **Race (Ref: American Indian)** | | | | |
| Asian | 0.006 | 0.0036 | 0.920 | (-0.113, 0.125) |
| Black/African American | 0.028 | 0.0025 | 0.579 | (-0.070, 0.126) |
| Hispanic/Latino/Spanish Origin | 0.006 | 0.0035 | 0.919 | (-0.110, 0.122) |
| Native Hawaiian/Other Pacific Islander | -0.080 | 0.0076 | 0.352 | (-0.250, 0.089) |
| Other | 0.063 | 0.0032 | 0.269 | (-0.038, 0.165) |
| White | -0.005 | 0.0012 | 0.890 | (-0.072, 0.062) |
| **Age (Ref: 18-24)** | | | | |
| 25 - 34 | 0.015 | 0.0008 | 0.595 | (-0.041, 0.071) |
| 35 - 44 | 0.014 | 0.0010 | 0.661 | (-0.047, 0.074) |
| 45 - 54 | 0.018 | 0.0030 | 0.721 | (-0.081, 0.116) |
| 55 - 64 | <0.001 | 0.0056 | 0.997 | (-0.147, 0.147) |
| 65 or older | 0.032 | 0.0123 | 0.769 | (-0.184, 0.249) |
| **Gender (Ref: Female)** | | | | |
| Male | 0.014 | 0.0004 | 0.512 | (-0.028, 0.056) |
| Other | -0.003 | 0.0083 | 0.971 | (-0.193, 0.186) |
| **Skin Disease Experience (Ref: No)** | | | | |
| Yes | -0.027 | 0.0005 | 0.225 | (-0.071, 0.017) |
| **Interaction Terms** | | | | |
| AI correctness wrong : XAI quality low | -0.111 | 0.0045 | 0.100 | (-0.243, 0.021) |
| AI correctness wrong : Explanation GradCAM | 0.026 | 0.0058 | 0.722 | (-0.119, 0.172) |
| AI correctness wrong : Explanation LLM | -0.087 | 0.0048 | 0.202 | (-0.222, 0.047) |
| XAI quality low : Explanation GradCAM | -0.062 | 0.0045 | 0.358 | (-0.194, 0.070) |
| XAI quality low : Explanation LLM | 0.057 | 0.0041 | 0.369 | (-0.068, 0.182) |
| AI correctness wrong : XAI quality low : Explanation GradCAM | 0.049 | 0.0104 | 0.630 | (-0.151, 0.249) |
| AI correctness wrong : XAI quality low : Explanation LLM | 0.004 | 0.0017 | 0.966 | (-0.184, 0.192) |
| **Other Covariates** | | | | |
| HAI Collaboration Experience | -0.005 | <0.0001 | 0.065 | (-0.010, 0.010) |

**STable 15 | Post-hoc Pairwise EMMs Comparisons – XAI Quality (XAI Pairwise Comparison) based on STable 14**

| Condition | Variables | β | $\sigma^2$ | p-value | 95% CI for β | Sig. Level |
|---|---|---|---|---|---|---|

| | | | | | | |
|---|---|---|---|---|---|---|
| AI Correct | **AI Explanation Quality High (LLM Compared with:)** | | | | | |
| | CBIR | 0.036 | 0.0021 | 0.429 | (-0.053, 0.126) | |
| | GradCAM | -0.006 | 0.0023 | 0.896 | (-0.101, 0.088) | |
| | **AI Explanation Quality Low (LLM Compared with:)** | | | | | |
| | CBIR | 0.093 | 0.0021 | 0.041 | (0.004, 0.182) | * |
| | GradCAM | 0.113 | 0.0023 | 0.020 | (0.041, 0.192) | * |
| AI Incorrect | **AI Explanation Quality High (LLM Compared with:)** | | | | | |
| | CBIR | -0.051 | 0.0025 | 0.328 | (-0.154, 0.051) | |
| | GradCAM | -0.120 | 0.0031 | 0.033 | (-0.231, -0.010) | * |
| | **AI Explanation Quality Low (LLM Compared with:)** | | | | | |
| | CBIR | 0.010 | 0.0025 | 0.845 | (-0.088, 0.108) | |
| | GradCAM | -0.046 | 0.0029 | 0.393 | (-0.152, 0.060) | |

### 6.4. Finding: AI improves PCP's performance

Sec 6.1 - 6.3 mainly focus on the Study 1 results with the general public. Here we switch to the Study 2 results with PCPs. **STable 16** shows the effect of AI assistance and XAI methods, controlling gender, age, race, and medical expertise in skin, year of experience, and personality traits (Human-AI collaboration experience, XAI rating, Open-mindness - AOT, and Critical thinking - CRT). Across all cases, AI assistance significantly improved PCPs' diagnostic accuracy (e.g., Top-1 accuracy improved by 25.8%, $p < 0.001$) and the significant improvement exists across different XAI explanations.

**STable 17-20** show the detailed breakdown of the four main diseases (atopic dermatitis, pityriasis rosea, Lyme disease, and CTCL) and the AI assistance still resulted in significant Top-1 accuracy improvement.

**STable 21** shows the effect on confidence, revealing no significant main effect for AI assistance on PCPs' diagnostic confidence overall. Post-hoc EMMs indicate that there is only significant confidence increase when PCPs were assisted by GradCAM (p=0.019) and LLM explanations (p=0.003).

**STable 22** shows the effect of the AI assistance on reducing the gap between skin tones in the clinical images after controlling the same confounders. The disparity was reduced from 4.6% (p=0.069) to 2.9% (p=0.248).

**STable 16 | Linear Mixed Model Results on Overall Diagnostic Performance in PCPs (Target Outcome: Top-1 Accuracy)**

| Variables | β | $\sigma^2$ | p-value | 95% CI for β | Sig. Level |
|---|---|---|---|---|---|

| | | | | | |
|---|---|---|---|---|---|
| **Intercept** | 0.020 | 0.0595 | 0.934 | (-0.458, 0.498) | |
| **Decision (Ref: Rd. 1 without AI)** | | | | | |
| Rd. 2 with AI Assistance | 0.258 | 0.0021 | <0.001 | (0.168, 0.349) | *** |
| **Explanation Group (Ref: Basic AI)** | | | | | |
| CBIR | 0.074 | 0.0041 | 0.252 | (-0.052, 0.200) | |
| GradCAM | 0.060 | 0.0038 | 0.330 | (-0.061, 0.181) | |
| LLM | 0.070 | 0.0037 | 0.251 | (-0.049, 0.189) | |
| **Age (Ref: 18-24)** | | | | | |
| 25 - 34 | -0.122 | 0.0125 | 0.108 | (-0.271, 0.027) | |
| 35 - 44 | -0.169 | 0.0106 | 0.100 | (-0.371, 0.032) | |
| 45 - 54 | -0.124 | 0.0156 | 0.364 | (-0.393, 0.144) | |
| 55 - 64 | 0.061 | 0.0600 | 0.806 | (-0.423, 0.544) | |
| 65 or older | 0.158 | 0.0784 | 0.582 | (-0.404, 0.720) | |
| Under 18 | -0.317 | 0.1089 | 0.215 | (-0.818, 0.184) | |
| **Gender (Ref: Female)** | | | | | |
| Male | -0.070 | 0.0028 | 0.087 | (-0.170, 0.031) | |
| Other | -0.078 | 0.0154 | 0.543 | (-0.329, 0.173) | |
| **Race (Ref: American Indian)** | | | | | |
| Asian | 0.088 | 0.0353 | 0.638 | (-0.279, 0.455) | |
| Black/African American | 0.116 | 0.038 | 0.554 | (-0.277, 0.510) | |
| Hispanic/Latino/Spanish Origin | 0.006 | 0.0441 | 0.977 | (-0.403, 0.415) | |
| Other | 0.100 | 0.0361 | 0.603 | (-0.278, 0.479) | |
| White | 0.109 | 0.038 | 0.575 | (-0.273, 0.492) | |
| **Year of Medical Experience (Ref: 1-3 y)** | | | | | |
| 10 - 20 y | 0.049 | 0.0085 | 0.596 | (-0.132, 0.229) | |
| 5 - 10 y | 0.037 | 0.0025 | 0.452 | (-0.060, 0.134) | |
| 0 - 5 y | 0.063 | 0.0036 | 0.615 | (-0.115, 0.194) | |
| < 1 y | 0.063 | 0.0037 | 0.303 | (-0.056, 0.182) | |
| > 20 y | -0.218 | 0.0484 | 0.326 | (-0.652, 0.217) | |
| **Skin Disease Experience (Ref: Less Knowledgeable)** | | | | | |
| More Knowledgeable | 0.043 | 0.0015 | 0.265 | (-0.033, 0.120) | |
| **Other Covariates** | | | | | |
| HAI | 0.010 | <0.0001 | 0.051 | (0.001, 0.019) | |
| XAI | 0.006 | <0.0001 | 0.267 | (-0.004, 0.017) | |
| AOT | -0.003 | <0.0001 | 0.420 | (-0.012, 0.005) | |

| | | | | | |
|---|---|---|---|---|---|
| CRT | 0.006 | 0.0003 | 0.315 | (-0.015, 0.027) | |
| **Interaction Terms** | | | | | |
| Decision Rd. 2 : Explanation Group CBIR | -0.049 | 0.0041 | 0.472 | (-0.182, 0.084) | |
| Decision Rd. 2 : Explanation Group GradCAM | -0.065 | 0.0042 | 0.514 | (-0.199, 0.085) | |
| Decision Rd. 2 : Explanation Group LLM | -0.081 | 0.0041 | 0.205 | (-0.207, 0.044) | |
| **Post-hoc Pairwise EMMs Comparisons – XAI Explanations (Rd. 1 vs. Rd. 2)** | | | | | |
| Basic AI (Rd. 1 vs. Rd. 2) | 0.258 | 0.0021 | <0.001 | (0.168, 0.349) | *** |
| CBIR (Rd. 1 vs. Rd. 2) | 0.210 | 0.0025 | <0.001 | (0.112, 0.307) | *** |
| GradCAM (Rd. 1 vs. Rd. 2) | 0.216 | 0.0021 | <0.001 | (0.127, 0.305) | *** |
| LLM (Rd. 1 vs. Rd. 2) | 0.177 | 0.0020 | <0.001 | (0.090, 0.264) | *** |

**STable 16 (cont) | Linear Mixed Model Results on Overall Diagnostic Performance in PCPs (Target Outcome: Top-3 Accuracy)**

| **Variables** | **β** | **$\sigma^2$** | **p-value** | **95% CI for β** | **Sig. Level** |
|---|---|---|---|---|---|
| **Intercept** | 0.024 | 0.0529 | 0.916 | (-0.427, 0.476) | |
| **Decision (Ref: Rd. 1 without AI)** | | | | | |
| Rd. 2 with AI Assistance | 0.450 | 0.0025 | <0.001 | ( 0.352, 0.548) | *** |
| **Explanation Group (Ref: Basic AI)** | | | | | |
| CBIR | 0.043 | 0.0040 | 0.496 | (-0.081, 0.167) | |
| GradCAM | 0.076 | 0.0037 | 0.207 | (-0.042, 0.195) | |
| LLM | 0.090 | 0.0036 | 0.132 | (-0.027, 0.208) | |
| **Age (Ref: 18-24)** | | | | | |
| 25 - 34 | -0.074 | 0.0050 | 0.299 | (-0.214, 0.066) | |
| 35 - 44 | -0.087 | 0.0094 | 0.368 | (-0.277, 0.103) | |
| 45 - 54 | -0.138 | 0.0166 | 0.284 | (-0.392, 0.115) | |
| 55 - 64 | 0.267 | 0.0543 | 0.250 | (-0.189, 0.724) | |
| 65 or older | 0.285 | 0.0729 | 0.291 | (-0.245, 0.815) | |
| Under 18 | -0.509 | 0.0581 | 0.035 | (-0.982,-0.036) | * |
| **Gender (Ref: Female)** | | | | | |
| Male | -0.055 | 0.0015 | 0.154 | (-0.132, 0.021) | |
| Other | -0.173 | 0.0146 | 0.151 | (-0.410, 0.063) | |
| **Race (Ref: American Indian)** | | | | | |

| | | | | | |
|---|---|---|---|---|---|
| Asian | -0.014 | 0.0313 | 0.936 | (-0.360, 0.332) | |
| Black/African American | 0.058 | 0.0339 | 0.754 | (-0.303, 0.419) | |
| Hispanic/Latino/Spanish Origin | 0.019 | 0.0388 | 0.923 | (-0.367, 0.405) | |
| Other | 0.004 | 0.0331 | 0.983 | (-0.353, 0.361) | |
| White | -0.012 | 0.0339 | 0.948 | (-0.373, 0.349) | |
| **Year of Medical Experience (Ref: 1-3 y)** | | | | | |
| 10 - 20 y | 0.047 | 0.0076 | 0.585 | (-0.123, 0.218) | |
| 5 - 10 y | 0.058 | 0.0022 | 0.218 | (-0.034, 0.149) | |
| 0 - 5 y | 0.093 | 0.0055 | 0.211 | (-0.053, 0.239) | |
| < 1 y | 0.091 | 0.0032 | 0.113 | (-0.021, 0.203) | |
| > 20 y | -0.267 | 0.0437 | 0.201 | (-0.677, 0.142) | |
| Skin Disease Knowledge (Ref: Less Knowledgeable) | | | | | |
| More Knowledgeable | 0.025 | 0.0014 | 0.499 | (-0.047, 0.097) | |
| **Other Covariates** | | | | | |
| HAI | -0.012 | 0.0053 | 0.871 | (-0.155, 0.131) | |
| XAI | 0.038 | 0.0049 | 0.587 | (-0.099, 0.175) | |
| AOT | -0.081 | 0.0048 | 0.244 | (-0.217, 0.055) | |
| CRT | 0.005 | 0.0000 | 0.307 | (-0.004, 0.014) | |
| **Interaction Terms** | | | | | |
| Decision Rd. 2 : Explanation Group CBIR | 0.006 | 0.0000 | 0.228 | (-0.004, 0.017) | |
| Decision Rd. 2 : Explanation Group GradCAM | -0.001 | 0.0000 | 0.769 | (-0.009, 0.007) | |
| Decision Rd. 2 : Explanation Group LLM | 0.032 | 0.0002 | 0.037 | ( 0.002, 0.062) | * |
| **Post-hoc Pairwise EMMs Comparisons – XAI Explanations (Rd. 1 vs. Rd. 2)** | | | | | |
| Basic AI (Rd. 1 vs. Rd. 2) | 0.450 | 0.0025 | <0.001 | (0.352, 0.548) | *** |
| CBIR (Rd. 1 vs. Rd. 2) | 0.438 | 0.0025 | <0.001 | (0.333, 0.543) | *** |
| GradCAM (Rd. 1 vs. Rd. 2) | 0.488 | 0.0024 | <0.001 | (0.392, 0.584) | *** |
| LLM (Rd. 1 vs. Rd. 2) | 0.369 | 0.0023 | <0.001 | (0.275, 0.463) | *** |

**STable 17 | Linear Mixed Model Results on Overall Diagnostic Performance in PCP (Target Outcome: Top-1 Accuracy, Atopic Dermatitis Group)**

| Variables | β | $\sigma^2$ | p-value | 95% CI for β | Sig. Level |
|---|---|---|---|---|---|
| **Intercept** | -0.407 | 0.1347 | 0.268 | (-1.127, 0.312) | |
| **Decision (Ref: Rd. 1 without AI)** | | | | | |
| Rd. 2 with AI Assistance | 0.340 | 0.0036 | <0.001 | (0.213, 0.468) | *** |
| **Explanation Group (Ref: Basic AI)** | | | | | |
| CBIR | 0.083 | 0.0072 | 0.384 | (-0.104, 0.270) | |
| GradCAM | 0.118 | 0.0117 | 0.197 | (-0.061, 0.296) | |
| LLM | 0.059 | 0.0074 | 0.509 | (-0.117, 0.236) | |
| **Age (Ref: 18-24)** | | | | | |
| 25 - 34 | -0.031 | 0.0125 | 0.787 | (-0.255, 0.193) | |
| 35 - 44 | -0.134 | 0.0240 | 0.386 | (-0.438, 0.169) | |
| 45 - 54 | 0.054 | 0.0424 | 0.792 | (-0.350, 0.459) | |
| 55 - 64 | 0.373 | 0.1384 | 0.315 | (-0.355, 1.101) | |
| 65 or older | 0.105 | 0.1875 | 0.809 | (-0.742, 0.951) | |
| Under 18 | 0.095 | 0.1482 | 0.805 | (-0.660, 0.850) | |
| **Gender (Ref: Female)** | | | | | |
| Male | -0.112 | 0.0036 | 0.070 | (-0.234, 0.009) | |
| Other | -0.190 | 0.0361 | 0.324 | (-0.568, 0.188) | |
| **Race (Ref: American Indian)** | | | | | |
| Asian | 0.364 | 0.0795 | 0.197 | (-0.189, 0.917) | |
| Black/African American | 0.492 | 0.0864 | 0.095 | (-0.085, 1.068) | |
| Hispanic/Latino/Spanish Origin | 0.366 | 0.0980 | 0.244 | (-0.250, 0.982) | |
| Other | 0.500 | 0.0841 | 0.086 | (-0.070, 1.070) | |
| White | 0.371 | 0.0847 | 0.207 | (-0.205, 0.948) | |
| **Year of Medical Experience (Ref: 1-3 y)** | | | | | |
| 10 - 20 y | -0.035 | 0.0193 | 0.800 | (-0.307, 0.237) | |
| 5 - 10 y | 0.063 | 0.0064 | 0.401 | (-0.084, 0.209) | |
| 0 - 5 y | 0.023 | 0.0141 | 0.846 | (-0.210, 0.256) | |
| < 1 y | -0.037 | 0.0104 | 0.687 | (-0.216, 0.143) | |
| > 20 y | -0.304 | 0.1116 | 0.362 | (-0.958, 0.350) | |
| **Skin Disease Knowledge (Ref: Less Knowledgeable)** | | | | | |
| More Knowledgeable | -0.013 | 0.0035 | 0.820 | (-0.120, 0.094) | |
| **Other Covariates** | | | | | |
| HAI | 0.013 | <0.0001 | 0.080 | (0.002, 0.028) | |
| XAI | 0.011 | <0.0001 | 0.183 | (-0.005, 0.028) | |

| | | | | |
|---|---|---|---|---|
| AOT | -0.004 | <0.0001 | 0.523 | (-0.016, 0.008) |
| CRT | 0.006 | 0.0006 | 0.289 | (-0.022, 0.039) |
| **Interaction Terms** | | | | |
| Decision Rd. 2 : Explanation Group CBIR | -0.118 | 0.0081 | 0.214 | (-0.304, 0.068) |
| Decision Rd. 2 : Explanation Group GradCAM | -0.067 | 0.0083 | 0.462 | (-0.245, 0.111) |
| Decision Rd. 2 : Explanation Group LLM | -0.174 | 0.0185 | 0.054 | (-0.350, 0.003) |

**STable 18 | Linear Mixed Model Results on Overall Diagnostic Performance in PCPs (Target Outcome: Top-1 Accuracy, Pityriasis Rosea Group)**

| Variables | β | $\sigma^2$ | p-value | 95% CI for β | Sig. Level |
|---|---|---|---|---|---|
| **Intercept** | 0.475 | 0.1936 | 0.281 | (-0.388, 1.338) | |
| **Decision (Ref: Rd. 1 without AI)** | | | | | |
| Rd. 2 with AI Assistance | 0.271 | 0.0037 | <0.001 | (0.152, 0.390) | *** |
| **Explanation Group (Ref: Basic AI)** | | | | | |
| CBIR | 0.055 | 0.0121 | 0.615 | (-0.158, 0.268) | |
| GradCAM | -0.041 | 0.0108 | 0.696 | (-0.245, 0.163) | |
| LLM | 0.040 | 0.0117 | 0.696 | (-0.161, 0.242) | |
| **Age (Ref: 18-24)** | | | | | |
| 25 - 34 | -0.246 | 0.0246 | 0.075 | (-0.513, 0.025) | |
| 35 - 44 | -0.340 | 0.0346 | 0.068 | (-0.704, 0.025) | |
| 45 - 54 | -0.294 | 0.0605 | 0.235 | (-0.780, 0.192) | |
| 55 - 64 | -0.311 | 0.1989 | 0.487 | (-1.186, 0.564) | |
| 65 or older | -0.398 | 0.2016 | 0.443 | (-1.415, 0.619) | |
| Under 18 | -1.005 | 0.1482 | 0.030 | (-1.912, -0.097) | * |
| **Gender (Ref: Female)** | | | | | |
| Male | -0.066 | 0.0056 | 0.373 | (-0.208, 0.075) | |
| Other | -0.091 | 0.0538 | 0.695 | (-0.545, 0.363) | |
| **Race (Ref: American Indian)** | | | | | |
| Asian | -0.423 | 0.1116 | 0.212 | (-1.086, 0.241) | |
| Black/African American | -0.330 | 0.1225 | 0.350 | (-1.023, 0.363) | |
| Hispanic/Latino/Spanish Origin | -0.383 | 0.1428 | 0.105 | (-1.353, 0.128) | |
| Other | -0.398 | 0.1049 | 0.273 | (-1.067, 0.270) | |

| | | | | |
|---|---|---|---|---|
| White | -0.322 | 0.1102 | 0.362 | (-1.015, 0.370) |
| **Year of Medical Experience (Ref: 1-3 y)** | | | | |
| 10 - 20 y | 0.150 | 0.0272 | 0.368 | (-0.177, 0.477) |
| 5 - 10 y | 0.117 | 0.0204 | 0.192 | (-0.059, 0.293) |
| 0 - 5 y | 0.113 | 0.0204 | 0.428 | (-0.167, 0.393) |
| < 1 y | 0.149 | 0.0121 | 0.176 | (-0.067, 0.364) |
| > 20 y | 0.022 | 0.016 | 0.956 | (-0.764, 0.808) |
| **Skin Disease Knowledge (Ref: Less Knowledgeable)** | | | | |
| More Knowledgeable | 0.012 | 0.005 | 0.866 | (-0.126, 0.150) |
| **Other Covariates** | | | | |
| HAI | -0.009 | <0.0001 | 0.921 | (-0.018, 0.001) |
| XAI | 0.012 | 0.0001 | 0.220 | (-0.007, 0.032) |
| AOT | <0.001 | <0.0001 | 0.999 | (-0.015, 0.015) |
| CRT | 0.023 | 0.0008 | 0.442 | (-0.029, 0.075) |
| **Interaction Terms** | | | | |
| Decision Rd. 2 : Explanation Group CBIR | -0.057 | 0.0062 | 0.524 | (-0.231, 0.118) |
| Decision Rd. 2 : Explanation Group GradCAM | -0.034 | 0.0071 | 0.688 | (-0.201, 0.132) |
| Decision Rd. 2 : Explanation Group LLM | -0.111 | 0.0062 | 0.189 | (-0.276, 0.054) |

**STable 19 | Linear Mixed Model Results on Overall Diagnostic Performance in PCPs (Target Outcome: Top-1 Accuracy, Lyme Group)**

| Variables | β | $\sigma^2$ | p-value | 95% CI for β | Sig. Level |
|---|---|---|---|---|---|
| **Intercept** | -0.136 | 0.0185 | 0.725 | (-0.458, 0.185) | |
| **Decision (Ref: Rd. 1 without AI)** | | | | | |
| Rd. 2 with AI Assistance | 0.260 | 0.0039 | <0.001 | (0.137, 0.384) | *** |
| **Explanation Group (Ref: Basic AI)** | | | | | |
| CBIR | 0.154 | 0.0098 | 0.117 | (-0.058, 0.367) | |
| GradCAM | 0.127 | 0.0088 | 0.176 | (-0.057, 0.311) | |
| LLM | 0.126 | 0.0086 | 0.176 | (-0.056, 0.308) | |
| **Age (Ref: 18-24)** | | | | | |
| 25 - 34 | -0.184 | 0.0240 | 0.125 | (-0.419, 0.051) | |
| 35 - 44 | -0.172 | 0.0296 | 0.288 | (-0.491, 0.146) | |

| | | | | | |
|---|---|---|---|---|---|
| 45 - 54 | -0.156 | 0.0296 | 0.470 | (-0.581, 0.268) | |
| 55 - 64 | 0.165 | 0.1521 | 0.672 | (-0.599, 0.929) | |
| 65 or older | 0.662 | 0.2052 | 0.144 | (-0.226, 1.550) | |
| Under 18 | -0.067 | 0.0169 | 0.869 | (-0.859, 0.725) | |
| **Gender (Ref: Female)** | | | | | |
| Male | -0.083 | 0.0042 | 0.200 | (-0.211, 0.044) | |
| Other | -0.044 | 0.0400 | 0.827 | (-0.441, 0.352) | |
| **Race (Ref: American Indian)** | | | | | |
| Asian | 0.433 | 0.0841 | 0.143 | (-0.147, 1.012) | |
| Black/African American | 0.414 | 0.0961 | 0.180 | (-0.191, 1.019) | |
| Hispanic/Latino/Spanish Origin | 0.320 | 0.0961 | 0.333 | (-0.327, 0.966) | |
| Other | 0.383 | 0.0955 | 0.201 | (-0.208, 0.977) | |
| White | 0.393 | 0.0955 | 0.203 | (-0.212, 0.998) | |
| **Year of Medical Experience (Ref: 1-3 y)** | | | | | |
| 10 - 20 y | 0.076 | 0.0213 | 0.601 | (-0.209, 0.361) | |
| 5 - 10 y | 0.031 | 0.0058 | 0.696 | (-0.123, 0.184) | |
| 0 - 5 y | 0.052 | 0.0156 | 0.675 | (-0.192, 0.297) | |
| < 1 y | 0.146 | 0.0213 | 0.129 | (-0.042, 0.334) | |
| > 20 y | -0.386 | 0.1225 | 0.271 | (-1.072, 0.300) | |
| **Skin Disease Knowledge (Ref: Less Knowledgeable)** | | | | | |
| More Knowledgeable | 0.087 | 0.0037 | 0.157 | (-0.033, 0.207) | |
| **Other Covariates** | | | | | |
| HAI | 0.016 | <0.0001 | 0.039 | (0.001, 0.031) | * |
| XAI | 0.004 | <0.0001 | 0.690 | (-0.004, 0.012) | |
| AOT | -0.001 | <0.0001 | 0.832 | (-0.014, 0.012) | |
| CRT | 0.021 | 0.0007 | 0.409 | (-0.029, 0.071) | |
| **Interaction Terms** | | | | | |
| Decision Rd. 2 : Explanation Group CBIR | -0.087 | 0.0077 | 0.236 | (-0.291, 0.117) | |
| Decision Rd. 2 : Explanation Group GradCAM | -0.067 | 0.0077 | 0.448 | (-0.240, 0.106) | |
| Decision Rd. 2 : Explanation Group LLM | -0.075 | 0.0077 | 0.395 | (-0.246, 0.097) | |

**STable 20 | Linear Mixed Model Results on Overall Diagnostic Performance in PCPs (Target Outcome: Top-1 Accuracy, CTCL Group)**

| Variables | β | $σ^2$ | p-value | 95% CI for β | Sig. Level |
|---|---|---|---|---|---|
| **Intercept** | -0.331 | 0.0812 | 0.246 | (-0.890, 0.228) | |
| **Decision (Ref: Rd. 1 without AI)** | | | | | |
| Rd. 2 with AI Assistance | 0.212 | 0.0042 | 0.001 | (0.085, 0.338) | ** |
| **Explanation Group (Ref: Basic AI)** | | | | | |
| CBIR | 0.040 | 0.0064 | 0.613 | (-0.116, 0.196) | |
| GradCAM | 0.032 | 0.0058 | 0.671 | (-0.117, 0.182) | |
| LLM | 0.056 | 0.0056 | 0.457 | (-0.091, 0.203) | |
| **Age (Ref: 18-24)** | | | | | |
| 25 - 34 | -0.022 | 0.0158 | 0.804 | (-0.195, 0.151) | |
| 35 - 44 | -0.083 | 0.0256 | 0.487 | (-0.318, 0.152) | |
| 45 - 54 | -0.153 | 0.0256 | 0.338 | (-0.466, 0.160) | |
| 55 - 64 | 0.027 | 0.0784 | 0.925 | (-0.537, 0.591) | |
| 65 or older | 0.096 | 0.0829 | 0.773 | (-0.559, 0.752) | |
| Under 18 | 0.127 | 0.1089 | 0.670 | (-0.569, 0.825) | |
| **Gender (Ref: Female)** | | | | | |
| Male | -0.028 | 0.0023 | 0.564 | (-0.122, 0.066) | |
| Other | -0.026 | 0.0144 | 0.861 | (-0.319, 0.267) | |
| **Race (Ref: American Indian)** | | | | | |
| Asian | 0.472 | 0.0515 | 0.031 | (0.044, 0.900) | * |
| Black/African American | 0.381 | 0.0520 | 0.095 | (-0.066, 0.828) | |
| Hispanic/Latino/Spanish Origin | 0.457 | 0.0595 | 0.061 | (-0.020, 0.934) | |
| Other | 0.414 | 0.0484 | 0.066 | (-0.027, 0.856) | |
| White | 0.486 | 0.0520 | 0.033 | (0.040, 0.933) | * |
| **Year of Medical Experience (Ref: 1-3 y)** | | | | | |
| 10 - 20 y | 0.051 | 0.0213 | 0.632 | (-0.159, 0.262) | |
| 5 - 10 y | -0.008 | 0.0117 | 0.886 | (-0.122, 0.105) | |
| 0 - 5 y | 0.053 | 0.0085 | 0.724 | (-0.148, 0.213) | |
| < 1 y | 0.010 | 0.0125 | 0.893 | (-0.129, 0.148) | |
| > 20 y | -0.187 | 0.0676 | 0.469 | (-0.724, 0.350) | |
| **Skin Disease Knowledge (Ref: Less Knowledgeable)** | | | | | |
| More Knowledgeable | 0.072 | 0.0020 | 0.111 | (-0.017, 0.161) | |
| **Other Covariates** | | | | | |

| | | | | |
|---|---|---|---|---|
| HAI | 0.010 | <0.0001 | 0.077 | (0.001, 0.022) |
| XAI | 0.004 | <0.0001 | 0.562 | (-0.009, 0.017) |
| AOT | -0.009 | <0.0001 | 0.076 | (-0.018, 0.001) |
| CRT | -0.005 | <0.0001 | 0.803 | (-0.042, 0.032) |
| **Interaction Terms** | | | | |
| Decision Rd. 2 : Explanation Group CBIR | 0.074 | 0.0020 | 0.434 | (-0.111, 0.259) |
| Decision Rd. 2 : Explanation Group GradCAM | -0.002 | 0.0081 | 0.987 | (-0.179, 0.175) |
| Decision Rd. 2 : Explanation Group LLM | 0.019 | 0.0064 | 0.832 | (-0.157, 0.194) |

**STable 21 | Linear Mixed Model Results on Overall Diagnostic Performance in PCP (Target Outcome: Confidence)**

| Variables | β | $\sigma^2$ | p-value | 95% CI for β | Sig. Level |
|---|---|---|---|---|---|
| **Intercept** | 0.388 | 0.0520 | 0.089 | (-0.059, 0.834) | |
| **Decision (Ref: Rd. 1 without AI)** | | | | | |
| Rd. 2 with AI Assistance | 0.018 | 0.0001 | 0.131 | (-0.005, 0.042) | |
| **Explanation Group (Ref: Basic AI)** | | | | | |
| CBIR | -0.031 | 0.0025 | 0.547 | (-0.134, 0.071) | |
| GradCAM | 0.035 | 0.0030 | 0.487 | (-0.063, 0.133) | |
| LLM | 0.032 | 0.0023 | 0.513 | (-0.064, 0.129) | |
| **Age (Ref: 18-24)** | | | | | |
| 25 - 34 | -0.073 | 0.0127 | 0.083 | (-0.263, 0.016) | |
| 35 - 44 | -0.140 | 0.0117 | 0.147 | (-0.329, 0.049) | |
| 45 - 54 | -0.210 | 0.0441 | 0.102 | (-0.465, 0.042) | |
| 55 - 64 | -0.231 | 0.0534 | 0.318 | (-0.685, 0.223) | |
| 65 or older | -0.565 | 0.0620 | 0.036 | (-1.093, -0.038) | * |
| Under 18 | -0.376 | 0.0600 | 0.118 | (-0.846, 0.095) | |
| **Gender (Ref: Female)** | | | | | |
| Male | 0.095 | 0.0058 | 0.014 | (0.020, 0.171) | * |
| Other | -0.191 | 0.0365 | 0.111 | (-0.427, 0.044) | |
| **Race (Ref: American Indian)** | | | | | |
| Asian | -0.004 | 0.0276 | 0.980 | (-0.349, 0.340) | |

| | | | | | |
|---|---|---|---|---|---|
| Black/African American | -0.002 | 0.0335 | 0.992 | (-0.361, 0.358) | |
| Hispanic/Latino/Spanish Origin | -0.084 | 0.0376 | 0.667 | (-0.468, 0.300) | |
| Other | -0.032 | 0.0296 | 0.860 | (-0.327, 0.263) | |
| White | -0.181 | 0.0324 | 0.522 | (-0.577, 0.215) | |
| **Year of Medical Experience (Ref: 1-3 y)** | | | | | |
| 10 - 20 y | 0.061 | 0.0074 | 0.477 | (-0.108, 0.231) | |
| 5 - 10 y | -0.003 | 0.0056 | 0.726 | (-0.107, 0.102) | |
| 0 - 5 y | -0.083 | 0.0055 | 0.263 | (-0.228, 0.062) | |
| < 1 y | 0.049 | 0.0049 | 0.386 | (-0.062, 0.161) | |
| > 20 y | 0.189 | 0.0437 | 0.363 | (-0.219, 0.597) | |
| **Skin Disease Knowledge (Ref: Less Knowledgeable)** | | | | | |
| More Knowledgeable | 0.137 | 0.0013 | <0.001 | (0.065, 0.208) | *** |
| **Other Covariates** | | | | | |
| HAI | 0.003 | <0.0001 | 0.505 | (-0.006, 0.012) | |
| XAI | 0.004 | <0.0001 | 0.432 | (-0.006, 0.014) | |
| AOT | 0.012 | <0.0001 | 0.002 | (0.005, 0.020) | ** |
| CRT | -0.016 | <0.0001 | 0.280 | (-0.046, 0.013) | |
| **Interaction Terms** | | | | | |
| Decision Rd. 2 : Explanation Group CBIR | 0.005 | 0.0003 | 0.758 | (-0.029, 0.040) | |
| Decision Rd. 2 : Explanation Group GradCAM | 0.010 | 0.0003 | 0.569 | (-0.024, 0.043) | |
| Decision Rd. 2 : Explanation Group LLM | 0.017 | 0.0003 | 0.322 | (-0.016, 0.050) | |
| **Post-hoc Pairwise EMMs Comparisons – XAI Explanations (Rd. 1 vs. Rd. 2)** | | | | | |
| Basic AI (Rd. 1 vs. Rd. 2) | 0.018 | 0.0001 | 0.131 | (-0.005, 0.042) | |
| CBIR (Rd. 1 vs. Rd. 2) | 0.024 | 0.0001 | 0.066 | (-0.002, 0.049) | |
| GradCAM (Rd. 1 vs. Rd. 2) | 0.028 | 0.0001 | 0.019 | (0.005, 0.051) | * |
| LLM (Rd. 1 vs. Rd. 2) | 0.035 | 0.0001 | 0.003 | (0.012, 0.058) | ** |

**STable 22 | Linear Mixed Model Results on Diagnostic Performance Across Skin Tones in PCP (Target Outcome: Top-1 Accuracy)**

| **Variables** | **β** | **$\sigma^2$** | **p-value** | **95% CI for β** | **Sig. Level** |
|---|---|---|---|---|---|
| **Intercept** | 0.041 | 0.0548 | 0.862 | (-0.419, 0.500) | |

| | | | | | |
|---|---|---|---|---|---|
| **Decision (Ref: Rd. 1 without AI)** | | | | | |
| Rd. 2 with AI Assistance | 0.223 | 0.0006 | <0.001 | (0.173, 0.272) | *** |
| **Skin Tone (Ref: Dark Skin)** | | | | | |
| Light | 0.046 | 0.0006 | 0.069 | (-0.004, 0.095) | |
| **Age (Ref: 18-24)** | | | | | |
| 25 - 34 | -0.125 | 0.0055 | 0.091 | (-0.270, 0.020) | |
| 35 - 44 | -0.176 | 0.0102 | 0.076 | (-0.369, 0.018) | |
| 45 - 54 | -0.147 | 0.0177 | 0.269 | (-0.407, 0.113) | |
| 55 - 64 | 0.016 | 0.0557 | 0.945 | (-0.448, 0.480) | |
| 65 or older | 0.121 | 0.0724 | 0.665 | (-0.426, 0.668) | |
| Under 18 | -0.311 | 0.1089 | 0.202 | (-0.788, 0.166) | |
| **Gender (Ref: Female)** | | | | | |
| Male | -0.067 | 0.0045 | 0.090 | (-0.143, 0.010) | |
| Other | -0.066 | 0.0156 | 0.596 | (-0.310, 0.178) | |
| **Race (Ref: American Indian)** | | | | | |
| Asian | 0.078 | 0.0331 | 0.669 | (-0.279, 0.435) | |
| Black/African American | 0.100 | 0.0369 | 0.603 | (-0.276, 0.476) | |
| Hispanic/Latino/Spanish Origin | 0.008 | 0.0400 | 0.967 | (-0.385, 0.400) | |
| Other | 0.091 | 0.0353 | 0.629 | (-0.278, 0.479) | |
| White | 0.104 | 0.0376 | 0.585 | (-0.270, 0.478) | |
| **Year of Medical Experience (Ref: 1-3 y)** | | | | | |
| 10 - 20 y | 0.065 | 0.0079 | 0.464 | (-0.109, 0.239) | |
| 5 - 10 y | 0.045 | 0.002 | 0.351 | (-0.049, 0.139) | |
| 0 - 5 y | 0.048 | 0.0023 | 0.525 | (-0.100, 0.196) | |
| < 1 y | 0.071 | 0.0035 | 0.225 | (-0.044, 0.187) | |
| > 20 y | -0.170 | 0.0441 | 0.417 | (-0.581, 0.241) | |
| **Skin Disease Knowledge (Ref: Less Knowledgeable)** | | | | | |
| More Knowledgeable | 0.042 | 0.0014 | 0.252 | (-0.030, 0.115) | |
| **Other Covariates** | | | | | |
| HAI | 0.009 | <0.0001 | 0.070 | (-0.001, 0.018) | |
| XAI | 0.006 | <0.0001 | 0.216 | (-0.004, 0.017) | |
| AOT | -0.003 | <0.0001 | 0.448 | (-0.011, 0.005) | |
| CRT | 0.017 | 0.0003 | 0.283 | (-0.014, 0.048) | |
| **Interaction Terms** | | | | | |
| Decision Rd. 2 : Skin Tone Light | -0.017 | 0.0013 | 0.641 | (-0.087, 0.053) | |

| | | | | |
|---|---|---|---|---|
| **Post-hoc Pairwise EMMs Comparisons – Decision Rounds (Light vs. Dark Skins)** | | | | |
| Rd. 1 (Light vs. Dark Skins) | 0.046 | 0.0006 | 0.069 | (-0.004, 0.095) |
| Rd. 2 (Light vs. Dark Skins) | 0.029 | 0.0006 | 0.248 | (-0.020, 0.079) |

#### 6.5. Finding: PCPs are resilient to AI deference when AI is wrong

**STable 23** presents the Linear Mixed Model results on the change in Top-1 Accuracy after AI assistance, investigating the effect of AI correctness and explanation type. The model reveals a strong, significant negative effect ($p < 0.001$) on accuracy change when the AI is incorrect, confirming that PCPs' performance drops when the AI provides wrong advice. However, the magnitude of the drop is less than a complete deference to the wrong AI advice, demonstrating resilience. The LLM explanation group showed a trend towards a smaller accuracy drop when the AI was wrong compared to the Basic AI group, indicated by a positive interaction term ($\beta = 0.128$) with a borderline significance ($p=0.117$). **STable 24** provides post-hoc pairwise comparisons to further examine the influence of LLM-based explanations on accuracy change under correct and incorrect AI advice. Critically, under AI incorrectness, there were no significant differences between the LLM group and other explanation groups (Basic AI, CBIR, GradCAM), suggesting that LLM explanations did not significantly *worsen* the outcome, which supports the idea that PCPs maintain their own judgment.

**STable 26** analyzes the factors influencing accuracy specifically under conditions of AI Correctness, examining the role of XAI Quality (High vs. Low) and Explanation Type. When the AI was correct, the GradCAM group with High XAI Quality showed a significantly lower accuracy than the CBIR group ($p < 0.001$), suggesting that high-quality GradCAM explanations did not translate to superior performance compared to CBIR. Conversely, the negative impact of incorrect AI was significant ($p = 0.002$), highlighting the detrimental effect of erroneous AI recommendations on performance. **STable 27** offers EMM results of XAI Quality across different explanation types and AI correctness states. The comparisons reinforce that under AI-Correct scenarios, the LLM group with High XAI Quality performed significantly worse than the GradCAM group ($p < 0.001$), indicating complexity in how explanation quality impacts outcomes. Conversely, under AI-Incorrect scenarios, the differences between LLM and other explanations across both high and low XAI quality were non-significant with seldom diagnostic accuracy change, further supporting the resilience of PCPs to poor explanations when the AI is wrong.

**STable 23 | Linear Mixed Model Results on Diagnostic Performance Across AI Correctness and Explanations in PCP (Target Outcome: Top-1 Accuracy Change after AI Assistance)**

| **Variables** | **β** | **$\sigma^2$** | **p-value** | **95% CI for β** | **Sig. Level** |
|---|---|---|---|---|---|
| **Intercept** | -0.011 | 0.0437 | 0.960 | (-0.440, 0.418) | |

| | | | | | |
|---|---|---|---|---|---|
| **AI Correctness (Ref: Correct)** | | | | | |
| Wrong | -0.349 | 0.0035 | <0.001 | (-0.464, -0.234) | *** |
| **Explanation (Ref: Basic)** | | | | | |
| CBIR | -0.048 | 0.0031 | 0.462 | (-0.176, 0.080) | |
| GradCAM | -0.035 | 0.0038 | 0.570 | (-0.158, 0.087) | |
| LLM | -0.100 | 0.0038 | 0.106 | (-0.221, 0.021) | |
| **Race (Ref: American Indian)** | | | | | |
| Asian | 0.360 | 0.0279 | 0.031 | (0.032, 0.688) | * |
| Black/African American | 0.380 | 0.0306 | 0.029 | (0.038, 0.723) | * |
| Hispanic/Latino/Spanish Origin | 0.392 | 0.0313 | 0.036 | (0.036, 0.757) | * |
| Other | 0.352 | 0.0296 | 0.041 | (0.014, 0.690) | * |
| White | 0.360 | 0.0306 | 0.039 | (0.018, 0.702) | * |
| **Gender (Ref: Female)** | | | | | |
| Male | -0.019 | 0.0014 | 0.613 | (-0.087, 0.054) | |
| Other | 0.006 | 0.0132 | 0.956 | (-0.218, 0.231) | |
| **Age (Ref: 18-24)** | | | | | |
| 25 - 34 | -0.069 | 0.0077 | 0.308 | (-0.202, 0.064) | |
| 35 - 44 | -0.138 | 0.0085 | 0.132 | (-0.318, 0.042) | |
| 45 - 54 | -0.128 | 0.0150 | 0.297 | (-0.368, 0.112) | |
| 55 - 64 | 0.010 | 0.0493 | 0.964 | (-0.422, 0.442) | |
| 65 or older | -0.078 | 0.0655 | 0.760 | (-0.397, 0.242) | |
| Under 18 | 0.038 | 0.0524 | 0.868 | (-0.410, 0.486) | |
| **Skin Disease Knowledge (Ref: Less Knowledgeable)** | | | | | |
| More Knowledgeable | -0.026 | 0.0012 | 0.453 | (-0.094, 0.042) | |
| **Year of Medical Experience (Ref: 1-3 y)** | | | | | |
| 10 - 20 y | 0.092 | 0.0067 | 0.265 | (-0.070, 0.253) | |
| 5 - 10 y | 0.007 | 0.0020 | 0.883 | (-0.088, 0.093) | |
| 0 - 5 y | 0.092 | 0.0051 | 0.190 | (-0.046, 0.231) | |
| < 1 y | -0.003 | 0.0030 | 0.952 | (-0.110, 0.103) | |
| > 20 y | -0.065 | 0.0384 | 0.741 | (-0.454, 0.323) | |
| **Interaction Terms** | | | | | |
| AI correctness wrong : explanation CBIR | 0.087 | 0.0074 | 0.312 | (-0.082, 0.256) | |
| AI correctness wrong : explanation GradCAM | 0.079 | 0.0067 | 0.338 | (-0.083, 0.240) | |
| AI correctness wrong : explanation LLM | 0.128 | 0.0067 | 0.117 | (0.011, 0.245) | |
| **Other Covariates** | | | | | |

| | | | | |
|---|---|---|---|---|
| HAI | 0.008 | <0.0001 | 0.071 | (0.001, 0.017) |
| XAI | 0.006 | <0.0001 | 0.196 | (-0.003, 0.016) |
| AOT | -0.020 | <0.0001 | 0.672 | (-0.048, -0.003) |
| CRT | 0.002 | <0.0001 | 0.175 | (-0.009, 0.012) |

**STable 24 | Post-hoc Pairwise EMMs Comparisons – AI Correctness (XAI Pairwise Comparison) based on STable 23**

| Variables | β | $\sigma^2$ | p-value | 95% CI for β | Sig. Level |
|---|---|---|---|---|---|
| **AI Correct (LLM Compared with:)** | | | | | |
| Basic AI | -0.100 | 0.0038 | 0.105 | (-0.221, 0.021) | |
| CBIR | -0.052 | 0.0045 | 0.442 | (-0.183, 0.080) | |
| GradCAM | -0.064 | 0.0039 | 0.302 | (-0.186, 0.058) | |
| **AI Incorrect (LLM Compared with:)** | | | | | |
| Basic AI | 0.028 | 0.0038 | 0.649 | (-0.093, 0.149) | |
| CBIR | -0.011 | 0.0045 | 0.870 | (-0.143, 0.121) | |
| GradCAM | –0.015 | 0.0039 | 0.804 | (-0.138, 0.107) | |

**STable 25 | Linear Mixed Model Results on Differential Portion Across Medical Expertise among PCPs and Medical Students (Target Outcome: Deferential Portion)**

| Variables | β | $\sigma^2$ | p-value | 95% CI for β | Sig. Level |
|---|---|---|---|---|---|
| **Intercept** | 0.515 | 0.0346 | 0.006 | ( 0.150, 0.880) | ** |
| **Medical Role (Ref: Medical Student)** | | | | | |
| PCP | -0.067 | 0.0010 | 0.037 | (-0.130, -0.004) | * |
| **Skin Disease Knowledge (Ref: Less Knowledgeable)** | | | | | |
| More Knowledgeable | -0.073 | 0.0007 | 0.005 | (-0.124, -0.022) | ** |
| **Year of Medical Experience (Ref: 1-3 y)** | | | | | |
| 10 - 20 y | -0.164 | 0.0052 | 0.024 | (-0.306, -0.022) | * |
| 5 - 10 y | -0.040 | 0.0012 | 0.264 | (-0.109, 0.030) | |
| 0 - 5 y | -0.128 | 0.0027 | 0.013 | (-0.229, -0.027) | * |
| < 1 y | -0.044 | 0.0012 | 0.192 | (-0.110, 0.022) | |
| > 20 y | -0.031 | 0.0144 | 0.796 | (-0.267, 0.205) | |
| **Race (Ref: American Indian)** | | | | | |
| Asian | 0.329 | 0.0266 | 0.043 | ( 0.011, 0.648) | * |
| Black or African American | 0.262 | 0.0269 | 0.109 | (-0.058, 0.583) | |

| | | | | | |
|---|---|---|---|---|---|
| Hispanic or Latino or Spanish Origin | 0.262 | 0.0303 | 0.132 | (-0.079, 0.602) | |
| Other | 0.330 | 0.0269 | 0.044 | ( 0.009, 0.650) | * |
| White | 0.256 | 0.0269 | 0.119 | (-0.066, 0.578) | |
| **Gender (Ref: Female)** | | | | | |
| Male | 0.049 | 0.0007 | 0.067 | (-0.003, 0.102) | |
| Other | -0.252 | 0.0086 | 0.007 | (-0.434, -0.070) | ** |
| **Age (Ref: 18-24)** | | | | | |
| 25 - 34 | 0.065 | 0.0010 | 0.045 | ( 0.002, 0.128) | * |
| 35 - 44 | 0.110 | 0.0030 | 0.045 | ( 0.002, 0.218) | * |
| 45 - 54 | -0.157 | 0.0062 | 0.048 | (-0.312, -0.002) | * |
| 55 - 64 | 0.123 | 0.0110 | 0.245 | (-0.084, 0.329) | |
| 65 or older | 0.206 | 0.0645 | 0.417 | (-0.292, 0.704) | |
| Under 18 | 0.576 | 0.0740 | 0.034 | ( 0.042, 1.109) | * |
| **Other Covariates** | | | | | |
| HAI | 0.004 | 0.0000 | 0.282 | (-0.003, 0.010) | |
| XAT | 0.002 | 0.0000 | 0.478 | (-0.004, 0.009) | |
| AOT | 0.007 | 0.0000 | 0.006 | ( 0.002, 0.012) | ** |
| CRT | -0.044 | 0.0001 | <0.001 | (-0.066, -0.022) | *** |

**STable 26| Linear Mixed Model Results on Diagnostic Performance Across AI Explanations and Their Quality in PCP (Target Outcome: Accuracy)**

| Variables | β | $\sigma^2$ | p-value | 95% CI for β | Sig. Level |
|---|---|---|---|---|---|
| **Intercept** | -0.109 | 0.0844 | 0.708 | (-0.678, 0.461) | |
| **AI Correctness (Ref: AI Correct)** | | | | | |
| AI Incorrect | -0.271 | 0.0078 | 0.002 | (-0.443, -0.098) | *** |
| **XAI Quality (Ref: High)** | | | | | |
| Low | 0.008 | 0.0055 | 0.914 | (-0.138, 0.154) | |
| **Explanation (Ref: CBIR)** | | | | | |
| GradCAM | 0.551 | 0.026 | 0.001 | (0.235, 0.867) | ** |
| LLM | -0.050 | 0.0067 | 0.540 | (-0.210, 0.110) | |
| **Race (Ref: American Indian)** | | | | | |
| Asian | 0.386 | 0.0422 | 0.060 | (-0.016, 0.789) | |
| Black/African American | 0.443 | 0.0507 | 0.049 | (0.002, 0.884) | * |
| Hispanic/Latino/Spanish Origin | 0.508 | 0.0560 | 0.032 | (0.045, 0.972) | * |

| | | | | | |
|---|---|---|---|---|---|
| Other | 0.384 | 0.0483 | 0.080 | (-0.046, 0.815) | |
| White | 0.444 | 0.0509 | 0.049 | (0.002, 0.886) | * |
| **Gender (Ref: Female)** | | | | | |
| Male | 0.026 | 0.0035 | 0.656 | (-0.090, 0.143) | |
| Other | 0.039 | 0.0176 | 0.769 | (-0.221, 0.299) | |
| **Age (Ref: 18-24)** | | | | | |
| 25 - 34 | -0.071 | 0.0080 | 0.426 | (-0.246, 0.104) | |
| 35 - 44 | -0.040 | 0.0155 | 0.747 | (-0.285, 0.204) | |
| 45 - 54 | 0.010 | 0.0255 | 0.951 | (-0.303, 0.323) | |
| 55 - 64 | 0.131 | 0.0787 | 0.642 | (-0.419, 0.680) | |
| 65 or older | 0.062 | 0.1063 | 0.85 | (-0.577, 0.701) | |
| Under 18 | 0.059 | 0.0718 | 0.826 | (-0.466, 0.584) | |
| **Skin Disease Knowledge (Ref: Less Knowledgeable)** | | | | | |
| More Knowledgeable | -0.006 | 0.0025 | 0.902 | (-0.104, 0.092) | |
| **Year of Medical Experience (Ref: 1-3 y)** | | | | | |
| 10 - 20 y | 0.003 | 0.0130 | 0.981 | (-0.221, 0.226) | |
| 5 - 10 y | 0.007 | 0.0043 | 0.914 | (-0.121, 0.135) | |
| 0 - 5 y | 0.024 | 0.0106 | 0.816 | (-0.178, 0.225) | |
| < 1 y | -0.048 | 0.0065 | 0.55 | (-0.205, 0.110) | |
| > 20 y | -0.306 | 0.0646 | 0.228 | (-0.804, 0.192) | |
| **Interaction Terms** | | | | | |
| XAI quality Low : explanation GradCAM | 0.481 | 0.0642 | 0.058 | (-0.015, 0.978) | |
| XAI quality Low : explanation LLM | 0.054 | 0.0221 | 0.716 | (-0.237, 0.346) | |
| **Other Covariates** | | | | | |
| HAI | 0.013 | <0.0001 | 0.033 | (0.001, 0.024) | * |
| XAI | 0.006 | 0.0001 | 0.478 | (-0.010, 0.021) | |
| CRT | -0.036 | 0.0004 | 0.063 | (-0.073, 0.002) | |
| AOT | <0.001 | <0.0001 | 0.953 | (-0.011, 0.011) | |

**STable 27 | Post-hoc Pairwise EMMs Comparisons – XAI Quality (XAI Pairwise Comparison)**

| Condition | Variables | β | $\sigma^2$ | p-value | 95% CI for β | Sig. Level |
|---|---|---|---|---|---|---|
| AI Correct | **AI Explanation Quality High (LLM Compared with:)** | | | | | |
| | CBIR | -0.050 | 0.0067 | 0.540 | (-0.210, 0.110) | |
| | GradCAM | -0.602 | 0.0249 | <0.001 | (-0.910, -0.293) | *** |
| | **AI Explanation Quality Low (LLM Compared with:)** | | | | | |
| | CBIR | -0.117 | 0.0081 | 0.193 | (-0.293, 0.059) | |
| | GradCAM | -0.119 | 0.0059 | 0.119 | (-0.268, 0.031) | |
| AI Wrong | **AI Explanation Quality High (LLM Compared with:)** | | | | | |
| | CBIR | -0.003 | 0.0102 | 0.980 | (-0.201, 0.196) | |
| | GradCAM | -0.079 | 0.0318 | 0.660 | (-0.440, 0.271) | |
| | **AI Explanation Quality Low (LLM Compared with:)** | | | | | |
| | CBIR | -0.015 | 0.0079 | 0.863 | (-0.189, 0.159) | |
| | GradCAM | -0.023 | 0.0068 | 0.782 | (-0.185, 0.139) | |

### 6.6. Finding: Higher AI deference correlates with lower initial performance

**STable 28** investigates the diagnostic accuracy of the general public by deferential group. The non-deferential group demonstrated significantly higher initial diagnostic accuracy (Rd. 1) compared to the deferential group (p<0.001). AI assistance significantly improved accuracy for both groups (Rd. 2), but the benefit was greater for the deferential group, thus narrowing the performance gap (interaction effect: p<0.001).

**STable 29** investigates the Top-1 diagnostic accuracy of PCP by deferential group. Non-deferential PCPs showed significantly higher initial Top-1 accuracy compared to deferential PCPs (p<0.001). AI assistance significantly improved accuracy for all PCPs (p<0.001), with a slightly but significantly smaller gain for the non-deferential group (p=0.047) due to their higher baseline performance.

**STable 30** analyzes the factors associated with the deferential proportion among the general public. Older participants (55-64 years, p<0.001) and those with skin disease experience (p=0.001) showed significantly higher deference. Conversely, some minority race/ethnicity groups (Black or African American, Other) showed significantly lower deference than the American Indian reference group.

**STable 31** analyzes the factors associated with the deferential proportion among PCPs. Several race/ethnicity groups (Asian, Black/African American, Hispanic/Latino/Spanish Origin, Other, White) exhibited significantly higher deferential portions compared to the American Indian reference group. PCPs with more medical experience (45-54 years) showed a trend towards lower deference (p=0.094).

**STable 28 | Linear Mixed Model Results on Diagnostic Performance Across Deferential Group in General Public (Target Outcome: Accuracy)**

| Variables | β | $\sigma^2$ | p-value | 95% CI for β | Sig. Level |
|---|---|---|---|---|---|
| **Intercept** | 0.614 | 0.0008 | <0.001 | (0.557, 0.670) | *** |
| **Deferential Group (Ref: Deferential)** | | | | | |
| Non-Deferential | 0.082 | 0.0003 | <0.001 | (0.051, 0.114) | *** |
| **Decision (Ref: Rd. 1 without AI)** | | | | | |
| Rd. 2 with AI Assistance | 0.093 | 0.0001 | <0.001 | (0.074, 0.111) | *** |
| **Age (Ref: 18-24)** | | | | | |
| 25 - 34 | -0.004 | 0.0004 | 0.832 | (-0.042, 0.034) | |
| 35 - 44 | -0.003 | 0.0004 | 0.873 | (-0.045, 0.038) | |
| 45 - 54 | 0.039 | 0.0010 | 0.306 | (-0.035, 0.113) | |
| 55 - 64 | 0.078 | 0.0022 | 0.100 | (-0.015, 0.170) | |
| 65 or older | 0.056 | 0.0081 | 0.530 | (-0.120, 0.233) | |
| **Gender (Ref: Female)** | | | | | |
| Male | -0.056 | 0.0005 | 0.005 | (-0.087, -0.024) | ** |
| Other | -0.001 | 0.0006 | 0.989 | (-0.049, 0.047) | |
| **Race (Ref: American Indian)** | | | | | |
| Asian | 0.072 | 0.0014 | 0.049 | (0.000, 0.144) | * |
| Black/African American | 0.024 | 0.0013 | 0.504 | (-0.046, 0.094) | |
| Hispanic/Latino/Spanish Origin | 0.076 | 0.0014 | 0.046 | (0.001, 0.151) | * |
| Native Hawaiian/Other Pacific Islander | 0.022 | 0.0036 | 0.717 | (-0.096, 0.140) | |
| Other | 0.028 | 0.0014 | 0.444 | (-0.044, 0.100) | |
| White | 0.033 | 0.0006 | 0.161 | (-0.013, 0.079) | |
| **Skin Disease Experience (Ref: No)** | | | | | |
| Yes | 0.009 | 0.0002 | 0.574 | (-0.021, 0.039) | |
| **Interaction Terms** | | | | | |
| Deferential Group Non-Deferential : Decision Rd. 2 | -0.055 | 0.0002 | <0.001 | (-0.079, -0.030) | *** |
| **Other Covariates** | | | | | |
| HAI Collaboration Experience | 0.007 | <0.0001 | <0.001 | (0.004, 0.011) | *** |

| | | | | | |
|---|---|---|---|---|---|
| **Post-hoc Pairwise EMMs Comparisons – Decision Rounds (Deferential vs. Non-Deferential)** | | | | | |
| Rd. 1 (Deferential vs. Non-Deferential) | 0.0823 | 0.0003 | <0.001 | (0.051, 0.114) | *** |
| Rd. 2 (Deferential vs. Non-Deferential) | 0.0276 | 0.0003 | 0.083 | (-0.004, 0.059) | |

**STable 29 | Linear Mixed Model Results on Diagnostic Performance Across Deferential Group in PCP (Target Outcome: Top-1 Accuracy)**

| **Variables** | **β** | **$\sigma^2$** | **p-value** | **95% CI for β** | **Sig. Level** |
|---|---|---|---|---|---|
| **Intercept** | -0.067 | 0.0529 | 0.77 | (-0.517, 0.383) | |
| **Decision (Ref: Rd. 1 without AI)** | | | | | |
| Rd. 2 with AI Assistance | 0.235 | 0.0004 | <0.001 | (0.187, 0.284) | *** |
| **Deferential Group (Ref: Deferential)** | | | | | |
| Non-Deferential | 0.194 | 0.0035 | 0.001 | (0.079, 0.310) | ** |
| **Race (Ref: American Indian)** | | | | | |
| Asian | 0.208 | 0.0130 | 0.253 | (0.061, 0.355) | |
| Black/African American | 0.199 | 0.0353 | 0.289 | (-0.035, 0.433) | |
| Hispanic/Latino/Spanish Origin | 0.170 | 0.0420 | 0.407 | (-0.232, 0.573) | |
| Other | 0.210 | 0.0324 | 0.260 | (-0.155, 0.575) | |
| White | 0.206 | 0.0350 | 0.272 | (-0.161, 0.573) | |
| **Gender (Ref: Female)** | | | | | |
| Male | -0.055 | 0.0190 | 0.145 | (-0.279, 0.383) | |
| Other | -0.035 | 0.0144 | 0.768 | (-0.271, 0.200) | |
| **Age (Ref: 18-24)** | | | | | |
| 25 - 34 | -0.098 | 0.0052 | 0.175 | (-0.238, 0.043) | |
| 35 - 44 | -0.164 | 0.0052 | 0.085 | (-0.350, 0.023) | |
| 45 - 54 | -0.185 | 0.0164 | 0.149 | (-0.436, 0.066) | |
| 55 - 64 | 0.011 | 0.0052 | 0.963 | (-0.435, 0.456) | |
| 65 or older | 0.097 | 0.0718 | 0.718 | (-0.429, 0.623) | |
| Under 18 | -0.138 | 0.0595 | 0.570 | (-0.614, 0.338) | |
| **Skin Disease Knowledge (Ref: Less Knowledgeable)** | | | | | |
| More Knowledgeable | 0.034 | 0.0012 | 0.335 | (-0.335, 0.104) | |
| **Year of Medical Experience (Ref: 1-3 y)** | | | | | |
| 10 - 20 y | 0.046 | 0.0072 | 0.591 | (-0.122, 0.213) | |
| 5 - 10 y | 0.033 | 0.0072 | 0.475 | (-0.058, 0.124) | |

| | | | | | |
|---|---|---|---|---|---|
| 0 - 5 y | 0.073 | 0.0024 | 0.749 | (-0.054, 0.200) | |
| < 1 y | 0.045 | 0.0032 | 0.436 | (-0.068, 0.157) | |
| > 20 y | -0.115 | 0.0420 | 0.569 | (-0.512, 0.282) | |
| **Interaction Terms** | | | | | |
| Decision Rd. 2 : Deferential Group Non-Deferential | -0.118 | 0.0035 | 0.047 | (-0.234, -0.002) | * |
| **Other Covariates** | | | | | |
| HAI | 0.010 | <0.0001 | 0.038 | (0.001, 0.018) | * |
| XAI | 0.005 | <0.0001 | 0.298 | (-0.005, 0.015) | |
| CRT | 0.008 | <0.0001 | 0.591 | (-0.022, 0.039) | |
| AOT | -0.003 | <0.0001 | 0.482 | (-0.011, 0.005) | |
| **Post-hoc Pairwise EMMs Comparisons – Decision Rounds (Deferential vs. Non-Deferential)** | | | | | |
| Rd. 1 (Deferential vs. Non-Deferential) | 0.194 | 0.0035 | < 0.001 | (0.079, 0.310) | *** |
| Rd. 2 (Deferential vs. Non-Deferential) | 0.076 | 0.0035 | 0.196 | (-0.039, 0.192) | |

**STable 30 | Linear Mixed Model Results on Differential Portion Across XAI Explanations among General Public (Target Outcome: Deferential Portion)**

| Variables | β | $\sigma^2$ | p-value | 95% CI for β | Sig. Level |
|---|---|---|---|---|---|
| **Intercept** | 0.310 | 0.0066 | <0.001 | (0.151, 0.469) | *** |
| **Explanation Group (Ref: LLM)** | | | | | |
| Basic AI | -0.071 | 0.0027 | 0.169 | (-0.173, 0.030) | |
| CBIR | -0.040 | 0.0027 | 0.433 | (-0.141, 0.060) | |
| GradCAM | -0.074 | 0.0029 | 0.173 | (-0.181, 0.032) | |
| **Race (Ref: American Indian)** | | | | | |
| Asian | -0.006 | 0.0081 | 0.953 | (-0.201, 0.189) | |
| Black or African American | -0.287 | 0.0090 | 0.003 | (-0.474, -0.100) | ** |
| Hispanic/Latino/Spanish Origin | -0.099 | 0.0104 | 0.333 | (-0.298, 0.101) | |
| Native Hawaiian/Other Pacific Islander | -0.176 | 0.0250 | 0.474 | (-0.437, 0.084) | |
| Other | -0.276 | 0.0384 | 0.005 | (-0.469, -0.084) | ** |
| White | -0.090 | 0.0040 | 0.152 | (-0.213, 0.033) | |
| **Age (Ref: 18-24)** | | | | | |
| 25 - 34 | 0.067 | 0.0026 | 0.192 | (-0.034, 0.167) | |
| 35 - 44 | 0.095 | 0.0032 | 0.094 | (-0.016, 0.207) | |

| | | | | | |
|---|---|---|---|---|---|
| 45 - 54 | -0.139 | 0.0102 | 0.170 | (-0.337, 0.059) | |
| 55 - 64 | 0.596 | 0.0154 | <0.001 | (0.352, 0.839) | *** |
| 65 or older | 0.127 | 0.0581 | 0.597 | (-0.345, 0.599) | |
| **Gender (Ref: Female)** | | | | | |
| Male | -0.001 | 0.0014 | 0.985 | (-0.075, 0.074) | |
| Other | 0.269 | 0.0408 | 0.182 | (-0.126, 0.665) | |
| **Skin Disease Experience (Ref: No)** | | | | | |
| Yes | 0.135 | 0.0185 | 0.001 | (0.055, 0.215) | ** |
| **Other Covariates** | | | | | |
| HAI Collaboration Experience | 0.028 | <0.0001 | <0.001 | (0.019, 0.037) | *** |

**STable 31 | Linear Mixed Model Results on Differential Portion Across XAI Explanations among PCPs (Target Outcome: Deferential Portion)**

| **Variables** | **β** | **$\sigma^2$** | **p-value** | **95% CI for β** | **Sig. Level** |
|---|---|---|---|---|---|
| **Intercept** | 0.154 | 0.126 | 0.655 | (-0.521, 0.829) | |
| **Explanation Group (Ref: LLM)** | | | | | |
| Basic AI | 0.082 | 0.0055 | 0.268 | (-0.063, 0.228) | |
| CBIR | 0.046 | 0.0071 | 0.587 | (-0.119, 0.210) | |
| GradCAM | -0.053 | 0.0058 | 0.490 | (-0.202, 0.097) | |
| **Race (Ref: American Indian)** | | | | | |
| Asian | 0.939 | 0.0708 | <0.001 | (0.413, 1.465) | *** |
| Black/African American | 0.722 | 0.0784 | 0.010 | (0.174, 1.271) | ** |
| Hispanic/Latino/Spanish Origin | 1.158 | 0.0894 | <0.001 | (0.572, 1.745) | *** |
| Other | 0.851 | 0.0767 | 0.002 | (0.309, 1.394) | ** |
| White | 0.759 | 0.0784 | 0.007 | (0.211, 1.308) | ** |
| **Gender (Ref: Female)** | | | | | |
| Male | 0.110 | 0.0121 | 0.061 | (-0.005, 0.226) | |
| Other | 0.231 | 0.0338 | 0.209 | (-0.129, 0.590) | |
| **Age (Ref: 18-24)** | | | | | |
| 25 - 34 | 0.183 | 0.0357 | 0.092 | (-0.030, 0.396) | |
| 35 - 44 | 0.035 | 0.0216 | 0.813 | (-0.254, 0.324) | |
| 45 - 54 | -0.329 | 0.0384 | 0.094 | (-0.714, 0.056) | |
| 55 - 64 | -0.185 | 0.1260 | 0.601 | (-0.878, 0.508) | |
| 65 or older | -0.284 | 0.1689 | 0.489 | (-1.090, 0.521) | |

| | | | | | |
|---|---|---|---|---|---|
| Under 18 | 1.189 | 0.1347 | 0.001 | (0.471, 1.908) | ** |
| **Skin Disease Knowledge (Ref: Less Knowledgeable)** | | | | | |
| More Knowledgeable | -0.082 | 0.0231 | 0.141 | (-0.191, 0.027) | |
| **Year of Medical Experience (Ref: 1-3 y)** | | | | | |
| 10 - 20 y | -0.093 | 0.0174 | 0.482 | (-0.352, 0.166) | |
| 5 - 10 y | -0.084 | 0.0135 | 0.237 | (-0.223, 0.055) | |
| 0 - 5 y | -0.135 | 0.0128 | 0.232 | (-0.357, 0.086) | |
| < 1 y | -0.168 | 0.0350 | 0.054 | (-0.338, 0.003) | |
| > 20 y | 0.537 | 0.0980 | 0.091 | (-0.085, 1.160) | |
| **Other Covariates** | | | | | |
| HAI | 0.005 | <0.0001 | 0.441 | (-0.008, 0.018) | |
| XAT | -0.012 | <0.0001 | 0.152 | (-0.027, 0.004) | |
| CRT | -0.068 | 0.0005 | 0.004 | (-0.113, -0.022) | ** |
| AOT | 0.003 | <0.0001 | 0.595 | (-0.009, 0.015) | |

#### 6.7. Finding: Putting AI before human decisions amplifies deference

For the general public in Study 1, **STable 32** examines the effect of HAI paradigm (AI-First vs. Human-First) on diagnostic accuracy. The Human-First paradigm significantly lowered initial (Rd. 1) accuracy compared to the AI-First paradigm (p<0.001), but this performance gap was eliminated after receiving AI assistance (Rd. 2), indicating a significant positive effect of the Human-First paradigm on performance gain (interaction effect: p<0.001). **STable 33** analyzes the diagnostic performance across deferential groups under the AI-First paradigm. The non-deferential group showed a higher trend of accuracy in Round 2 compared to the deferential group (p=0.049). **STable 36** explores the deferential proportion across HAI paradigms and XAI methods. Participants in the AI-First paradigm showed a higher deferential proportion compared to the Human-First paradigm, although the difference was not statistically significant. Several minority race/ethnicity groups (Black or African American, Native Hawaiian/Other Pacific Islander, Other, White) exhibited significantly lower deferential portions compared to the American Indian reference group.

For the PCPs in Study 2, **STable 34** investigates the effect of HAI paradigm on Top-1 diagnostic accuracy. The Human-First paradigm resulted in significantly lower initial (Rd. 1) Top-1 accuracy compared to the AI-First paradigm (p<0.001). However, the Human-First paradigm showed a significantly larger improvement in accuracy after AI assistance (Rd. 2), effectively eliminating the performance gap (interaction effect: p<0.001). **STable 35** analyzes the diagnostic performance across deferential groups under the AI-First paradigm. The non-deferential group maintained a significantly higher accuracy in Round 2 compared to the deferential group (p=0.041). **STable 37** explores the deferential proportion across HAI paradigms and XAI methods. Unlike the general public, no significant difference in deferential proportion was found between the Human-First and AI-First paradigms across XAI groups. However, several race/ethnicity groups (Asian, Hispanic or Latino or Spanish Origin) showed significantly higher deferential portions compared to the American Indian reference group.

**STable 32 | Linear Mixed Model Results on Diagnostic Performance Across HAI Paradigm in General Public (Target Outcome: Accuracy, AI-First vs. Human-First)**

| Variables | β | $\sigma^2$ | p-value | 95% CI for β | Sig. Level |
|---|---|---|---|---|---|
| **Intercept** | 0.737 | 0.0006 | <0.001 | (0.690, 0.783) | *** |
| **Decision (Ref: Rd. 1 without AI)** | | | | | |
| Rd. 2 with AI Assistance | 0.002 | <0.0001 | 0.699 | (-0.009, 0.013) | |
| **HAI Paradigm (Ref: AI First)** | | | | | |
| Human First | -0.054 | 0.0001 | <0.001 | (-0.076, -0.033) | *** |
| **Race (Ref: American Indian)** | | | | | |
| Asian | 0.053 | 0.0008 | 0.006 | (-0.002, 0.107) | |
| Black/African American | 0.004 | 0.0007 | 0.867 | (-0.046, 0.055) | |
| Hispanic/Latino/Spanish Origin | 0.042 | 0.0010 | 0.174 | (-0.039, 0.122) | |
| Native Hawaiian/Other Pacific Islander | -0.019 | 0.0014 | 0.620 | (-0.094, 0.056) | |
| Other | 0.037 | 0.0014 | 0.174 | (-0.016, 0.090) | |
| White | 0.030 | 0.0004 | 0.106 | (-0.006, 0.067) | |
| **Age (Ref: 18-24)** | | | | | |
| 25 - 34 | -0.011 | 0.0002 | 0.460 | (-0.042, 0.019) | |
| 35 - 44 | -0.011 | 0.0003 | 0.477 | (-0.042, 0.020) | |
| 45 - 54 | 0.044 | 0.0007 | 0.090 | (-0.007, 0.096) | |
| 55 - 64 | 0.035 | 0.0016 | 0.379 | (-0.043, 0.112) | |
| 65 or older | -0.031 | 0.0039 | 0.637 | (-0.158, 0.097) | |
| **Gender (Ref: Female)** | | | | | |
| Male | -0.034 | 0.0006 | 0.001 | (-0.054, -0.013) | ** |
| Other | 0.019 | 0.0042 | 0.772 | (-0.109, 0.146) | |
| **Skin Disease Experience (Ref: No)** | | | | | |
| Yes | -0.008 | 0.0001 | 0.446 | (-0.030, 0.013) | |
| **Interaction Terms** | | | | | |
| Decision Rd. 2 : HAI Paradigm Human First | 0.059 | <0.0001 | <0.001 | (0.045, 0.074) | *** |
| **Other Covariates** | | | | | |

| | | | | | |
|---|---|---|---|---|---|
| HAI Collaboration Experience | 0.005 | <0.0001 | < 0.001 | (0.002, 0.007) | *** |
| Decision Time (in minute) | 0.001 | <0.0001 | 0.909 | (-0.014, 0.016) | |
| **Post-hoc Pairwise EMMs Comparisons – Decision Round (Human-First vs. AI-First)** | | | | | |
| Round 1 (Human-First vs. AI-First) | -0.054 | 0.0001 | <0.001 | (-0.076, -0.033) | *** |
| Round 2 (Human-First vs. AI-First) | 0.005 | 0.0001 | 0.650 | (-0.017, 0.026) | |

**STable 33 | Linear Mixed Model Results on Diagnostic Performance Across Deferential Group in General Public (Target Outcome: Accuracy, AI First)**

| **Variables** | **β** | **$\sigma^2$** | **p-value** | **95% CI for β** | **Sig. Level** |
|---|---|---|---|---|---|
| **Intercept** | 0.761 | 0.0014 | <0.001 | (0.687, 0.835) | *** |
| **Deferential Group (Ref: Deferential)** | | | | | |
| Non-Deferential | 0.015 | 0.0003 | 0.335 | (-0.016, 0.046) | |
| **Decision (Ref: Rd. 1 without AI)** | | | | | |
| Rd. 2 with AI Assistance | -0.006 | 0.0000 | 0.214 | (-0.015, 0.003) | |
| **Gender (Ref: Female)** | | | | | |
| Male | -0.027 | 0.0003 | 0.082 | (-0.058, 0.004) | |
| Other | 0.123 | 0.0177 | 0.356 | (-0.139, 0.385) | |
| **Race (Ref: American Indian)** | | | | | |
| Asian | 0.004 | 0.0019 | 0.933 | (-0.083, 0.090) | |
| Black/African American | -0.036 | 0.0014 | 0.354 | (-0.113, 0.040) | |
| Hispanic/Latino/Spanish Origin | -0.033 | 0.0027 | 0.528 | (-0.134, 0.069) | |
| Native Hawaiian/Other Pacific Islander | -0.070 | 0.0025 | 0.184 | (-0.174, 0.033) | |
| Other | 0.013 | 0.0018 | 0.760 | (-0.060, 0.085) | |
| White | 0.008 | 0.0011 | 0.811 | (-0.054, 0.069) | |
| **Age (Ref: 18-24)** | | | | | |
| 25 - 34 | -0.015 | 0.0005 | 0.514 | (-0.059, 0.029) | |
| 35 - 44 | -0.012 | 0.0005 | 0.596 | (-0.058, 0.033) | |
| 45 - 54 | 0.032 | 0.0014 | 0.400 | (-0.042, 0.105) | |
| 55 - 64 | 0.028 | 0.0059 | 0.713 | (-0.123, 0.180) | |
| 65 or older | -0.117 | 0.0151 | 0.211 | (-0.300, 0.066) | |
| **Skin Disease Experience (Ref: No)** | | | | | |
| Yes | -0.017 | 0.0003 | 0.276 | (-0.049, 0.014) | |
| **Interaction Terms** | | | | | |

| | | | | | |
|---|---|---|---|---|---|
| Deferential Group Non-Deferential : Decision Rd. 2 | 0.016 | 0.0000 | 0.012 | (0.003, 0.028) | * |
| **Other Covariates** | | | | | |
| HAI Collaboration Experience | 0.004 | 0.0000 | 0.015 | (0.001, 0.008) | * |
| Decision Time (in minute) | 0.000 | 0.0000 | 0.990 | (-0.000, 0.001) | |
| **Post-hoc Pairwise EMMs Comparisons – Decision Round (Non-Deferential vs. Deferential)** | | | | | |
| Round 1 (Non-Deferential vs. Deferential) | 0.015 | 0.0002 | 0.335 | (-0.016, 0.046) | |
| Round 2 (Non-Deferential vs. Deferential) | 0.031 | 0.0002 | 0.049 | (0.000, 0.062) | * |

**STable 34 | Linear Mixed Model Results on Diagnostic Performance Across HAI Paradigm in PCP (Target Outcome: Top-1 Accuracy, AI-First vs. Human-First)**

| Variables | β | $\sigma^2$ | p-value | 95% CI for β | Sig. Level |
|---|---|---|---|---|---|
| **Intercept** | 0.389 | 0.0384 | 0.047 | (0.005, 0.774) | * |
| **Decision (Ref: Rd. 1 without AI)** | | | | | |
| Rd. 2 with AI Assistance | 0.023 | 0.0005 | 0.468 | (-0.038, 0.084) | |
| **HAI Paradigm (Ref: AI First)** | | | | | |
| Human First | -0.226 | 0.0013 | <0.001 | (-0.297, -0.154) | *** |
| **Race (Ref: American Indian)** | | | | | |
| Asian | -0.150 | 0.0210 | 0.302 | (-0.435, 0.135) | |
| Black/African American | -0.083 | 0.0231 | 0.586 | (-0.380, 0.215) | |
| Hispanic/Latino/Spanish Origin | -0.098 | 0.0266 | 0.550 | (-0.418, 0.223) | |
| Other | -0.125 | 0.0237 | 0.403 | (-0.419, 0.169) | |
| White | -0.117 | 0.0222 | 0.433 | (-0.409, 0.175) | |
| **Gender (Ref: Female)** | | | | | |
| Male | -0.013 | 0.0014 | 0.695 | (-0.080, 0.054) | |
| Other | 0.019 | 0.0135 | 0.874 | (-0.210, 0.247) | |
| **Age (Ref: 18-24)** | | | | | |
| 25 - 34 | -0.046 | 0.0053 | 0.533 | (-0.190, 0.098) | |
| 35 - 44 | -0.083 | 0.0086 | 0.376 | (-0.266, 0.101) | |
| 45 - 54 | -0.061 | 0.0135 | 0.603 | (-0.289, 0.168) | |
| 55 - 64 | 0.081 | 0.0313 | 0.646 | (-0.260, 0.428) | |
| 65 or older | 0.076 | 0.0484 | 0.719 | (-0.302, 0.492) | |
| Under 18 | -0.440 | 0.0590 | 0.070 | (-0.917, 0.037) | |

| | | | | | |
|---|---|---|---|---|---|
| **Skin Disease Knowledge (Ref: Less Knowledgeable)** | | | | | |
| More Knowledgeable | 0.052 | 0.0011 | 0.109 | (-0.012, 0.117) | |
| **Year of Medical Experience (Ref: 1-3 y)** | | | | | |
| 10 - 20 y | 0.031 | 0.0067 | 0.702 | (-0.129, 0.191) | |
| 5 - 10 y | 0.011 | 0.0018 | 0.791 | (-0.073, 0.096) | |
| 0 - 5 y | 0.016 | 0.0039 | 0.790 | (-0.104, 0.137) | |
| < 1 y | 0.062 | 0.0031 | 0.222 | (-0.038, 0.162) | |
| > 20 y | -0.065 | 0.0266 | 0.688 | (-0.385, 0.254) | |
| **Interaction Terms** | | | | | |
| Decision Rd. 2 : HAI Paradigm Human First | 0.205 | 0.0004 | <0.001 | (0.145, 0.266) | *** |
| **Other Covariates** | | | | | |
| HAI | 0.010 | <0.0001 | 0.010 | (0.002, 0.018) | * |
| XAI | 0.004 | <0.0001 | 0.330 | (-0.004, 0.013) | |
| CRT | 0.012 | 0.0002 | 0.430 | (-0.017, 0.040) | |
| AOT | -0.001 | <0.0001 | 0.871 | (-0.008, 0.007) | |
| Decision Time (in minute) | <0.001 | <0.0001 | 0.303 | (-0.000, 0.001) | |
| **Post-hoc Pairwise EMMs Comparisons – Decision Round (Human-First vs. AI-First)** | | | | | |
| Round 1 (Human-First vs. AI-First) | -0.226 | 0.0013 | <0.001 | (-0.297, -0.154) | *** |
| Round 2 (Human-First vs. AI-First) | -0.020 | 0.0013 | 0.572 | (-0.091, 0.050) | |

**STable 35 | Linear Mixed Model Results on Diagnostic Performance Across Deferential Group in PCP (Target Outcome: Top-1 Accuracy, AI First)**

| Variables | β | $\sigma^2$ | p-value | 95% CI for β | Sig. Level |
|---|---|---|---|---|---|
| **Intercept** | 0.512 | 0.3950 | 0.194 | (-0.261, 1.286) | |
| **Deferential Group (Ref: Deferential)** | | | | | |
| Non-Deferential | 0.206 | 0.1060 | 0.053 | (-0.002, 0.415) | |
| **Decision (Ref: Rd. 1 without AI)** | | | | | |
| Rd. 2 with AI Assistance | -0.007 | 0.0020 | 0.001 | (-0.011,-0.003) | ** |
| **Race (Ref: American Indian)** | | | | | |
| Asian | -0.503 | 0.2830 | 0.075 | (-1.058, 0.051) | |
| Black/African American | -0.307 | 0.3060 | 0.316 | (-0.907, 0.293) | |
| Hispanic/Latino/Spanish Origin | -0.211 | 0.3200 | 0.510 | (-0.838, 0.416) | |
| Other | -0.456 | 0.2910 | 0.117 | (-1.026, 0.114) | |

| | | | | | |
|---|---|---|---|---|---|
| White | -0.424 | 0.2830 | 0.134 | (-0.978, 0.131) | |
| **Gender (Ref: Female)** | | | | | |
| Male | 0.032 | 0.0790 | 0.691 | (-0.124, 0.187) | |
| Other | 0.223 | 0.2700 | 0.410 | (-0.306, 0.751) | |
| **Age (Ref: 18-24)** | | | | | |
| 25 - 34 | 0.013 | 0.2010 | 0.948 | (-0.381, 0.407) | |
| 35 - 44 | -0.014 | 0.2360 | 0.952 | (-0.476, 0.448) | |
| 45 - 54 | 0.090 | 0.2990 | 0.765 | (-0.497, 0.676) | |
| 55 - 64 | 0.006 | 0.3980 | 0.987 | (-0.774, 0.787) | |
| 65 or older | -0.002 | 0.4470 | 0.997 | (-0.877, 0.874) | |
| **Skin Disease Knowledge (Ref: Less Knowledgeable)** | | | | | |
| More Knowledgeable | 0.053 | 0.0710 | 0.456 | (-0.086, 0.192) | |
| **Year of Medical Experience (Ref: 1-3 y)** | | | | | |
| 10 - 20 y | 0.100 | 0.2500 | 0.690 | (-0.390, 0.589) | |
| 5 - 10 y | -0.080 | 0.0960 | 0.403 | (-0.269, 0.108) | |
| 0 - 5 y | -0.022 | 0.1270 | 0.861 | (-0.270, 0.226) | |
| < 1 y | 0.009 | 0.1220 | 0.940 | (-0.229, 0.248) | |
| > 20 y | 0.094 | 0.3590 | 0.793 | (-0.609, 0.797) | |
| **Interaction Terms** | | | | | |
| Deferential Group Non-Deferential : Decision Rd. 2 | 0.012 | 0.0040 | 0.009 | ( 0.003, 0.020) | ** |
| **Other Covariates** | | | | | |
| HAI | 0.008 | 0.0090 | 0.383 | (-0.009, 0.025) | |
| XAI | 0.005 | 0.0100 | 0.576 | (-0.014, 0.024) | |
| CRT | 0.018 | 0.0370 | 0.634 | (-0.055, 0.090) | |
| AOT | 0.006 | 0.0090 | 0.491 | (-0.011, 0.023) | |
| Decision Time (in minute) | -0.006 | <0.0001 | <0.001 | (-0.008, -0.003) | *** |
| **Post-hoc Pairwise EMMs Comparisons – Decision Round (Non-Deferential vs. Deferential)** | | | | | |
| Round 1 (Non-Deferential vs. Deferential) | 0.206 | 0.0114 | 0.053 | (-0.002, 0.415) | |
| Round 2 (Non-Deferential vs. Deferential) | 0.218 | 0.0114 | 0.041 | (0.009, 0.426) | * |

**STable 36 | Linear Mixed Model Results on Deferential Proportion Across HAI Paradigms and XAI Methods in General Public (Target Outcome: Deferential Proportion)**

| Variables | β | $\sigma^2$ | p-value | 95% CI for β | Sig. Level |
|---|---|---|---|---|---|
| **Intercept** | 0.485 | 0.0053 | <0.001 | (0.341, 0.629) | *** |

| | | | | | |
|---|---|---|---|---|---|
| **HAI Paradigm (Ref: AI First)** | | | | | |
| Human First | -0.097 | 0.0034 | 0.170 | (-0.210, 0.017) | |
| **Explanation Group (Ref: LLM)** | | | | | |
| Basic AI | -0.048 | 0.0035 | 0.418 | (-0.163, 0.067) | |
| CBIR | -0.033 | 0.0031 | 0.551 | (-0.143, 0.076) | |
| GradCAM | -0.063 | 0.0031 | 0.264 | (-0.173, 0.047) | |
| **Race (Ref: American Indian)** | | | | | |
| Asian | -0.124 | 0.0154 | 0.102 | (-0.274, 0.025) | |
| Black or African American | -0.155 | 0.0112 | 0.027 | (-0.292, -0.017) | * |
| Hispanic or Latino or Spanish Origin | -0.111 | 0.0117 | 0.181 | (-0.274, 0.052) | |
| Native Hawaiian/Other Pacific Islander | -0.262 | 0.0270 | 0.012 | (-0.466, -0.057) | * |
| Other | -0.255 | 0.0106 | 0.001 | (-0.399, -0.111) | ** |
| White | -0.103 | 0.0026 | 0.043 | (-0.203, -0.003) | * |
| **Age (Ref: 18-24)** | | | | | |
| 25 - 34 | 0.077 | 0.0014 | 0.051 | (-0.000, 0.153) | |
| 35 - 44 | 0.104 | 0.0017 | 0.014 | (0.018, 0.187) | * |
| 45 - 54 | -0.053 | 0.0050 | 0.451 | (-0.192, 0.085) | |
| 55 - 64 | 0.575 | 0.0115 | <0.001 | (0.365, 0.785) | *** |
| 65 or older | -0.177 | 0.0310 | 0.314 | (-0.521, 0.167) | |
| **Gender (Ref: Female)** | | | | | |
| Male | -0.042 | 0.0006 | 0.142 | (-0.098, 0.014) | |
| Other | 0.036 | 0.0276 | 0.839 | (-0.310, 0.382) | |
| **Skin Disease Knowledge (Ref: No)** | | | | | |
| Yes | 0.047 | 0.0009 | 0.115 | (-0.011, 0.105) | |
| **Other Covariates** | | | | | |
| HAI | 0.020 | <0.0001 | <0.001 | (0.014, 0.027) | *** |
| Decision Time (in minute) | <0.001 | <0.0001 | 1.000 | (-0.000, 0.000) | |
| **Interaction Terms** | | | | | |
| HAI Paradigm Human First : Explanation Group Basic | -0.023 | 0.0062 | 0.769 | (-0.178, 0.131) | |
| HAI Paradigm Human First : Explanation Group CBIR | -0.025 | 0.0061 | 0.744 | (-0.173, 0.124) | |
| HAI Paradigm Human First : Explanation Group GradCAM | -0.078 | 0.0061 | 0.749 | (-0.178, 0.128) | |

| Post-hoc Pairwise EMMs Comparisons – Explanation Group (Human First vs. AI First) | | | | |
|---|---|---|---|---|
| Basic AI (Human First vs. AI First) | -0.097 | 0.0034 | 0.096 | (-0.210, 0.171) |
| CBIR (Human First vs. AI First) | -0.098 | 0.0029 | 0.067 | (-0.203, 0.007) |
| GradCAM (Human First vs. AI First) | -0.099 | 0.0032 | 0.084 | (-0.210, 0.013) |
| LLM (Human First vs. AI First) | -0.074 | 0.0029 | 0.170 | (-0.179, 0.032) |

**STable 37 | Linear Mixed Model Results on Deferential Proportion Across HAI Paradigms and XAI Methods in PCPs (Target Outcome: Deferential Proportion)**

| Variables | β | $\sigma^2$ | p-value | 95% CI for β | Sig. Level |
|---|---|---|---|---|---|
| **Intercept** | 0.644 | 0.0713 | 0.016 | (0.121, 1.166) | * |
| **HAI Paradigm (Ref: AI First)** | | | | | |
| Human First | -0.121 | 0.0071 | 0.150 | (-0.286, 0.044) | |
| **Explanation Group (Ref: LLM)** | | | | | |
| Basic AI | 0.013 | 0.0083 | 0.895 | (-0.174, 0.199) | |
| CBIR | 0.069 | 0.0125 | 0.547 | (-0.155, 0.294) | |
| GradCAM | 0.014 | 0.0061 | 0.878 | (-0.166, 0.194) | |
| **Race (Ref: American Indian)** | | | | | |
| Asian | 0.412 | 0.0380 | 0.035 | (0.030, 0.794) | * |
| Black or African American | 0.248 | 0.0416 | 0.218 | (-0.147, 0.644) | |
| Hispanic or Latino or Spanish Origin | 0.537 | 0.0475 | 0.014 | (0.111, 0.964) | * |
| Other | 0.324 | 0.0488 | 0.107 | (-0.070, 0.718) | |
| White | 0.360 | 0.0392 | 0.069 | (-0.029, 0.748) | |
| **Age (Ref: 18-24)** | | | | | |
| 25 - 34 | 0.052 | 0.0077 | 0.594 | (-0.140, 0.245) | |
| 35 - 44 | -0.005 | 0.0156 | 0.970 | (-0.249, 0.240) | |
| 45 - 54 | -0.144 | 0.0237 | 0.349 | (-0.446, 0.158) | |
| 55 - 64 | -0.073 | 0.0571 | 0.759 | (-0.539, 0.393) | |
| 65 or older | 0.168 | 0.0718 | 0.558 | (-0.395, 0.731) | |
| **Gender (Ref: Female)** | | | | | |
| Male | -0.006 | 0.0023 | 0.816 | (-0.100, 0.087) | |
| Other | 0.193 | 0.0243 | 0.217 | (-0.130, 0.517) | |
| **Skin Disease Knowledge (Ref: Less Knowledgeable)** | | | | | |
| More Knowledgeable | -0.038 | 0.0031 | 0.384 | (-0.125, 0.048) | |
| **Other Covariates** | | | | | |
| HAI | -0.001 | <0.0001 | 0.297 | (-0.005, 0.016) | |

| | | | | | |
|---|---|---|---|---|---|
| XAI | 0.006 | <0.0001 | 0.853 | (-0.013, 0.011) | |
| CRT | -0.039 | 0.0004 | 0.049 | (-0.077, -0.000) | * |
| AOT | 0.003 | <0.0001 | 0.574 | (-0.007, 0.012) | |
| Decision Time (in minute) | <0.001 | <0.0001 | 1.000 | (0.000, 0.000) | |
| **Interaction Terms** | | | | | |
| HAI Paradigm Human First : Explanation Group Basic | -0.023 | 0.0146 | 0.306 | (-0.113, 0.360) | |
| HAI Paradigm Human First : Explanation Group CBIR | 0.031 | 0.0146 | 0.824 | (-0.244, 0.306) | |
| HAI Paradigm Human First : Explanation Group GradCAM | 0.022 | 0.0142 | 0.850 | (-0.202, 0.245) | |
| **Post-hoc Pairwise EMMs Comparisons – Explanation Group (Human First vs. AI First)** | | | | | |
| Basic AI (Human First vs. AI First) | 0.002 | 0.0076 | 0.978 | (-0.168, 0.173) | |
| CBIR (Human First vs. AI First) | -0.090 | 0.0121 | 0.412 | (-0.305, 0.125) | |
| GradCAM (Human First vs. AI First) | -0.099 | 0.0060 | 0.198 | (-0.251, 0.052) | |
| LLM (Human First vs. AI First) | -0.121 | 0.0071 | 0.150 | (-0.286, 0.044) | |
